# Artificial intelligence-enabled event adjudication: estimating delayed cardiovascular effects of respiratory viruses

**DOI:** 10.1101/2020.11.12.20230706

**Authors:** Shinichi Goto, Max Homilius, Jenine E. John, James G. Truslow, Andreas A. Werdich, Alexander J. Blood, Brian H. Park, Calum A. MacRae, Rahul C. Deo

**Affiliations:** One Brave Idea and Division of Cardiovascular Medicine, Department of Medicine, Brigham and Women’s Hospital, Boston, MA, USA; Harvard Medical School, Boston, MA, USA; Center for Digital Health Innovation and Department of Medicine, University of California San Francisco, San Francisco, CA, USA

## Abstract

Healthcare systems ideally should be able to draw lessons from historical data, including whether common exposures are associated with adverse clinical outcomes. Unfortunately, structured clinical data, such as encounter diagnostic codes in electronic health records, suffer from multiple limitations and biases, limiting effective learning. We hypothesized that a machine learning approach to automate ascertainment of clinical events and disease history from medical notes would improve upon using structured data and enable the estimation of real-world risks. We sought to test this approach to address a timely goal: estimating the delayed risk of adverse cardiovascular events (i.e. after the index infection) in patients infected with respiratory viruses. Using 4,151 cardiologist-labeled notes as gold standard, we trained a series of neural network models to automate event adjudication for heart failure hospitalization, acute coronary syndrome, stroke, and coronary revascularization and to identify past medical history for heart failure. Though performance varied by task, in nearly all cases, our models surpassed the use of structured data in terms of sensitivity for a given specificity level and enabled principled evaluation of classification thresholds, which is typically impossible to do with diagnostic codes. Deploying our models on more than 17 million notes for 267,596 patients across an extensive integrated delivery network, we found that patients infected with respiratory syncytial virus had a 23% increased risk of delayed heart failure hospitalization over a subsequent 4-year period compared with propensity-score matched patients who had the same test but with negative results (p = 0.003, log-rank). In contrast, we found no such increased risk in patients with a positive influenza viral test compared with a negative test (rate ratio 0.98, p = 0.71). We conclude that convolutional neural network-based models enable accurate clinical labeling at scale, thereby unlocking timely insights from unstructured clinical data.

## Introduction

The elusive goal of a learning healthcare system requires accurate clinical information. Structured healthcare data, although increasingly accessible, suffers from biases ubiquitous in observational data, including misclassification bias. In particular, diagnostic codes, widely used for billing and research, typically have low sensitivity and specificity^1, 2^. When used to establish an association of exposures to a clinical outcome, such limitations can lead to misclassified outcomes and interfere with adjustment procedures for confounding and selection bias^3^.

Nonetheless, the detection of such associations is a priority and often requires analysis of large-scale real-world data ^4^. Furthermore, such observational data is often the only choice for urgent clinical questions with potential public health implications. One example of such a pressing topic is the cardiovascular consequences of common respiratory viruses. The immediate adverse impact of respiratory viruses such as influenza, respiratory syncytial virus (RSV), and most recently, severe acute respiratory syndrome coronavirus 2 (SARS-CoV-2) is well recognized and includes acute heart failure^5, 6^, myocardial infarction^7, 8^, myocarditis^9–11^, and stroke^12^. However, one unresolved question is whether there is a delayed increased risk in such patients well after the incident infection.

We sought to address these questions, first developing machine learning methods to ascertain and adjudicate clinical events and past medical history at scale, and then applying these to patient data across multiple institutions within a large integrated delivery network.

## Results

### Building a large compendium of cardiologist-labeled notes

The overview of our approach is shown in **Figure 1**. To train an accurate model for event adjudication using medical notes, we labeled a large compendium of discharge summaries and progress notes. A total of 2,967 discharge summaries (1,372 for derivation set, 592 for validation set and 1,003 for test set) and 1,184 progress notes (588 for derivation set, 231 for validation set and 365 for test set) were obtained from the Massachusetts General Brigham (MGB) Enterprise Data Warehouse (EDW) database, which represents multiple institutions across an integrated delivery network. Each discharge summary was manually annotated by cardiologists following previously published guidelines for 4 possible diagnoses^13^: acute coronary syndrome (ACS), heart failure (HF) as the primary cause of hospitalization, percutaneous coronary intervention or coronary artery bypass graft surgery (PCI/CABG), and ischemic stroke. Progress notes were labeled according to a 4-class system for heart failure history (see Methods). Discharge summaries were derived from 17 different institutions while progress notes were from 46 different institutions with differing levels of specialized care **(Table S1 and S2)**. The labeled discharge summaries were then used to train the event models and the progress notes were used to train the note-level past-history model **(Figure S1)**. Because single notes are not exhaustive in documenting the past medical history of a patient, we developed a model that integrated the output from multiple notes to estimate past-history of a specific disease at a given point in time. To this end, all notes for an additional 863 patients (403 for derivation, 156 for validation and 304 for test) were used to label past history at a given timepoint, and were used in combination with the note-level past-history model to train a patient level past-history model **(Figure S2)**.

**Figure 1.**
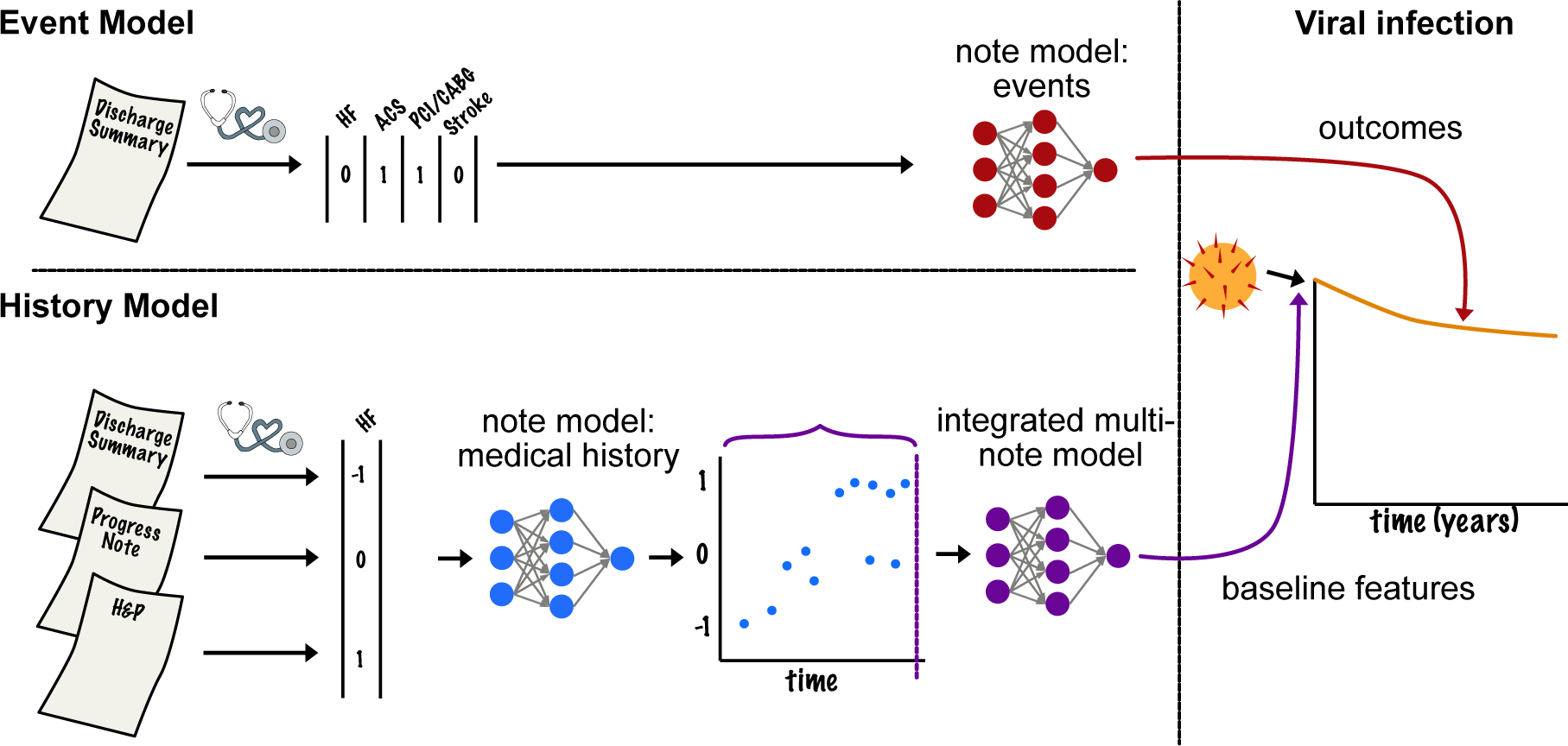
Schematic illustration of the study flow. A large compendium of cardiologist-labeled discharge summaries and progress notes were used to train four cardiovascular event adjudication models and a heart failure history model using neural networks. The models were deployed on 17 million notes for 267,596 patients to evaluate the impact of respiratory virus infections on delayed cardiovascular events.

**Table 1.** Sensitivity and specificity of ICD-10 code.

| Model | Sensitivity | Specificity |
| --- | --- | --- |
| PCI/CABG | 0.30 | 1.00 |
| ACS | 0.78 | 0.95 |
| Heart failure hospitalization | 0.68 | 0.95 |
| Ischemic stroke | 0.95 | 0.93 |

### Neural network-based models have high accuracy for event adjudication

We trained four event adjudication models based on a neural network architecture using the labeled discharge summaries as training and testing data **(Figure S3)**. The neural network-based classifiers had excellent accuracy for detecting events with area under the curve of receiver operating characteristics (AUC-ROC) of 0.97 (95%CI 0.95-0.98) for heart failure hospitalization, 1.0 (0.99-1.00) for PCI/CABG, 0.97 (0.95-0.98) for ACS and 0.98 (0.97-0.99) for ischemic stroke (**Figure 2**). In comparison, structured diagnostic data (ICD-10 codes for ACS/HF/IS and Current Procedural Terminology (CPT) codes for PCI/CABG) had excellent specificity but only modest sensitivity with the exception of ischemic stroke, for which diagnostic codes had excellent sensitivity for the notes we examined **(Table 1)**. The good performance of the ICD-10 codes for stroke may be attributable to the fact that most of the stroke cases we considered were admitted to neurology intensive care units, which may not be the case at other institutions/settings. Overall, our models were able to provide increased sensitivity at a 90% specificity threshold for events **(Table 2).** Based on these results, we predicted events based on our event adjudication models for HF hospitalization, PCI/CABG and ACS and used ICD-10 codes to define ischemic stroke events.

**Figure 2.**
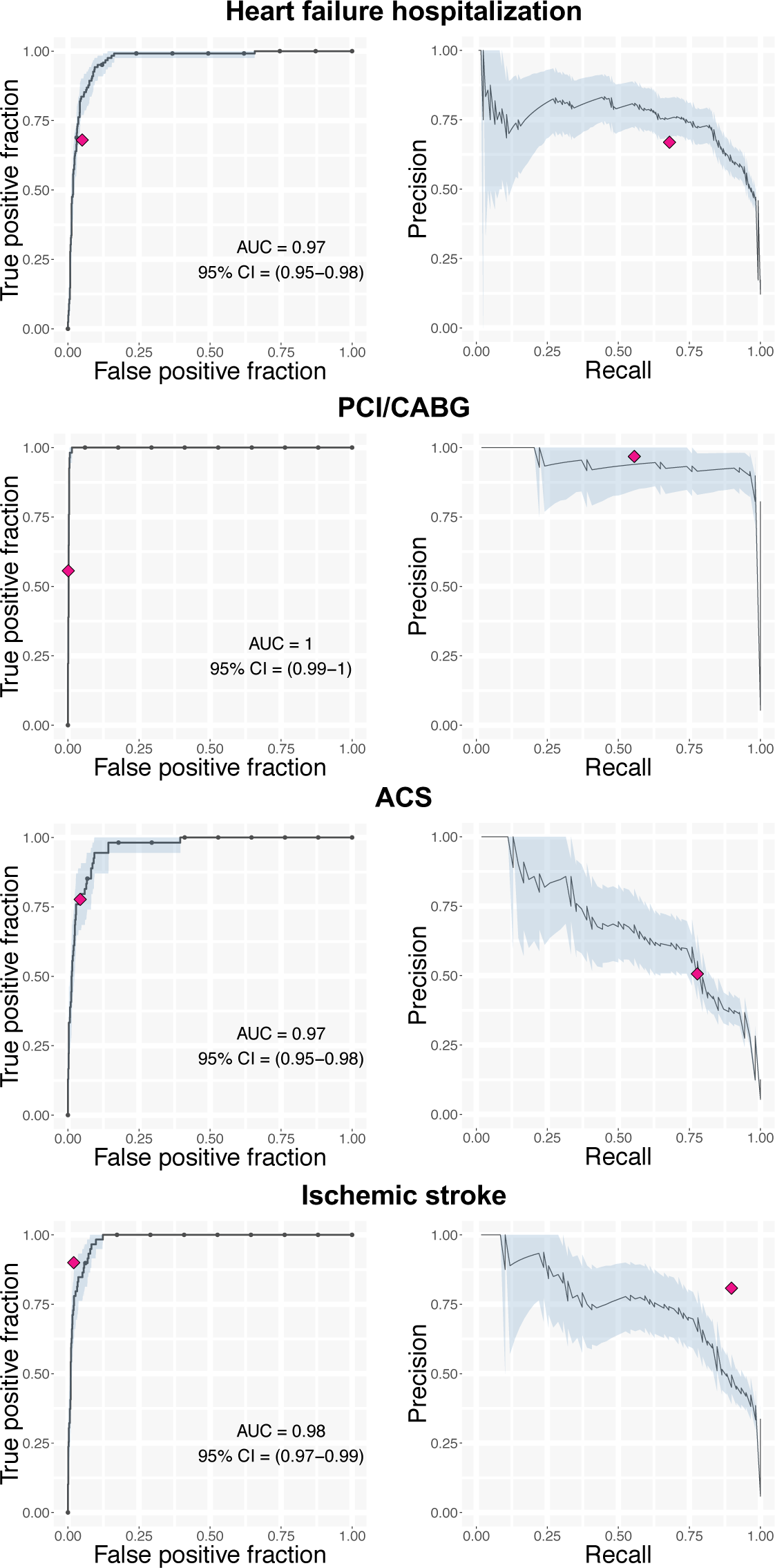
Accuracy of event adjudication model to classify hospitalized events. Receiver operating characteristics curves (left) and precision-recall curves (right) for event adjudication models for PCI/CABG, ACS, heart failure hospitalization and ischemic stroke in the test dataset. Light blue area indicates 95% confidence interval and the pink diamond indicates the sensitivity and specificities of the ICD-10 code-based diagnosis.

**Table 2.** Cutoffs, sensitivity and specificity of the event adjudication model.

| Model | Cutoff used for survival analysis | Sensitivity (95% CI) | Specificity (95% CI) |
| --- | --- | --- | --- |
| PCI/CABG | 0.73 | 1.00 (1.00-1.00) | 0.99 (0.98-1.00) |
| ACS | 0.04 | 0.91 (0.83-0.98) | 0.91 (0.89-0.93) |
| Heart failure hospitalization | 0.12 | 0.94 (0.90-0.98) | 0.91 (0.89-0.93) |
| Ischemic stroke | 0.59 | 0.98 (0.95-1.00) | 0.90 (0.88-0.92) |

### Patients positive for respiratory syncytial virus infection have a higher risk of subsequent heart failure hospitalization compared with those with negative results

A scalable method of accurate event adjudication enables a breadth of downstream applications of clinical and economic value to hospital systems and public health measures. We focused on a timely area, the association of respiratory virus infection with remote cardiovascular events. Using data from the MGB EDW, we identified 267,596 patients with diagnostic test results for a respiratory virus. Of these, we first focused on the 57,828 patients with respiratory syncytial virus (RSV) diagnostic test results, out of which 35,056, 35,828 and 35,778 patients were included in the final cohort for predicting heart failure hospitalization, urgent revascularization and ischemic stroke analysis respectively **(Figure S4-S6)**.

By selecting patients based on having undergone a viral test, we introduced possible selection (collider) bias^14^, given that clinical suspicion for a respiratory virus implies an increased likelihood of a cardiopulmonary etiology **(Figure S7)**. We thus used propensity-score matching to identify patients with a negative test result but with a similar probability of having a positive result for viral infection, as judged by values of other clinical (confounding) variables^15^. Propensity-score matching with 1:16 ratio resulted in well-balanced baseline characteristics between RSV positive and negative patients for all 3 outcomes (standardized mean difference <0.1 for all characteristics, **Tables S3-S8**). To focus on delayed outcomes rather than acute outcomes, we defined events as occurring at least 30 days after the test result was obtained (and matched patients by comorbidities up to that point). RSV positive patients had an event rate of 10.5/100 person-years of heart failure hospitalization which was significantly higher compared to those with negative results who had an event rate of 8.6/100 person-years (n=26,027, median follow-up 368 days, p = 0.0032, **Figure 3 upper panel**). There was no significant difference between RSV positive patients and negative patients for urgent revascularization, defined as the conjunction of a diagnosis of acute coronary syndrome and either PCI or CABG (n=26,724, median follow-up 388 days, 0.8/100 person-years for test positive patients and 0.7/100 person-years for test negative patients, p = 0.49) and ischemic stroke (n=26,690, median follow-up 389 days, 1.1/100 person-years for test positive and 0.9/100 person-years for test negative, p = 0.46). To evaluate if this analysis was driven mostly by early events, we restricted events to those occurring at least 90 days after the test result (patient selection and baseline characteristics shown in **Figures S8-S10** and **Tables S9-14**). This sensitivity analysis showed similar results with a longer median follow-up period of 418, 444 and 442 days for heart failure hospitalization, urgent revascularization and ischemic stroke, respectively, suggesting that our results are driven by delayed events rather than acute events **(Figure S11 upper panel)**.

**Figure 3.**
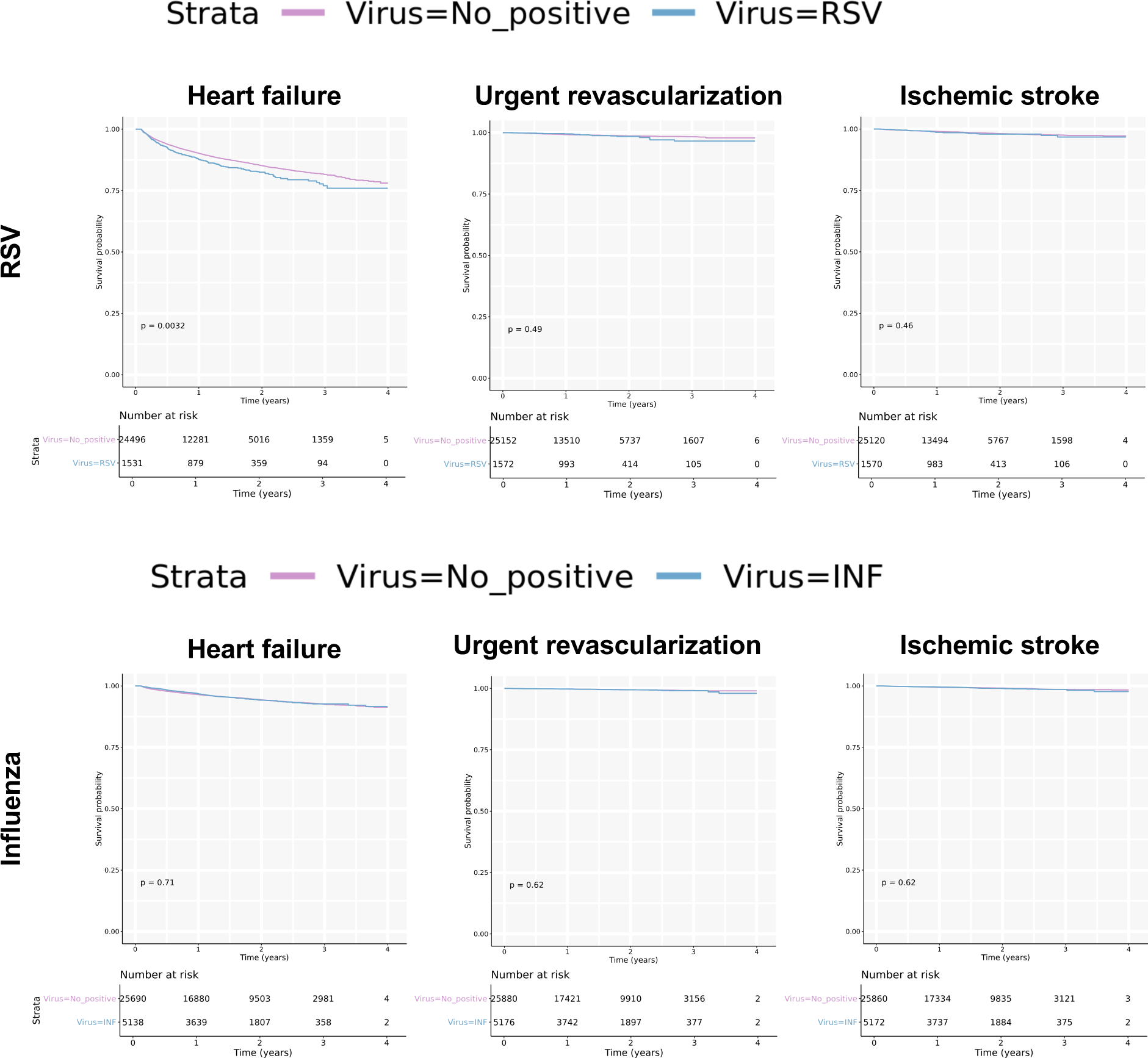
Survival analysis for RSV and influenza infection on cardiovascular events. Kaplan-Meier plot comparing event rates of heart failure hospitalization, urgent revascularization and ischemic stroke in virus positive and negative population for RSV and influenza. Abbreviations, RSV: respiratory syncytial virus, INF: influenza virus.

### Patients positive for influenza virus infection do not have an increased risk of delayed cardiovascular events compared to those with negative results

We identified 81,250 patients with influenza test results, of which 58,522, 59,594 and 59,523 patients were included in the final cohort for heart failure hospitalization, urgent revascularization, and ischemic stroke, respectively **(Figure S12-S14).** As for RSV, we used propensity-score matching to adjust for differences in baseline characteristics between the virus positive and negative population. This matching procedure resulted in 1:5 matching between influenza positive and negative patients for all 3 outcomes (standardized mean difference <0.1, **Tables S15-S20**). In contrast to the RSV virus, there were no significant differences between influenza positive patients and negative patients for delayed heart failure hospitalization (n=30,828, median follow-up 614 days, 2.8/100 person-years for test positive and 2.9/100 person-years for test negative, p = 0.71), urgent revascularization (n=31,056, median follow-up 643 days, 0.3/100 person-years for test positive and 0.3/100 person-years for test negative, p = 0.62) or ischemic stroke (n=31,032, median follow-up 653 days, 0.5/100 person-years for test positive and 0.5/100 person-years for test negative, p = 0.62, **Figure 3 lower panel**). As with RSV, we also performed an analysis by using 90 days as the early event exclusion period (patient selection and baselines shown in **Figures S15-S17** and **Tables 21-26**). The results were similar to the main analysis with median follow up period of 670, 682 and 653 days for heart failure hospitalization, urgent revascularization and ischemic stroke, respectively **(Figure S11 lower panel).**

### Sensitivity analysis using a neural network-based model for heart failure history shows robustness of results

The past medical history of heart failure used for propensity-score matching was derived from an aggregate use of ICD-10 codes from problem lists, encounter diagnoses, medical history, and admission diagnoses. Given concern for inaccuracy of structured data, our finding that RSV infection was associated with higher rate of heart failure hospitalization may have been affected by the definition of HF history (which has large impact on this outcome). To overcome this concern, we first sought to evaluate the accuracy of the ICD-10 code-based approach to define HF history. To this end, we performed a systematic effort of labeling 863 patients and found that the ICD-10 approach nonetheless had a good accuracy with sensitivity 0.80 and specificity 0.97 (**Figure 4A**). Although the performance was good, the use of structured data typically leaves no flexibility to explore robustness of the result to different thresholds. We were particularly concerned about the impact of a reduced sensitivity. Therefore, we developed a neural network-based approach for learning past medical history at a given point in time using a neural network-based strategy. We trained three separate models: a model for note-informativeness, a model for heart failure history classification, and finally a time series model that used an aggregate of note scores across time. The final model had an excellent discriminative ability with ROC-AUC of 0.97 (**Figure 4A**). Although the accuracy at the mandatory point of comparison was not superior to the ICD-10 code-based definition, the model enabled evaluating robustness of the results to the selected cutoff point. We assessed conclusions using cutoffs with high sensitivity, high specificity and a balanced sensitivity/specificity ratio. Different cutoffs gave different absolute rates of heart failure history **(Table S26)**. Nonetheless, propensity score matching using model-based heart failure history was successful with SMD<0.1 for all baseline values **(Table S27-32)** and the overall results were unchanged for the association of heart failure hospitalization with viral infection, with p-values ranging from 0.0030 to 0.0043 for RSV and 0.28 to 0.70 for influenza (**Figure 4B**).

**Figure 4.**
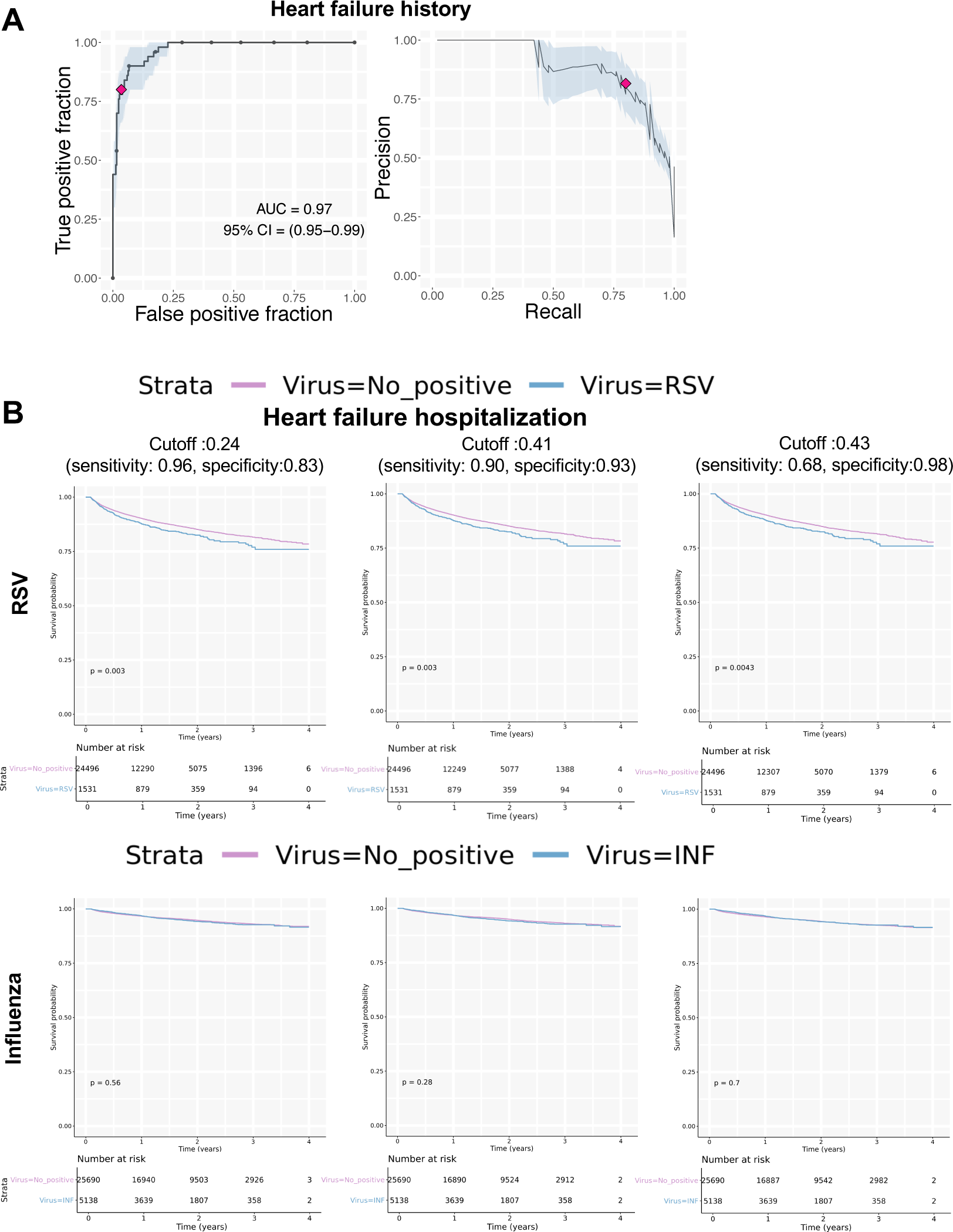
Sensitivity analysis evaluating different thresholds for a past medical history of heart failure on the risk of heart failure hospitalization with viral infection. (A) Receiver operating characteristics curves for the heart failure past medical history model. (B) Kaplan-Meier plot comparing event rates of heart failure hospitalization in virus positive and negative populations for RSV and influenza with various cutoffs for heart failure history. Sensitivity and specificity for detecting heart failure histories with the corresponding cutoffs are shown. Abbreviations, RSV: respiratory syncytial virus, INF: influenza virus.

## Discussion

Here we outline two primary contributions; 1) machine learning models for automated event adjudication for cardiovascular events and past medical history and 2) application of these models to estimate the risk of delayed heart failure hospitalizations in individuals with respiratory virus infections. In this context, we observed an estimated 23% higher risk of delayed heart failure hospitalizations in individuals infected with RSV compared to those who tested negative but no change in the risk for such events following influenza infection.

Our work labeling a large collection of notes across over 17 institutions within an integrated delivery network adds further support to the limitations of diagnostic codes for describing cardiovascular events^2^. The accuracy of diagnostic codes varied strongly with the type of event: at one extreme, ischemic stroke was very well captured whereas there were no clear diagnostic codes for coronary revascularization, and, at least within our database, the use of CPT codes had very low sensitivity. Reimbursement incentives are known to shape the completeness of diagnostic codes^16^ and there is also further variation from center to center based on provider and institutional coding practices. Nonetheless, a systematic labeling approach can provide local estimates of the adequacy of diagnostic code for any specific application, and further simulation-based modeling can be used to understand the impact of misclassification bias on any downstream uses of the data.

Although we found that a historical compilation encompassing encounter diagnoses, admission diagnoses, problem lists, and medical history is highly accurate, at least for heart failure, the sensitivity was relatively low compared to specificity, which is in agreement with previous studies showing that ICD-10 codes for past medical history suffers primarily from problems of sensitivity^17^. The ICD-10 code-based approach forces a single sensitivity/specificity and does not allow adjustment or optimization. In comparison, our model-based definition can provide various sensitivity/specificity ratio by adjusting the threshold. This feature enabled us to perform a sensitivity analysis showing that our results are robust to the sensitivity/specificity of the past medical history detection. Furthermore, we anticipate that our approach should generalize well to capturing conditions for which there are no diagnostic codes (such as inadequate response to a medication).

In terms of prior work, most of the focus on EHR-based automated diagnoses has been on the use of heuristic combinations of diagnostic codes, medications, labs, and regular expression matching within unstructured data^2^. Recent advances in convolutional neural network-based text processing have motivated their application to the problem of note classification^18, 19^. Our own experience with this process emphasized the need for both domain (i.e., cardiovascular disease) and data science expertise, with an iterative examination of failure cases, augmented labeling, and retraining of models.

Our models enabled scalable analysis of millions of discharge summaries and progress notes. We see multiple applications of such an approach, whether it be for pharmacovigilance, quality control, or patient selection for a clinical trial according to criteria that may be poorly reflected in structured data. For our specific application, we were able to conclude that influenza virus infection does not appear to contribute to any delayed increase in heart failure hospitalization, while RSV infection contributes a 23% increase in risk that is apparent across an entire four-year period.

Prior work focusing on respiratory viruses have emphasized the acute cardiovascular manifestations of viral infections in a limited subset of patients^5, 20^. In the case of RSV, it is well recognized that patients with RSV infections have a higher burden of cardiovascular disease, although this may reflect selection bias, given that cardiovascular consequences of viral infection may add to pre-existing disease to drive presentation to healthcare systems. However, our results are striking in that even controlling for heart failure diagnoses prior to and up to 90 days after the incident event, we still see an increased risk of future events. While previous reports show increased risk of cardiovascular events in patients with influenza infections, they focus entirely on acute events in the context of the infection^6–8^. Our analysis excludes acute events and thus, does not contradict these findings. It is entirely plausible that influenza virus increases heart failure hospitalization in the acute phase, either through direct myocardial involvement^21^ or indirectly through a systemic inflammatory response as with many other infections^22^, but patients may not have prolonged damage to the heart.

The primary limitation of our work is the unknown contribution of unmeasured confounders with significant impact on both heart failure hospitalization outcomes and probability of positive test for viral infection. Since our analysis focused only on those who had been tested for the virus, these population likely had symptoms suggestive of a cardiopulmonary illness. Thus, the negative results for the virus results may be consistent with other reasons for having respiratory symptoms. In the case of influenza virus, we observed a classic example of collider bias requiring propensity score matching approaches^23^. Nonetheless, such approaches inevitably can suffer from model misspecification. However, it should be noted that possible approaches to address these limitations such as randomizing patients to be infected by a virus or performing a cohort study testing every individual regardless of the symptoms are not feasible.

An open question is whether our work has relevance to other respiratory viruses, such as SARS- COV2. Although SARS-COV2, like influenza^21^ and RSV, can cause myocarditis^9–11, 24^, there is no clarity on the prevalence of this effect given the selection bias in published reports. Nonetheless, it is reasonable to assume the possibility of long-term cardiac consequences for SARS-COV2, that will only be addressed with multi-year surveillance and methodologic approaches such as our own.

## Methods

### Ethics statement

Institutional review board approval was obtained for all aspects of this study,

### Selection and labeling of clinical notes for event model training and evaluation

The Massachusetts General Brigham (MGB) Electronic Data Warehouse (EDW), which includes notes from >20 provider locations across a large integrated delivery network, was the source of notes and structured clinical data used in this study. Two sets of discharge summaries were used for model training and testing. The training set, consisting of 1964 discharge summaries, were partially enriched with the use of ICD10 codes for stroke, myocardial infarction, and heart failure. The test set, consisting of 1,003 discharge summaries, was selected randomly from patients above who had undergone electrocardiograms within the centers participating in the EDW.

Following previously published guideline^13^, one of 4 cardiologists labeled each discharge summary for 4 possible diagnoses: acute coronary syndrome, heart failure as the primary cause of hospitalization, percutaneous coronary intervention or coronary artery bypass surgery (PCI/CABG), or ischemic stroke.

### Processing of clinical notes

We used SpaCy v2.2.4^25^ to tokenize medical notes. We extended the English language model (en-core-web-large) tokenization rules by adding a comprehensive set of abbreviations that are commonly used in medical records. We used fasttext v0.9.2^26^ to train 100-dimensional bigram word embeddings with the parameters ‘fasttext skipgram -wordNgrams 2’. Word embeddings were trained on a compendium of more than 10 million clinical notes ranging from 2002 to 2020, based on more than 330,000 subjects, with up to 50 notes per subject.

### Training of event adjudication models

The event adjudication model was constructed with the combination of 1D-CNN and long-short-term memory (LSTM). It consisted of a word embedding layer, which converts words into word vectors, followed by a 1D-CNN and LSTM combination network (schematic shown in **Figure S1**). The model was trained using data from derivation dataset. Each discharge summary was labeled as positive or negative for 4 outcomes of interest (heart failure hospitalization, PCI/CABG, ACS and ischemic stroke) and 4 separate models were trained for each outcome label. The model was trained to minimize the binary cross entropy between model prediction and the label using RMSprop optimizer with initial learning rate of 0.0001. The model was trained for 150 epochs. At the end of each epoch, ROC-AUC on the validation dataset was calculated. The final model was chosen as the model with highest ROC-AUC on the validation cohort across all 150 epochs.

### Selection of test results for respiratory virus infection

Using a data dictionary, we identified identifiers for diagnostic tests for influenza A and B, parainfluenza, respiratory syncytial virus (RSV), coronavirus, and adenovirus and downloaded all relevant viral test results within the MGB-EDW. We limited analysis to tests having at least 5000 patients. The test results were reported as free text. While most results were reported simply as positive or negative, in some cases, the descriptions of the viral test results were complex. Therefore, a physician (SG) reviewed all possible texts in the report and compiled a conversion table from the texts to positive/negative results. Since comprehensive reporting in the EDW started on Feb 2016, we included only results after that date.

### Survival analysis for cardiovascular events

To evaluate the influence of viral infection on 3 cardiovascular events (heart failure hospitalization, urgent revascularization and ischemic stroke), we performed a survival analysis for these outcomes. Due to limited numbers or follow-up length for other viruses, we focused our analysis on influenza and RSV. To avoid possible bias caused by the COVID-19 outbreak in Boston area, all the follow up were censored at February 2020. All viral test results between February 2016 and February 2020 were identified. Positive patients were identified as those having at least one positive result for the virus of interest within the study period and the date of first positive result was used as the index date. Patients who had the test for the virus of interest but did not have any positive results were identified as a control population within that cohort. Index dates were identified as the date of first viral test for control patients. All patients who had other viral infections before the index date were excluded.

Outcome events were defined bases on the event text model predictions for heart failure hospitalization and urgent revascularization and ICD-10 codes for ischemic stroke. The cutoffs for the events were selected as the value that gave the best sensitivity at a specificity over 90%. Urgent revascularization was defined as positive results for PCI/CABG and ACS for the same discharge summary. All 3 outcome events were analyzed independently. Patients were censored at the first event, death, last encounter in the system or the end of study period, whichever occurred first. To account for confounders, we used a propensity-score matching method^15^. A propensity score for a positive test was calculated with age, sex, race, smoking status, history of heart failure, history of coronary artery disease, history of atrial fibrillation, history of type 2 diabetes mellitus, history of COPD, history of hypertension, history of dyslipidemia, use of loop diuretic, use of aspirin, use of P2Y12 inhibitors and the method for viral test using logistic regression. In this analysis, the past histories were identified using ICD-10 codes. The between group difference in baseline characteristics was assessed by the standardized mean difference (SMD) where a value under 0.1 is considered negligible. Patients with positive viral results were matched to controls (without replacement) using a nearest neighbor method, as implemented in the *MatchIt* package (version 3.0.2) in *R* (version 3.5.1)^27^. The matching ratio was set to the highest number that did not introduce substantial difference in any baseline characteristics (SMD<0.1).

### Training and testing of a past medical history model

Since past medical history is defined using a cumulative record and not on a single note, we took a 2-step approach for training the history model. Our first step focused on classifying the note. Since some notes are totally unrelated to the history of disease (such as nutrition notes), we trained a model to classify if the note is informative for a specific history or not and a second model to actually classify if the history is positive or negative. To this end, each note was initially labeled with 3 labels (positive/negative and uninformative). During labeling, we also noticed that there were some notes that stated an ambiguous condition (e.g. extremely low ejection fraction without noting if the patient have symptoms or not for heart failure). We labeled these by a 4th label (ambiguous) and excluded these from the training data. Two sets of notes were used for model training and testing from the MGB-EDW database where the training set were partially enriched with the use of ICD10 codes heart failure and the test set selected randomly. The final numbers of notes were 819 for training set and 365 for the test set. The note-level models resulted in note-level accuracy of AUC 0.93 for informativeness and 0.93 for the presence of heart failure history **(Figure S2A).**

The second step focused on combining all the note-level output to identify the past medical history status for each patient for a specific date. To this end, we built a time-series neural network-model that takes note-level prediction for informativeness and past medical history status along with the dates of the note relative to the index date as input. The model produces a single binary output reflecting the probability of the patient having the disease history at the index date **(Figure S2B)**. For this model, we labeled a separate patient dataset (with no overlap with the note level history model patients) consisting of 561 patient-date pairs for training and 305 patient-date pairs for testing. The patients in the training datasets were partially enriched for past medical history using the event adjudication model to select patients who had an event before the index date. In contrast, the patients in test set were randomly selected from a group of patients 60 years and above who had received an electrocardiogram at MGB. The labeling was done by reading all the notes associated with that patient to identify if the patient had a positive history of the disease at the index date. The input data was constructed as time-series data of the note-level output for all the notes within ± 2500 days of index date. Each output value was inserted into a vector of length 5000 (each element corresponding to the date relative to index date). The neural network was trained to take this vector as input and a binary 1/0 value as output.

### Sensitivity analysis with various thresholds for heart failure history

To evaluate if the results of our survival analysis was sensitive to the sensitivity/specificity of the past medical history detection method, repeated the survival analysis for heart failure hospitalization using the heart failure history defined based on the time-series history model with various cutoff points. We used 3 cutoff points for this analysis, each of which giving (1) highest specificity with sensitivity ≥ 0.95, (2) sensitivity and specificity both over > 0.90, (3) highest sensitivity with specificity ≥ 0.95. The cutoffs were 0.24 (sensitivity=0.96, specificity=0.83), 0.41 (sensitivity=0.90, specificity=0.93) and 0.43 (sensitivity=0.68, specificity=0.98). The propensity score matching was re-done using the model-based heart failure history instead of ICD-10 code-based heart failure history.

## Data Availability

The data that support the findings of this study are available on request from the corresponding author R.C.D. upon approval of the data sharing committees of the respective institutions. The data are not publicly available due to the presence of information that could compromise research participant privacy.

## Supplementary Figures

**Figure S1:**
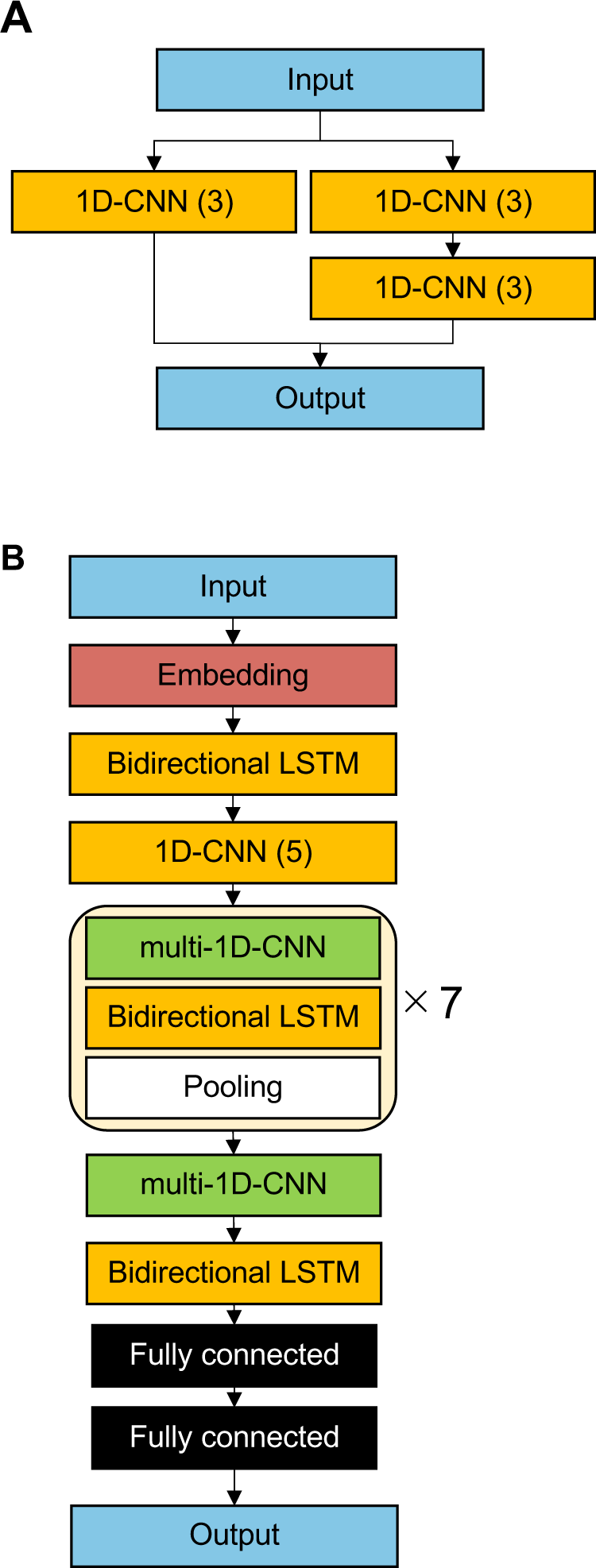
Schematic drawing of the network structure of the event adjudication model. (A) Structure of the multi-1D-CNN module and (B) the full network structure of the model

**Figure S2:**
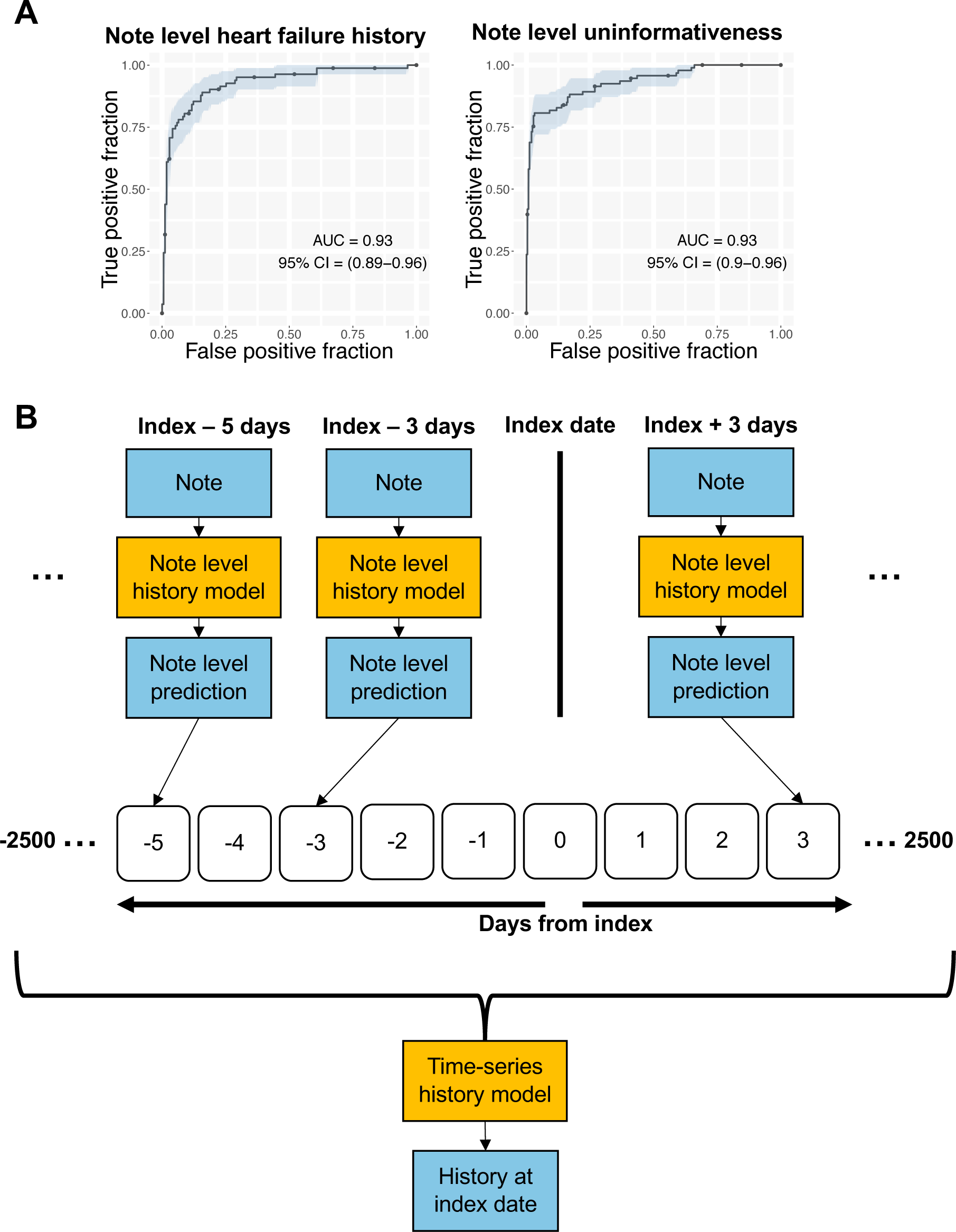
Past medical history model. (A) ROC curves showing performance of note-level past medical history and uninformativeness model. (B) Schematic drawing for time-series model for past medical history.

**Figure S3.**
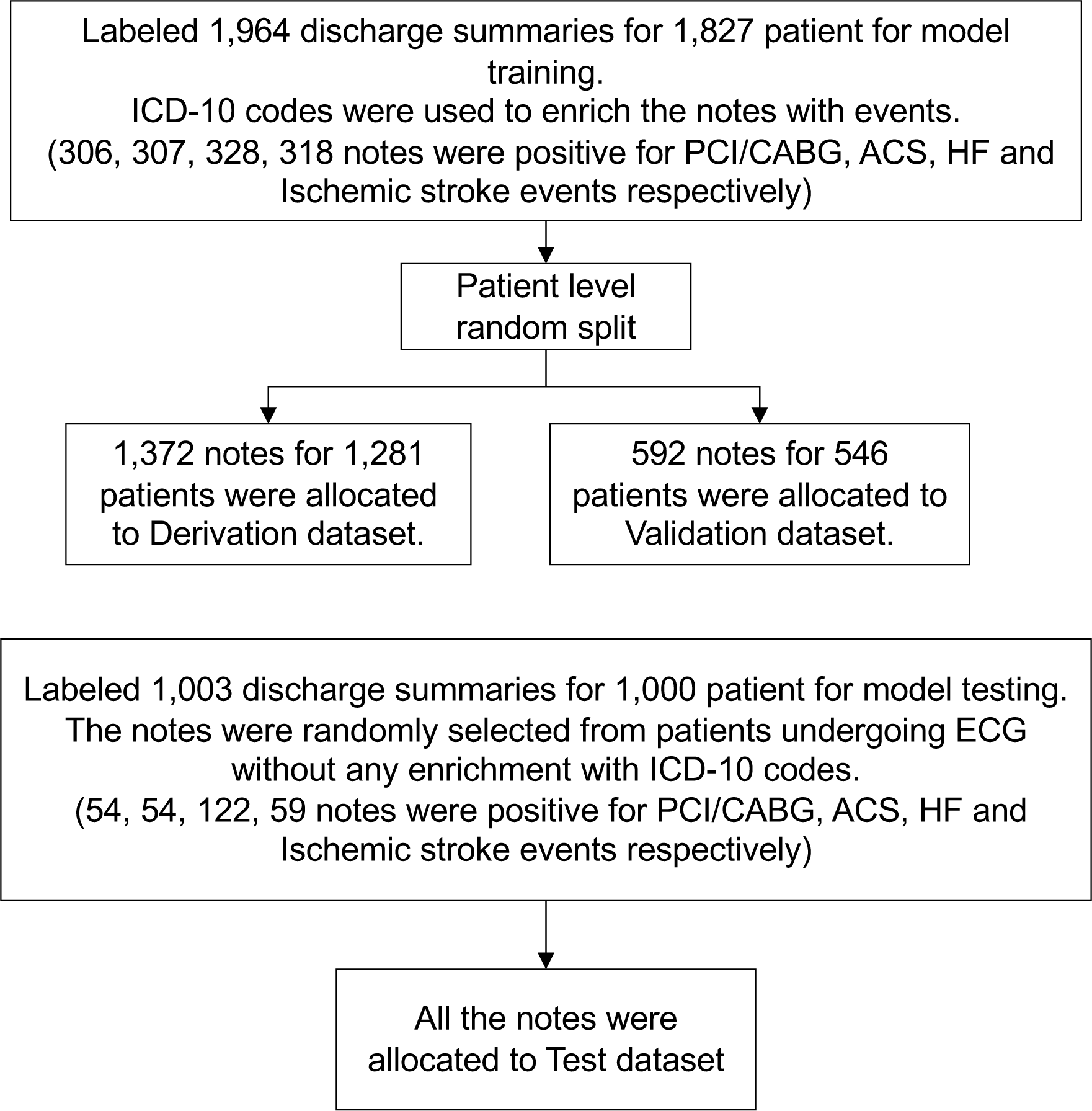
Flow diagram of notes selection used for training and testing event adjudication models.

**Figure S4.**
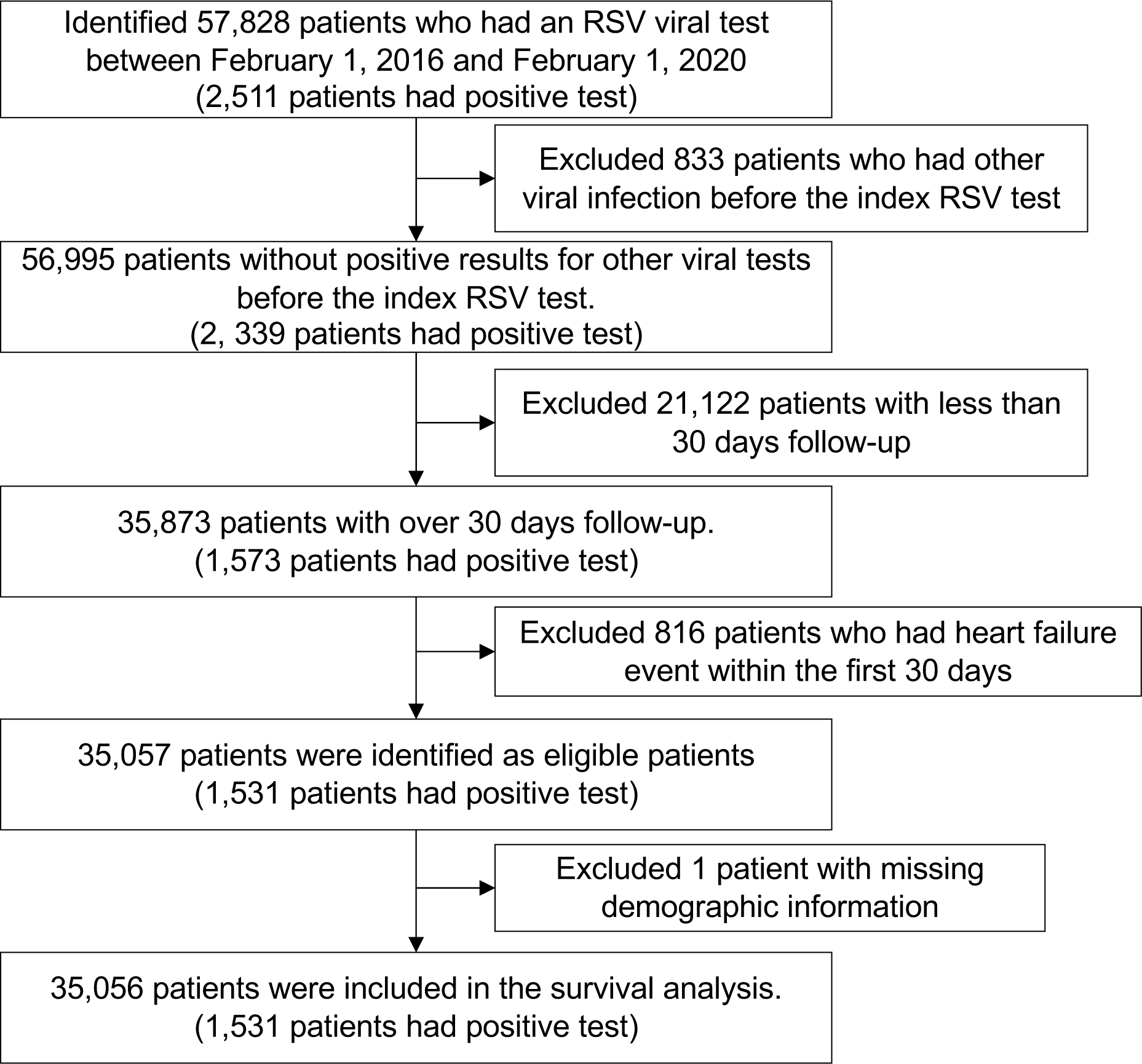
Flow diagram of patient selection for survival analysis for heart failure hospitalization in patients who had an RSV viral test between February 1, 2016 and February 1, 2020.

**Figure S5.**
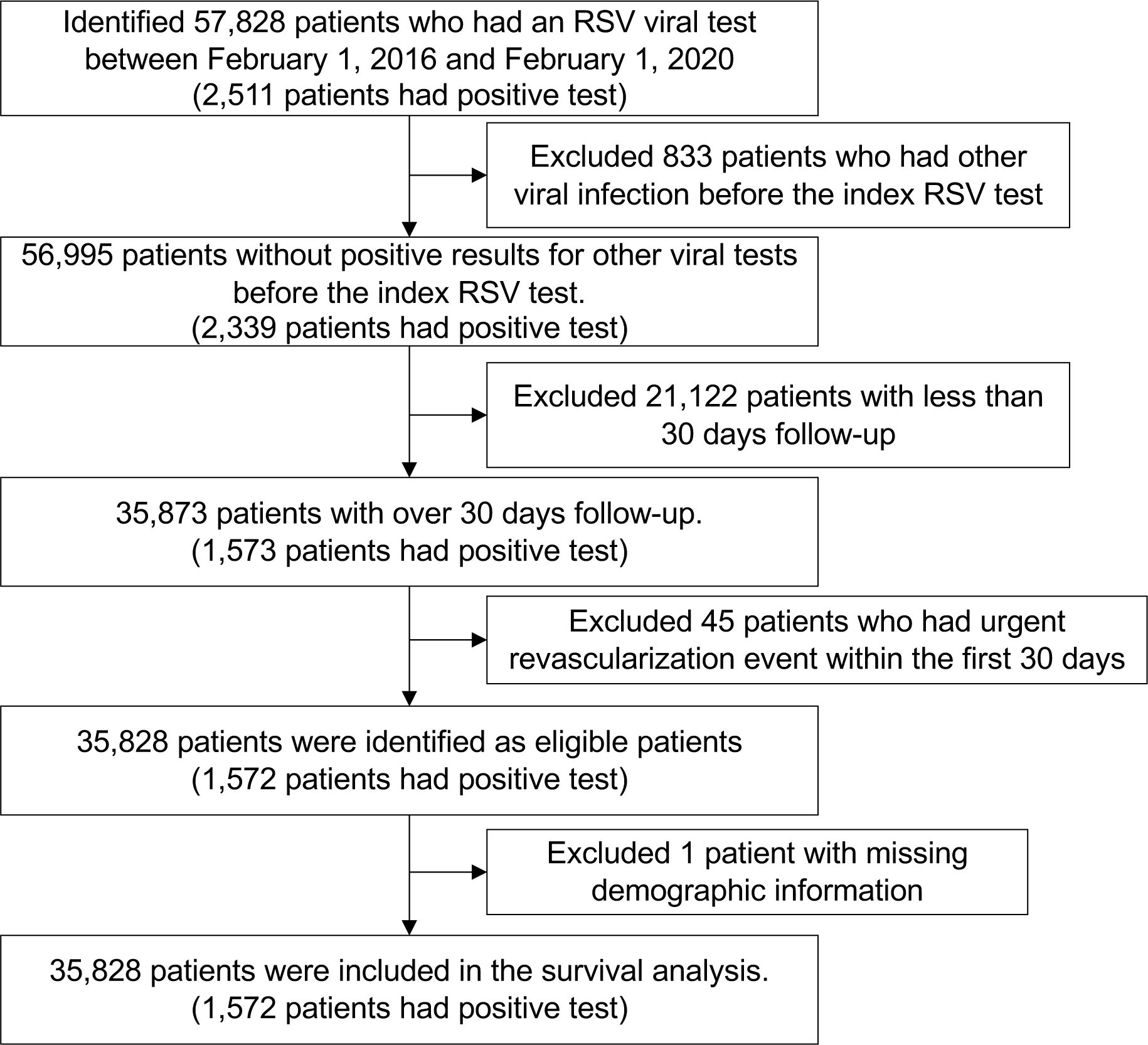
Flow diagram of patient selection for survival analysis on urgent revascularization in patients undergoing RSV viral tests.

**Figure S6.**
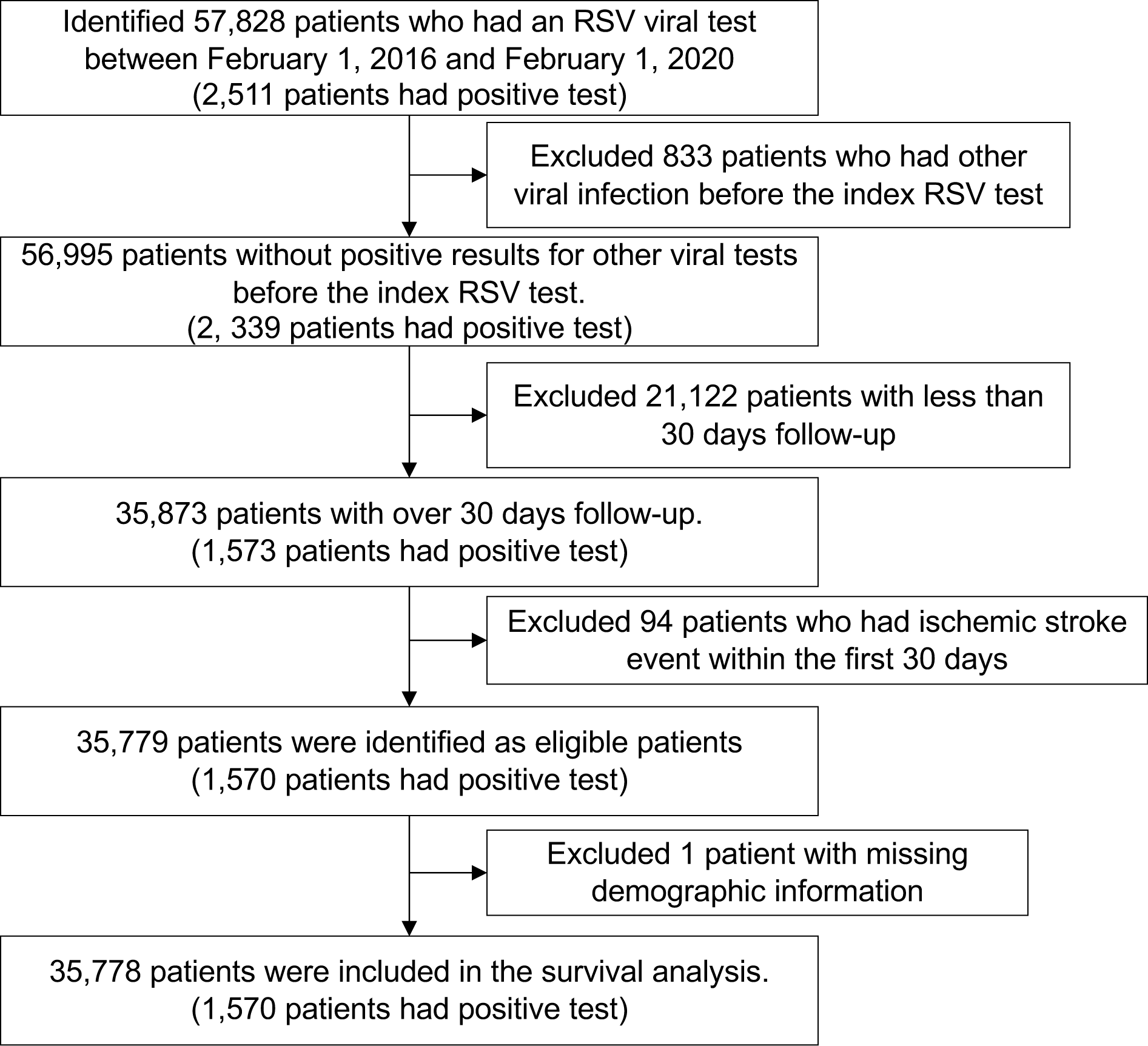
Flow diagram of patient selection for survival analysis on ischemic stroke in atients undergoing RSV viral tests.

**Figure S7.**
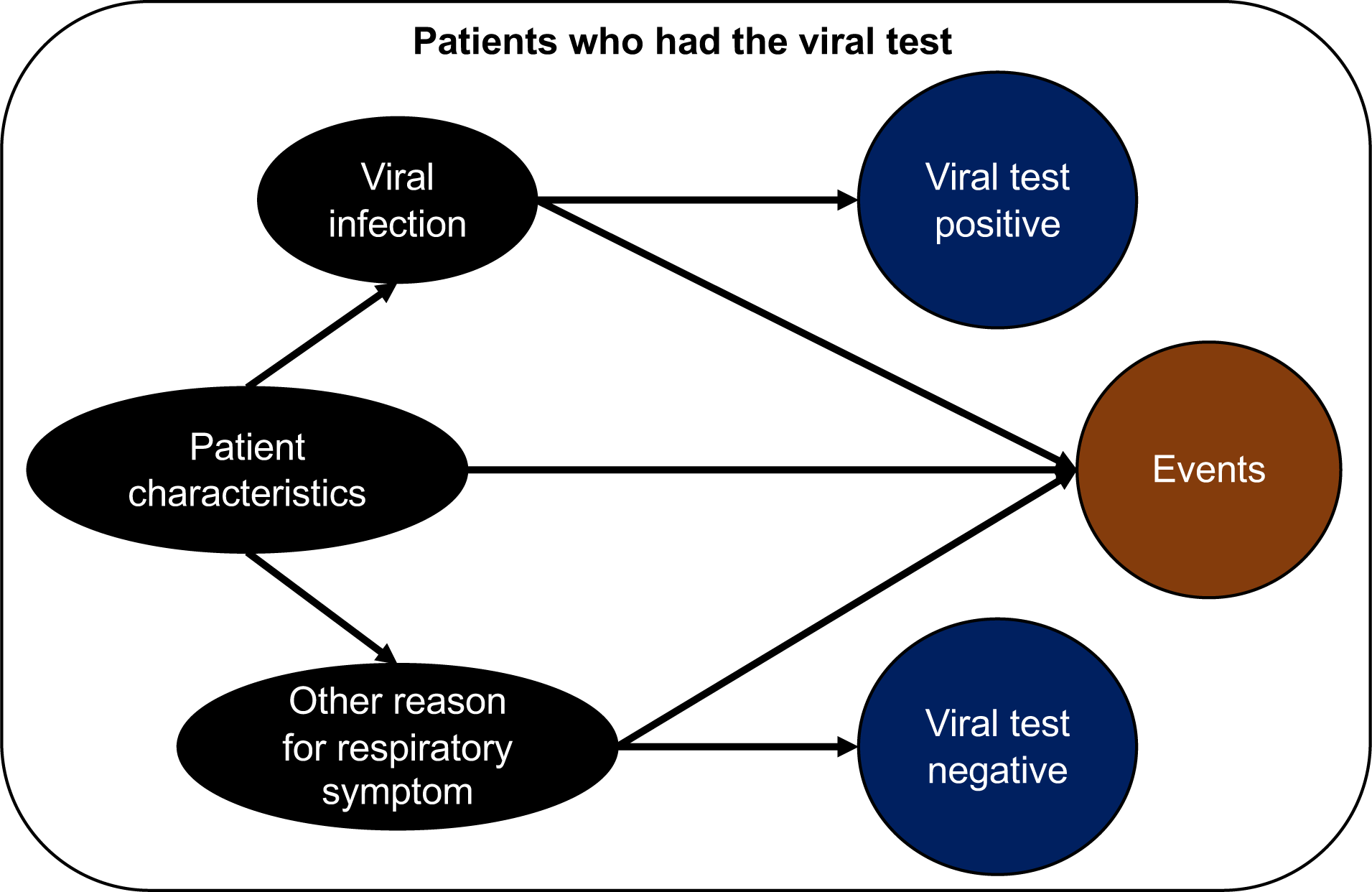
Graphical model illustrating potential selection bias in patients who underwent testing for a respiratory virus. Selecting patients based on having undergone a viral test induces an inverse association between viral infection and other causes of respiratory symptoms (collider bias). The latter may also influence the probability of delayed cardiovascular events, and thus were handled by adjustment.

**Figure S8.**
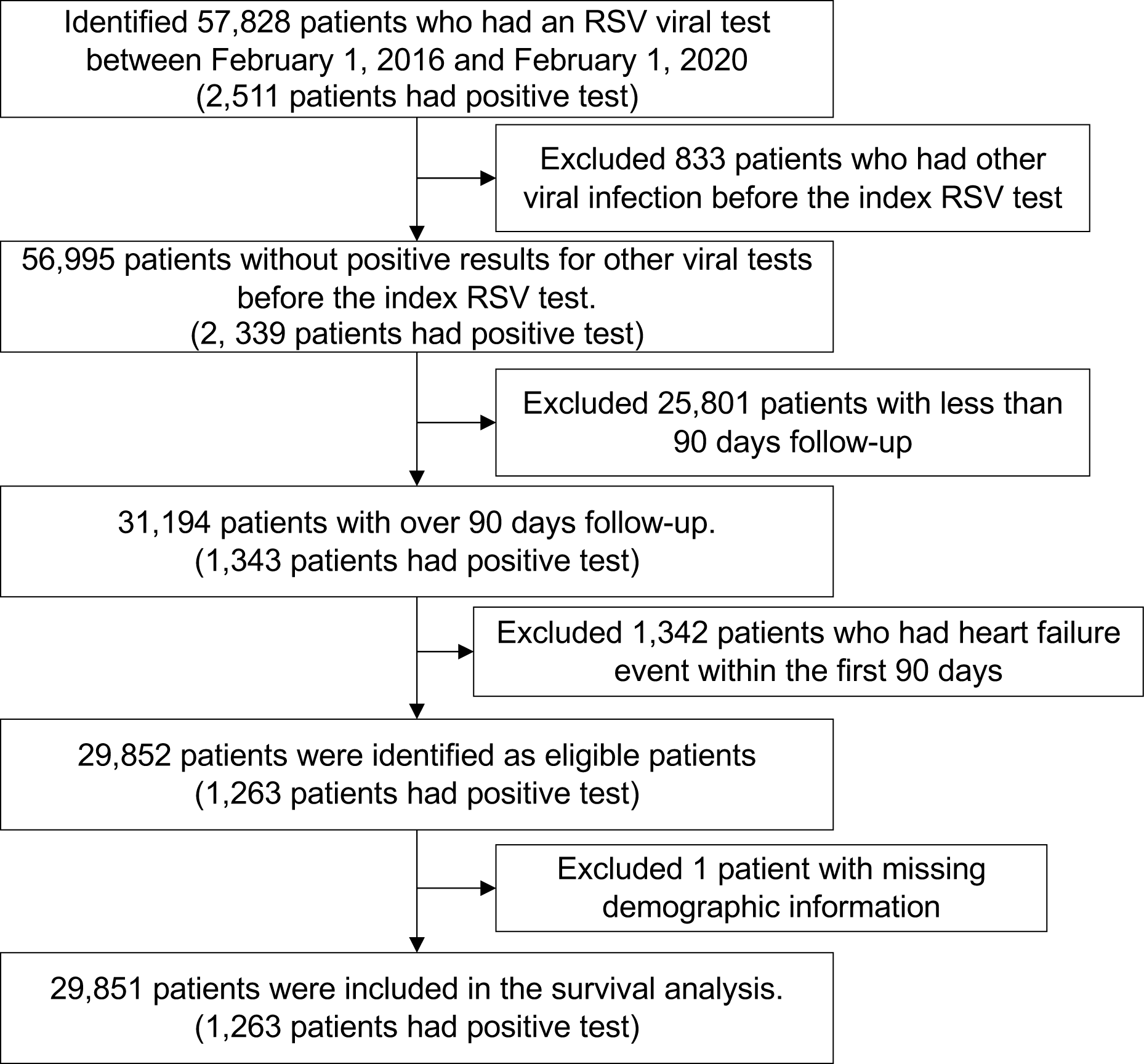
Flow diagram of patient selection for survival analysis on heart failure in patients undergoing RSV viral tests (excluding those who had event within the first 90 days).

**Figure S9.**
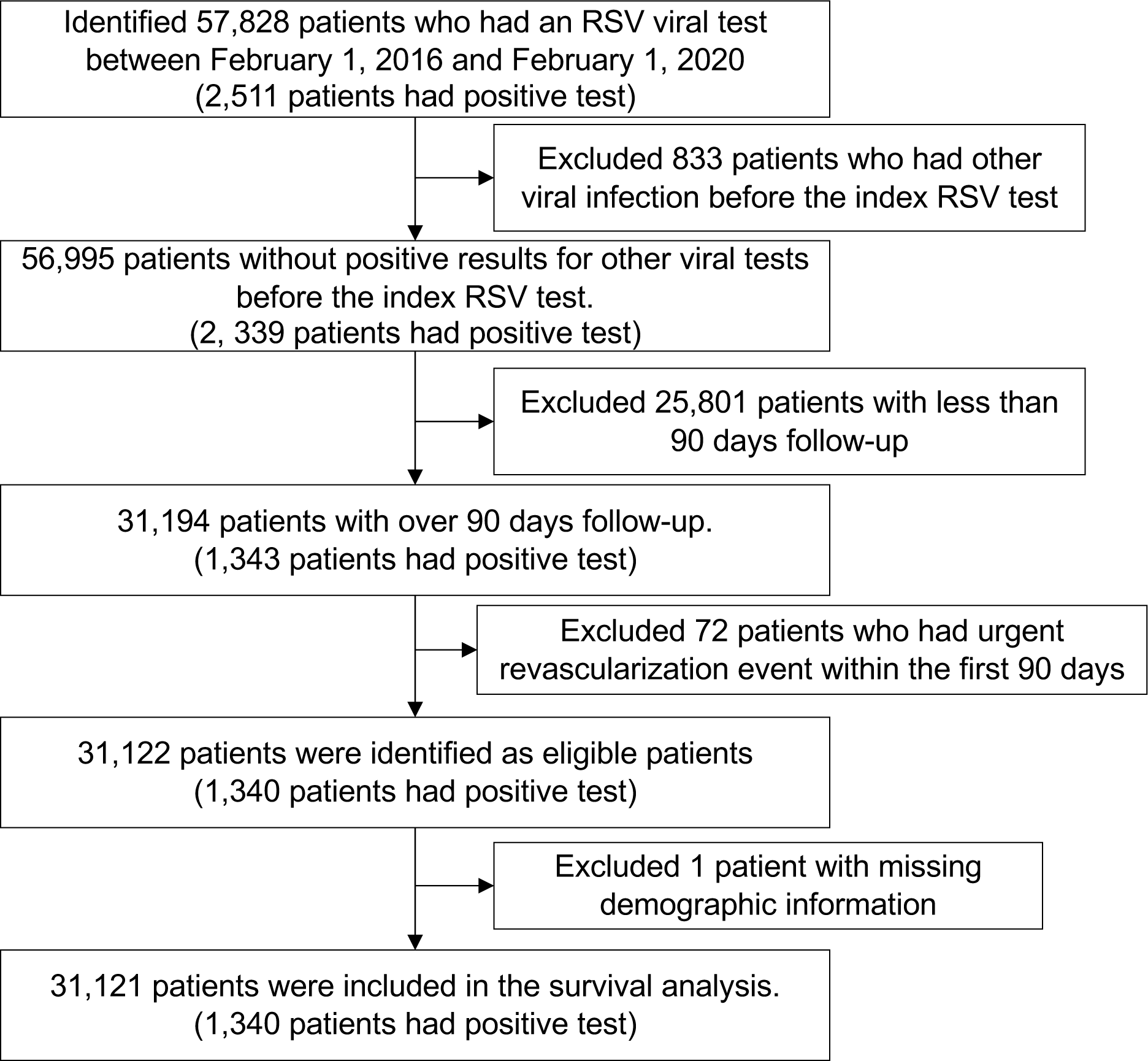
Flow diagram of patient selection for survival analysis on urgent revascularization in patients undergoing RSV viral tests (excluding those who had event within the first 90 days).

**Figure S10.**
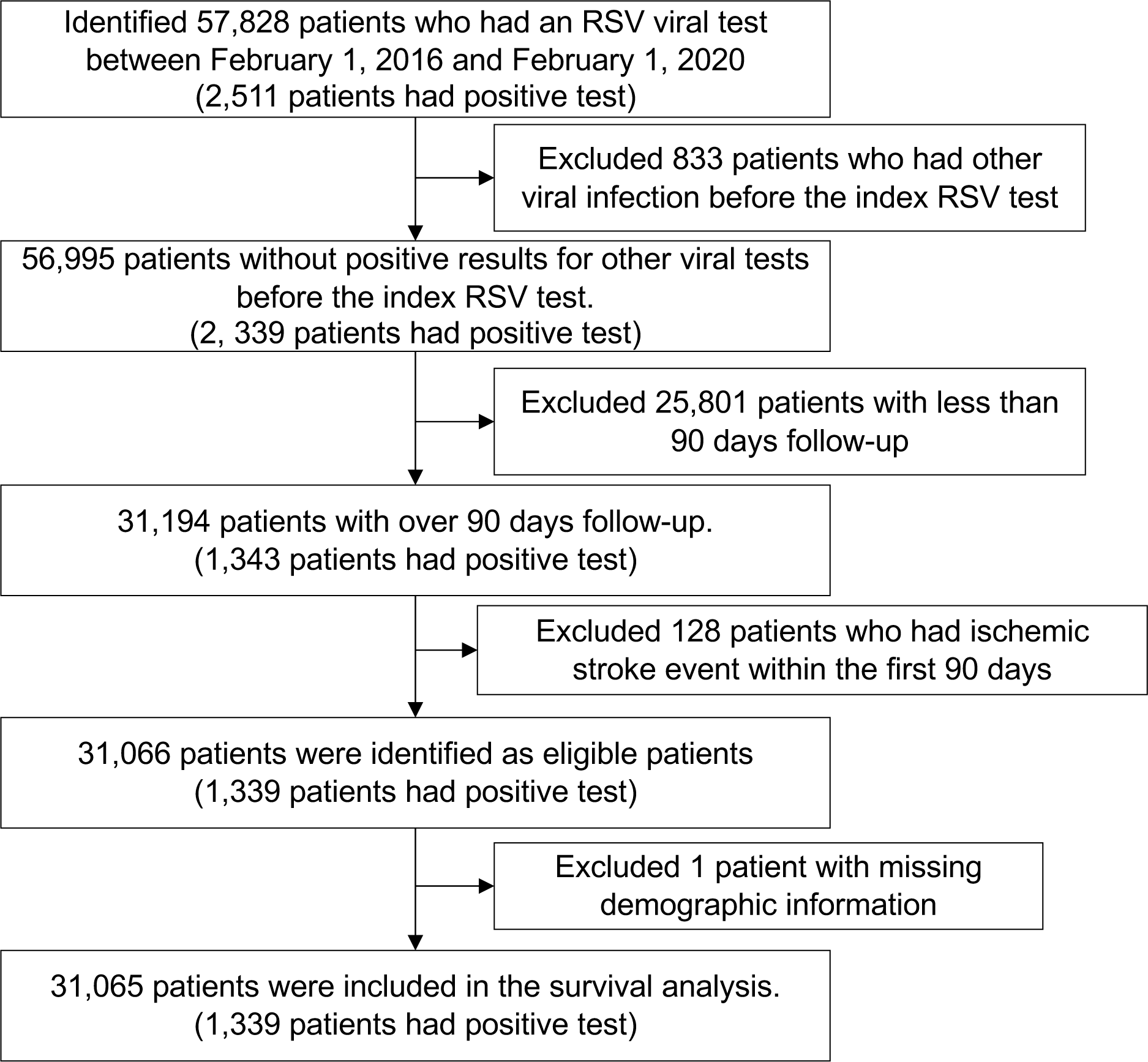
Flow diagram of patient selection for survival analysis on ischemic stroke in patients undergoing RSV viral tests (excluding those who had event within the first 90 days).

**Figure S11.**
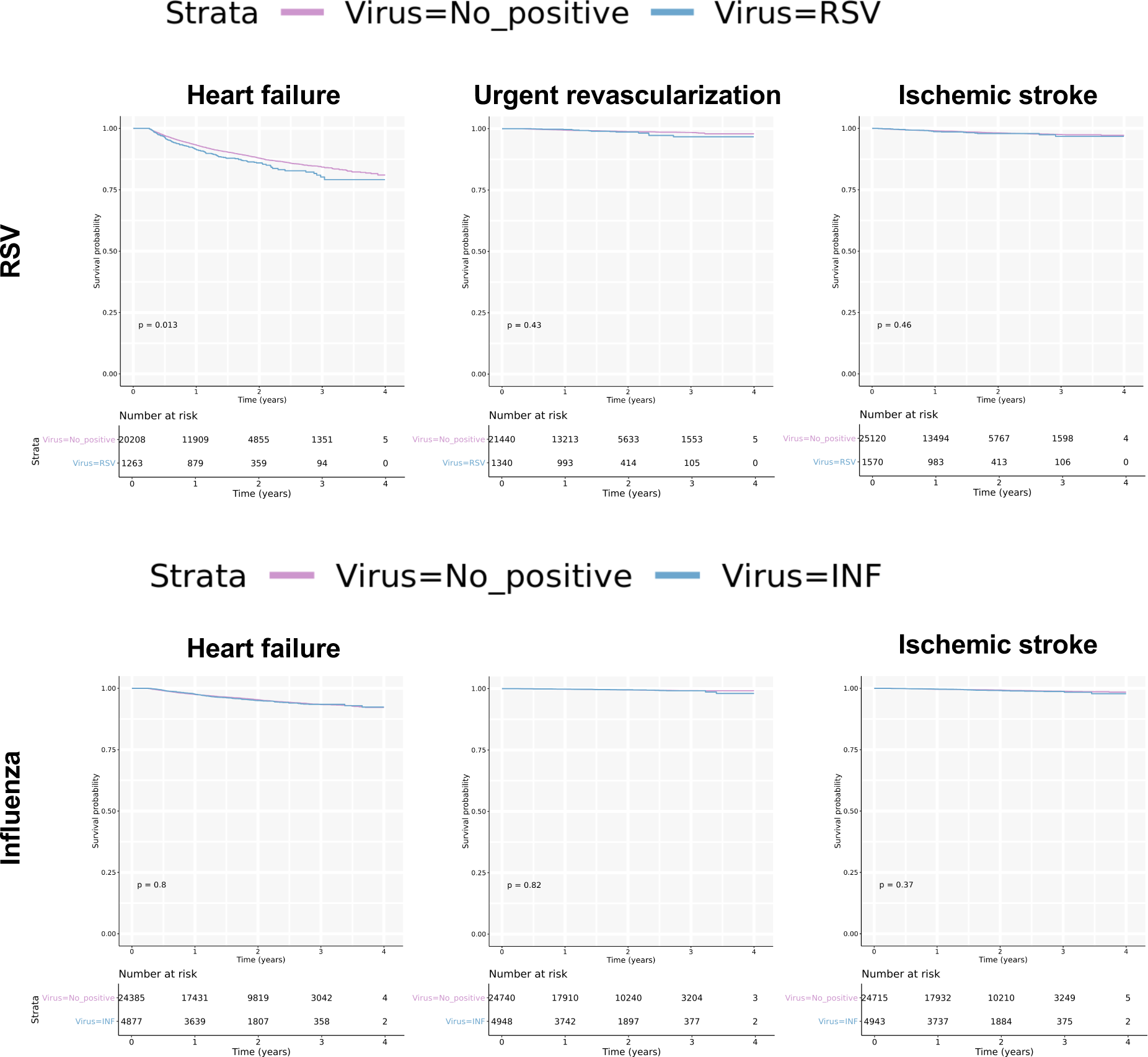
Survival analysis for RSV and influenza infection on cardiovascular events (excluding those who had event within the first 90 days). Kaplan-Meier plot comparing event rates of heart failure hospitalization, urgent revascularization and ischemic stroke in virus positive and negative population for RSV and influenza. Abbreviations, RSV: respiratory syncytial virus, INF: influenza virus.

**Figure S12.**
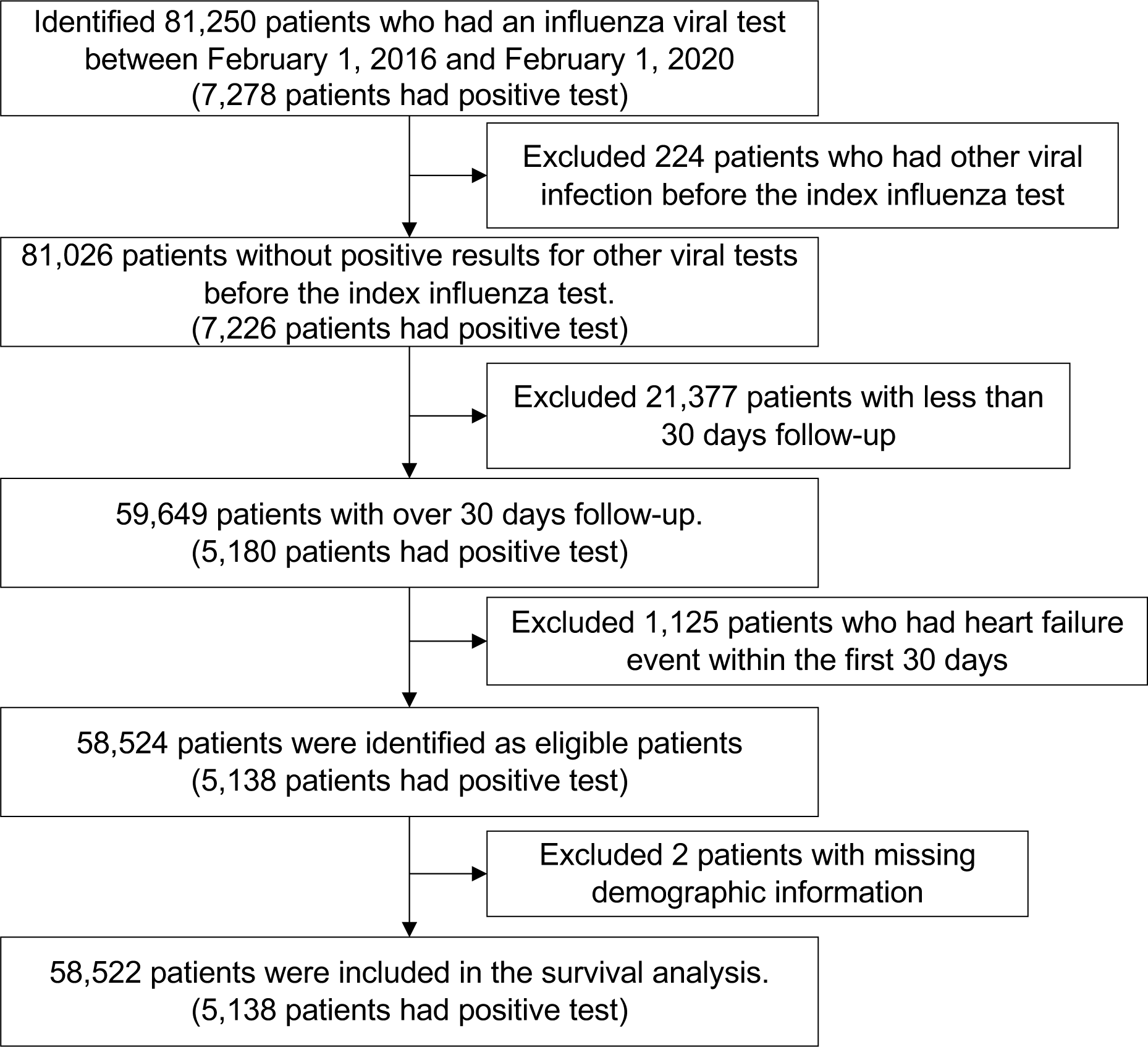
Flow diagram of patient selection for survival analysis on heart failure hospitalization in patients undergoing influenza viral tests.

**Figure S13.**
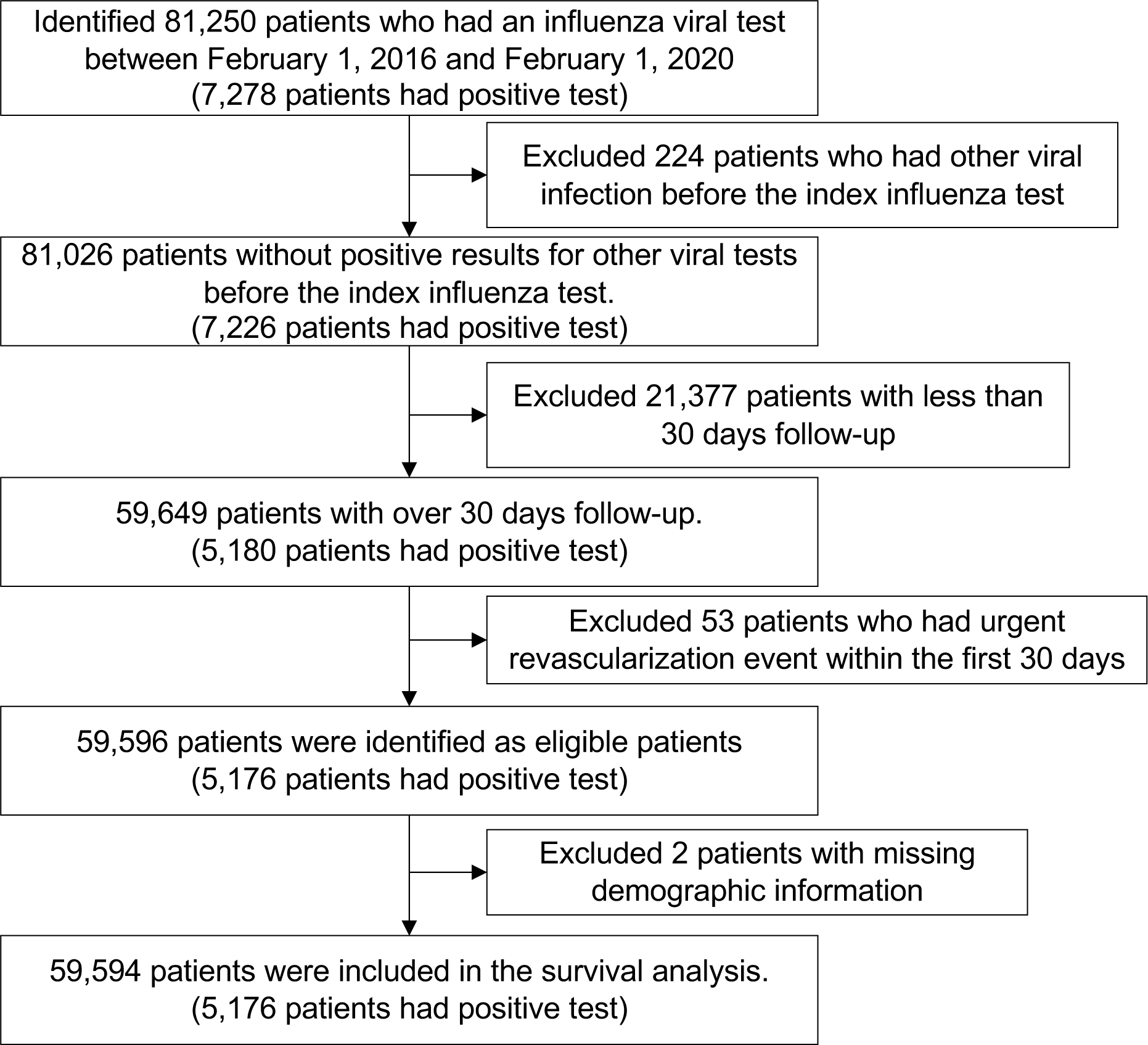
Flow diagram of patient selection for survival analysis on urgent revascularization in patients undergoing influenza viral tests.

**Figure S14.**
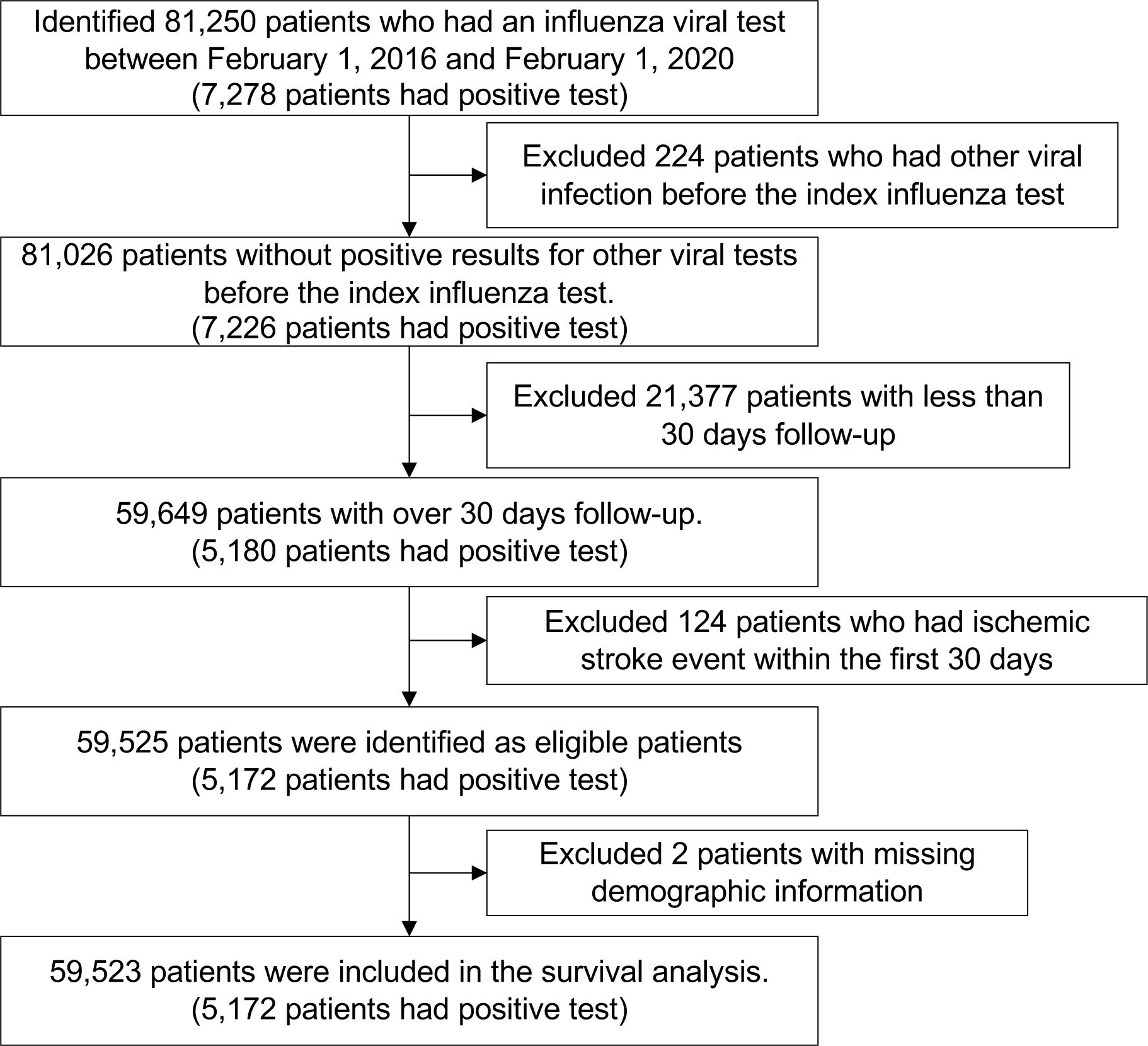
Flow diagram of patient selection for survival analysis on ischemic stroke in patients undergoing influenza viral tests.

**Figure S15.**
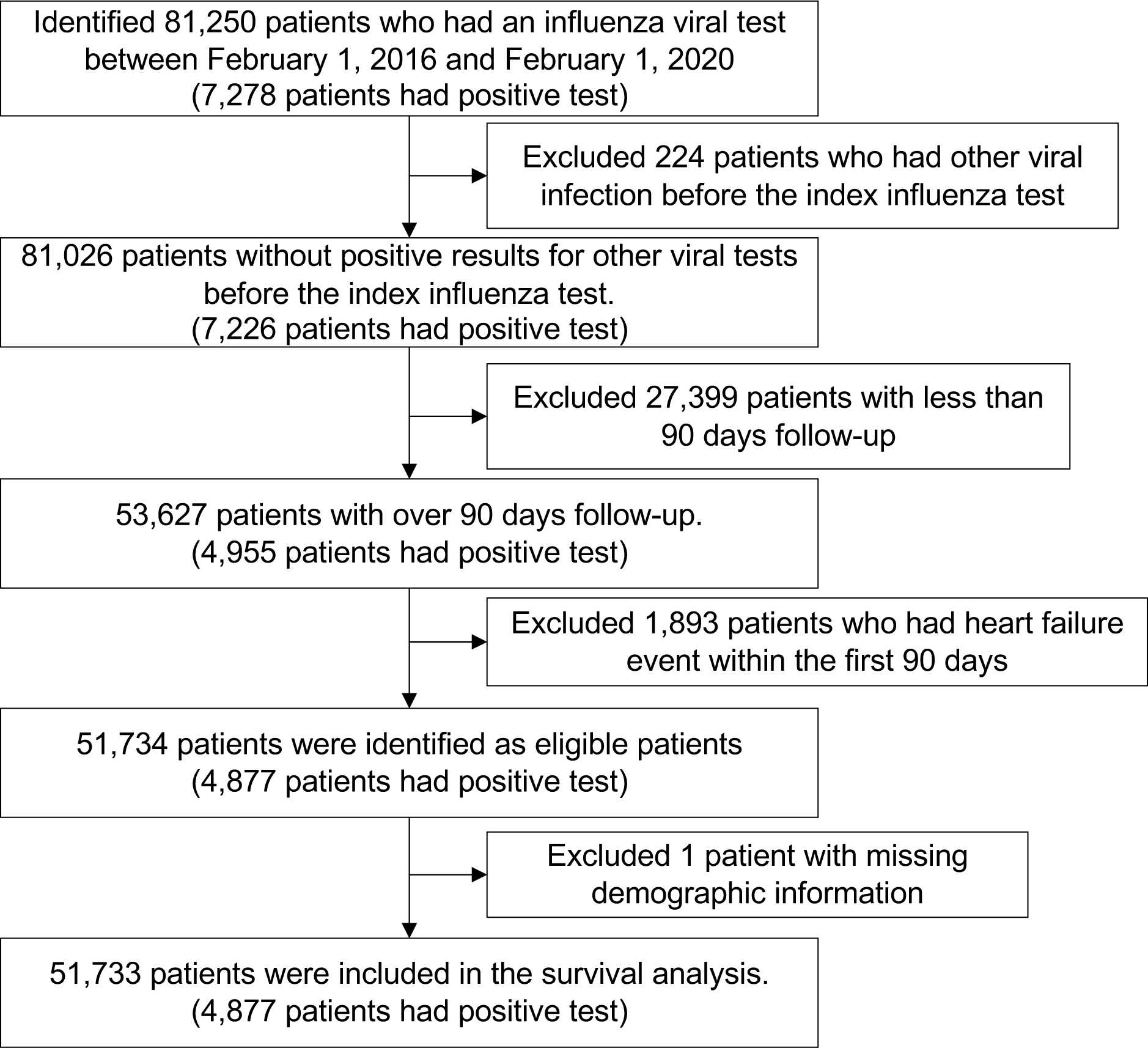
Flow diagram of patient selection for survival analysis on heart failure in patients undergoing influenza viral tests (excluding those who had event within the first 90 days).

**Figure S16.**
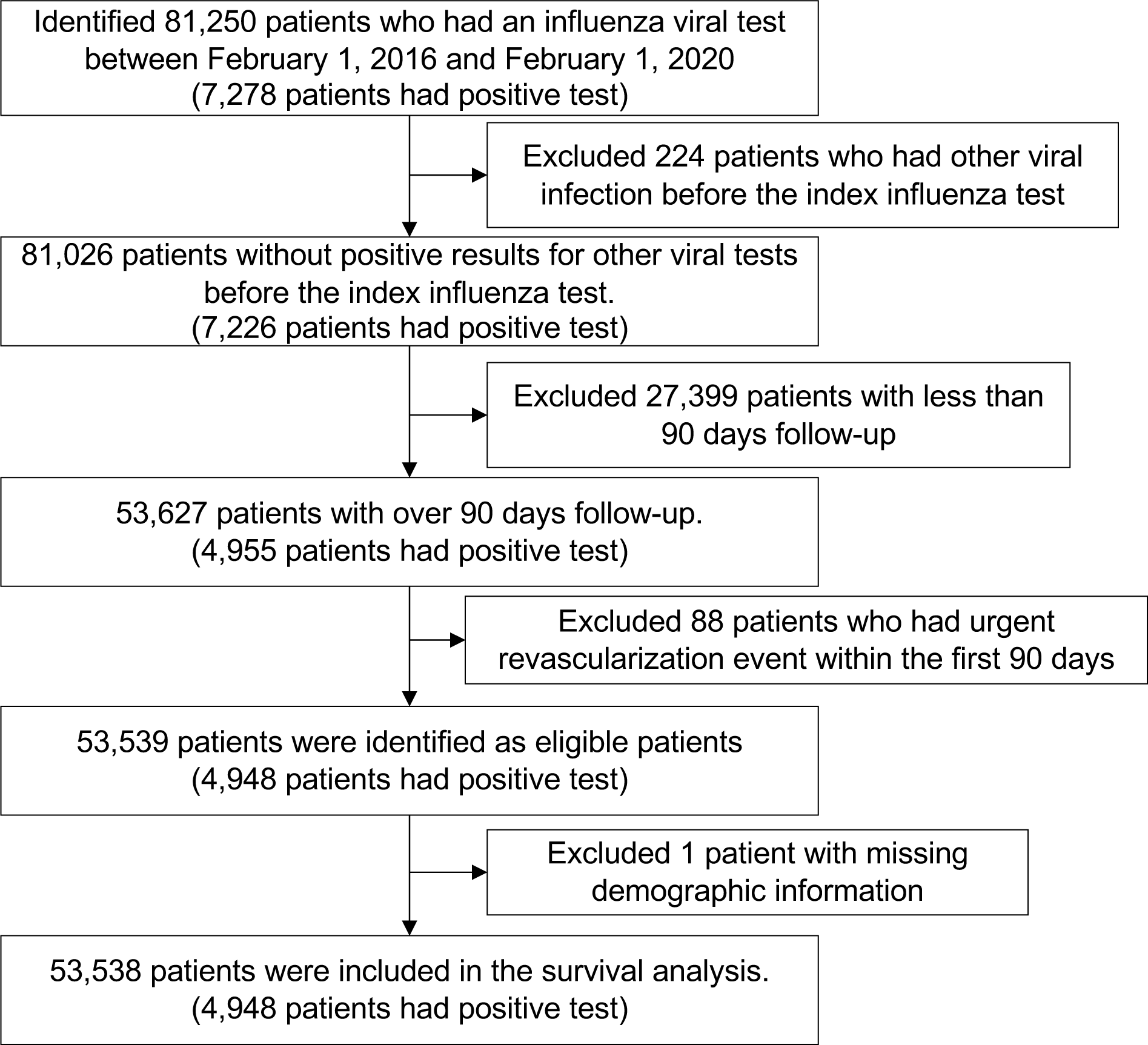
Flow diagram of patient selection for survival analysis on urgent revascularization in patients undergoing influenza viral tests (excluding those who had event within the first 90 days).

**Figure S17.**
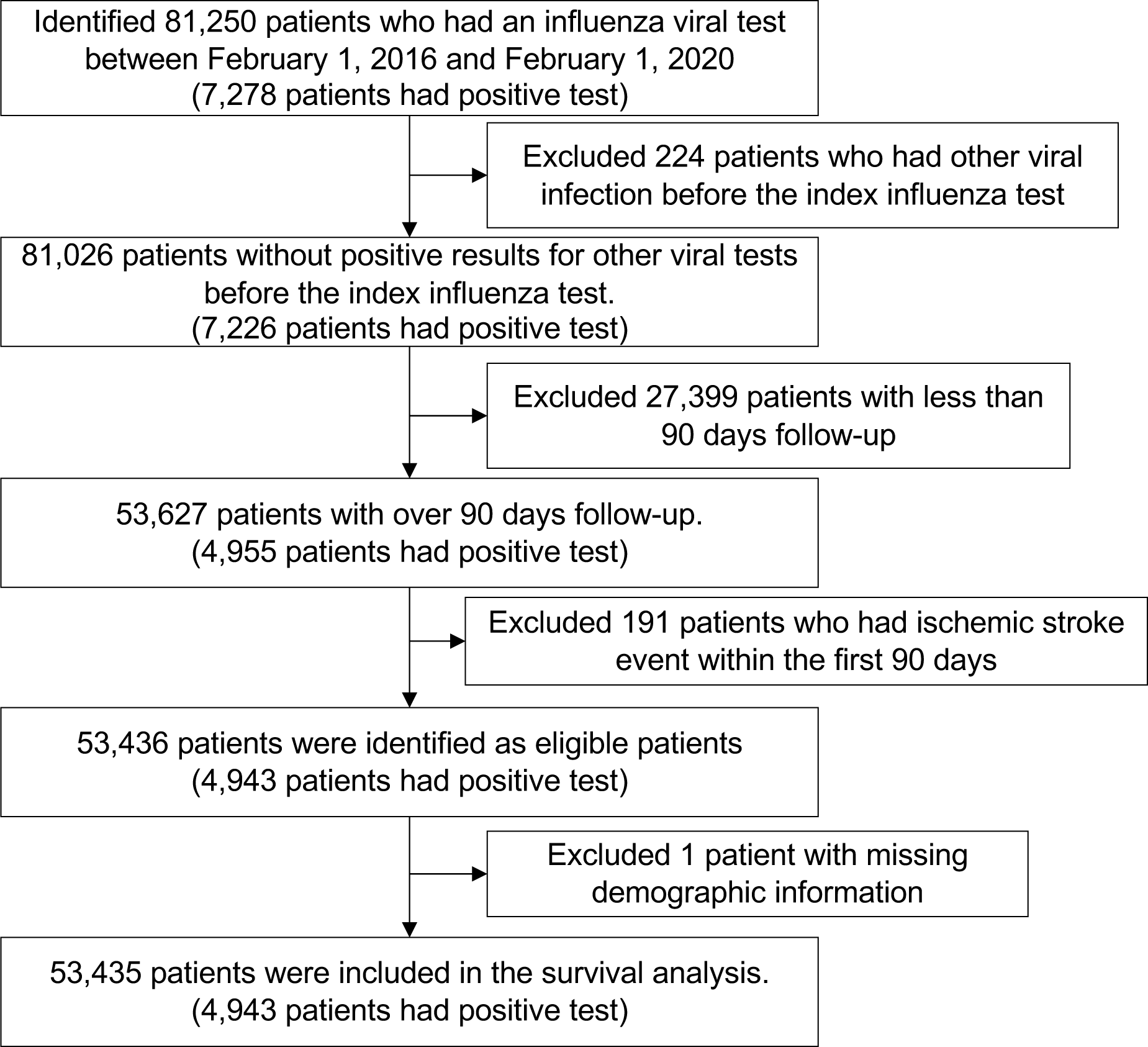
Flow diagram of patient selection for survival analysis on ischemic stroke in patients undergoing influenza viral tests (excluding those who had event within the first 90 days).

## Supplementary Tables

**Table S1.** Demographics for the notes used for training and testing event models

|  | Training dataset | Test dataset |
| --- | --- | --- |
| Numbers of notes | 1964 | 1003 |
| **Events** |  |  |
| PCI/CABG, n (%) | 306 (15.6) | 54 (5.4) |
| ACS, n (%) | 307 (15.6) | 54 (5.4) |
| Heart failure hospitalization, n (%) | 328 (16.7) | 122 (16.7) |
| Ischemic stroke, n (%) | 318 (16.2) | 59 (5.9) |
| **Institutions** |  |  |
| Brigham and Women's Hospital, n (%) | 602 (30.7) | 140 (14.0) |
| Brigham and Women's Faulkner Hospital, n (%) | 121 (6.2) | 110 (11.0) |
| Massachusetts General Hospital, n (%) | 632 (32.2) | 529 (52.7) |
| Cooley Dickinson Health Care, n (%) | 43 (2.2) | 0 (0.0) |
| Martha's Vineyard Hospital, n (%) | 6 (0.3) | 0 (0.0) |
| McLean Hospital, n (%) | 2 (0.1) | 0 (0.0) |
| Nantucket Cottage Hospital, n (%) | 1 (0.1) | 0 (0.0) |
| Massachusetts Eye and Ear, n (%) | 1 (0.1) | 0 (0.0) |
| Newton-Wellesley Hospital, n (%) | 155 (7.9) | 123 (12.3) |
| North Shore Medical Center, n (%) | 173 (8.8) | 101 (10.1) |
| South shore hospital, n (%) | 5 (0.3) | 0 (0.0) |
| Spaulding Hospital Cambridge, n (%) | 4 (0.2) | 0 (0.0) |
| Spaulding Hospital North Shore, n (%) | 4 (0.2) | 0 (0.0) |
| Spaulding Nursing and Therapy Center Brighton, n (%) | 1 (0.1) | 0 (0.0) |
| Spaulding Rehabilitation Hospital, n (%) | 15 (0.8) | 0 (0.0) |
| Union Hospital, n (%) | 1 (0.1) | 0 (0.0) |
| Wentworth-Douglass Hospital, n (%) | 18 (0.9) | 0 (0.0) |
| Unclear | 180 (9.2) | 0 (0.0) |

**Table S2.** Demographics for the notes used for training and testing history model

|  | Training dataset | Test dataset |
| --- | --- | --- |
| Numbers of notes | 819 | 366 |
| **HF history label** |  |  |
| Yes, n (%) | 181 (22.1) | 82 (22.4) |
| No, n (%) | 371 (45.3) | 171 (46.7) |
| Ambiguous, n (%) | 45 (5.5) | 19 (5.2) |
| Uninformative, n (%) | 222 (27.1) | 94 (25.7) |
| **Institutions** |  |  |
| Brigham and Women's Hospital, n (%) | 422 (51.5) | 189 (51.6) |
| Brigham and Women's Faulkner Hospital, n (%) | 37 (4.5) | 12 (3.3) |
| BWH Primary care Foxborough, n (%) | 24 (2.9) | 11 (3.0) |
| BWH Primary care Brookline, n (%) | 4 (0.5) | 3 (0.8) |
| BWH Primary care Norwood, n (%) | 2 (0.2) | 1 (0.3) |
| BWH Primary care Pembroke, n (%) | 0 (0.0) | 1 (0.3) |
| BWH at Newton Corner, n (%) | 4 (0.5) | 4 (1.1) |
| BWH Harbor Medical Associates, n (%) | 4 (0.5) | 3 (0.8) |
| Faulkner Community Physicians at Hyde Park, n (%) | 3 (0.4) | 0 (0.0) |
| Faulkner Community Physicians at West Roxbury, n (%) | 3 (0.4) | 4 (1.1) |
| Massachusetts General Hospital, n (%) | 102 (12.5) | 37 (10.1) |
| MGH West Medical Group, n (%) | 0 (0.0) | 1 (0.3) |
| MGH Danvers, n (%) | 0 (0.0) | 1 (0.3) |
| MGH Waltham, n (%) | 1 (0.1) | 1 (0.3) |
| MGH North Shore Center, n (%) | 1 (0.1) | 0 (0.0) |
| MGH Beacon Hill Primary Care, n (%) | 0 (0.0) | 1 (0.3) |
| MGH Downtown Primary Care, n (%) | 1 (0.1) | 0 (0.0) |
| MGB Urgent care, n (%) | 0 (0.0) | 2 (0.5) |
| Foxborough Health Care Center, n (%) | 21 (2.6) | 14 (3.8) |
| Dana-Farber Cancer Institute, n (%) | 60 (7.3) | 22 (6.0) |
| Essex Neurological Associates, n (%) | 2 (0.2) | 0 (0.0) |
| Lown Cardiovascular Group, n (%) | 1 (0.1) | 0 (0.0) |
| Cooley Dickinson Health Care, n (%) | 1 (0.1) | 1 (0.3) |
| Charles River Medical Associates, n (%) | 12 (1.5) | 8 (2.2) |
| Martha's Vineyard Hospital, n (%) | 2 (0.2) | 1 (0.3) |
| Milford Regional Physician Group, n (%) | 1 (0.1) | 1 (0.3) |
| Nantucket Cottage Hospital, n (%) | 1 (0.1) | 0 (0.0) |
| Massachusetts Eye and Ear, n (%) | 9 (1.1) | 5 (1.4) |
| Newton-Wellesley Hospital, n (%) | 34 (4.2) | 13 (3.6) |
| North End Waterfront Health, n (%) | 2 (0.2) | 0 (0.0) |
| North Shore Medical Center, n (%) | 40 (4.9) | 15 (4.1) |
| Ophthalmic Consultants of Boston, n (%) | 2 (0.2) | 0 (0.0) |
| Ophthalmic Consultants of Boston in Plymouth, n (%) | 1 (0.1) | 0 (0.0) |
| Ophthalmic Consultants of Boston in Yarmouth, n (%) | 2 (0.2) | 4 (1.1) |
| Peabody Podiatry, n (%) | 0 (0.0) | 2 (0.5) |
| Pentucket Medical Haverhill, n (%) | 1 (0.1) | 0 (0.0) |
| South shore hospital, n (%) | 2 (0.2) | 1 (0.3) |
| Spaulding Rehabilitation Hospital, n (%) | 2 (0.2) | 1 (0.3) |
| Spaulding Hospital Cambridge, n (%) | 2 (0.2) | 1 (0.3) |
| Spaulding Hospital Braintree, n (%) | 1 (0.1) | 0 (0.0) |
| Spaulding Hospital Framingham, n (%) | 1 (0.1) | 0 (0.0) |
| Spaulding Hospital Malden, n (%) | 0 (0.0) | 1 (0.3) |
| Spaulding Hospital Peabody, n (%) | 0 (0.0) | 3 (0.8) |
| Spaulding Hospital Salem, n (%) | 1 (0.1) | 0 (0.0) |
| Spaulding Hospital Wellesley, n (%) | 1 (0.1) | 0 (0.0) |
| Winchester Hospital, n (%) | 2 (0.2) | 0 (0.0) |
| Unclear | 7 (0.9) | 2 (0.5) |

BWH: Brigham and Women's Hospital, MGH: Massachusetts General Hospital, MGB: Mass General Brigham.

**Table S3.** Baseline characteristics for survival analysis of heart failure hospitalization stratified by RSV infection (unmatched)

|  | RSV positive | RSV negative | SMD |
| --- | --- | --- | --- |
| Number patients | 1531 | 33,525 |  |
| **Demographics** |  |  |  |
| Age, years ± SD | 64.6 ± 18.3 | 59.1 ± 19.3 | 0.292 |
| Gender, Male (%) | 643 (42.0) | 14,801 (44.1) | 0.043 |
| History of Smoking, Yes (%) | 731 (47.7) | 15,172 (45.3) | 0.050 |
| Race |  |  | 0.085 |
| White, n (%) | 1,143 (74.7) | 23,942 (71.4) |  |
| Black, n (%) | 156 (10.2) | 3,686 (11.0) |  |
| Hispanic, n (%) | 43 (2.8) | 1,087 (3.2) |  |
| Asian, n (%) | 49 (3.2) | 1,249 (3.7) |  |
| Others, n (%) | 107 (7.0) | 2,530 (7.5) |  |
| Unknown, n (%) | 33 (2.2) | 1,031 (3.1) |  |
| **Disease history** |  |  |  |
| Heart failure, n (%) | 355 (23.2) | 5,337 (15.9) | 0.184 |
| CAD, n (%) | 361 (23.6) | 5,693 (17.0) | 0.165 |
| COPD, n (%) | 413 (27.0) | 6,257 (18.7) | 0.196 |
| Hyperlipidemia, n (%) | 823 (53.8) | 14,729 (43.9) | 0.197 |
| Hypertension, n (%) | 1,014 (66.2) | 18,533 (55.3) | 0.226 |
| Atrial fibrillation, n (%) | 332 (21.7) | 5,011 (14.9) | 0.175 |
| Type 2 DM, n (%) | 444 (29.0) | 7,809 (23.3) | 0.130 |
| **Medication** |  |  |  |
| ASA, n (%) | 626 (40.9) | 11,879 (35.4) | 0.112 |
| P2Y<sub>12</sub> Inhibitor, n (%) | 74 (4.8) | 1,510 (4.5) | 0.016 |
| Statin | 718 (46.9) | 13,133 (39.2) | 0.156 |
| ACEi, n (%) | 300 (19.6) | 6,689 (20.0) | 0.009 |
| ARB, n (%) | 215 (14.0) | 3,631 (10.8) | 0.097 |
| Beta blocker, n (%) | 654 (42.7) | 11,645 (34.7) | 0.164 |
| Loop diuretic, n (%) | 445 (29.1) | 6,601 (19.7) | 0.220 |
| MRA, n (%) | 71 (4.6) | 1,313 (3.9) | 0.036 |
| **Virus testing method** |  |  |  |
| Method |  |  | 0.023 |
| A, n (%) | 784 (51.2) | 16,788 (50.1) |  |
| B, n (%) | 747 (48.8) | 16,737 (49.9) |  |

Abbreviations. SMD: standardized mean difference, CAD: coronary heart disease, COPD: chronic obstructive pulmonary disease, DM: diabetes mellitus, ASA: acetyl-salicylic acid, ACEi: angiotensin-converting enzyme inhibitor, ARB: angiotensin II receptor blocker, MRA: mineralocorticoid receptor antagonist

**Table S4.** Baseline characteristics for survival analysis of urgent revascularization stratified by RSV infection (unmatched)

|  | RSV positive | RSV negative | SMD |
| --- | --- | --- | --- |
| Number patients | 1572 | 34,255 |  |
| **Demographics** |  |  |  |
| Age, years ± SD | 64.8 ± 18.2 | 59.4 ± 19.3 | 0.287 |
| Gender, Male (%) | 660 (42.0) | 15,152 (44.2) | 0.045 |
| History of Smoking, Yes (%) | 758 (48.2) | 15,583 (45.5) | 0.055 |
| Race |  |  | 0.081 |
| White, n (%) | 1,171 (74.5) | 24,515 (71.6) |  |
| Black, n (%) | 163 (10.4) | 3,761 (11.0) |  |
| Hispanic, n (%) | 43 (2.7) | 1,103 (3.2) |  |
| Asian, n (%) | 49 (3.1) | 1,268 (3.7) |  |
| Others, n (%) | 112 (7.1) | 2,562 (7.5) |  |
| Unknown, n (%) | 34 (2.2) | 1,046 (3.1) |  |
| **Disease history** |  |  |  |
| Heart failure, n (%) | 386 (24.6) | 5,834 (17.0) | 0.186 |
| CAD, n (%) | 373 (23.7) | 5,966 (17.4) | 0.157 |
| COPD, n (%) | 430 (27.4) | 6,522 (19.0) | 0.198 |
| Hyperlipidemia, n (%) | 853 (54.3) | 15,202 (44.4) | 0.199 |
| Hypertension, n (%) | 1,050 (66.8) | 19,144 (55.9) | 0.225 |
| Atrial fibrillation, n (%) | 350 (22.3) | 5,365 (15.7) | 0.169 |
| Type 2 DM, n (%) | 461 (29.3) | 8,087 (23.6) | 0.130 |
| **Medication** |  |  |  |
| ASA, n (%) | 652 (41.5) | 12,275 (35.8) | 0.116 |
| P2Y<sub>12</sub> Inhibitor, n (%) | 76 (4.8) | 1,576 (4.6) | 0.011 |
| Statin | 748 (47.6) | 13,574 (39.6) | 0.161 |
| ACEi, n (%) | 313 (19.9) | 6,900 (20.1) | 0.006 |
| ARB, n (%) | 222 (14.1) | 3,772 (11.0) | 0.094 |
| Beta blocker, n (%) | 681 (43.3) | 12,139 (35.4) | 0.162 |
| Loop diuretic, n (%) | 475 (30.2) | 7,090 (20.7) | 0.220 |
| MRA, n (%) | 80 (5.1) | 1,397 (4.1) | 0.048 |
| **Virus testing method** |  |  |  |
| Method |  |  | 0.026 |
| A, n (%) | 808 (51.4) | 17,159 (50.1) |  |
| B, n (%) | 764 (48.6) | 17,096 (49.9) |  |

Abbreviations. SMD: standardized mean difference, CAD: coronary heart disease, COPD: chronic obstructive pulmonary disease, DM: diabetes mellitus, ASA: acetyl-salicylic acid, ACEi: angiotensin-converting enzyme inhibitor, ARB: angiotensin II receptor blocker, MRA: mineralocorticoid receptor antagonist

**Table S5.** Baseline characteristics for survival analysis of stroke stratified by RSV infection (unmatched)

|  | RSV positive | RSV negative | SMD |
| --- | --- | --- | --- |
| Number patients | 1570 | 34,208 |  |
| **Demographics** |  |  |  |
| Age, years ± SD | 64.8 ± 18.2 | 59.4 ± 19.3 | 0.288 |
| Gender, Male (%) | 659 (42.0) | 15,133 (44.2) | 0.046 |
| History of Smoking, Yes (%) | 756 (48.2) | 15,573 (45.5) | 0.053 |
| Race |  |  | 0.079 |
| White, n (%) | 1,169 (74.5) | 24,494 (71.6) |  |
| Black, n (%) | 163 (10.4) | 3,747 (11.0) |  |
| Hispanic, n (%) | 43 (2.7) | 1,103 (3.2) |  |
| Asian, n (%) | 49 (3.1) | 1,262 (3.7) |  |
| Others, n (%) | 112 (7.1) | 2,561 (7.5) |  |
| Unknown, n (%) | 34 (2.2) | 1,041 (3.0) |  |
| **Disease history** |  |  |  |
| Heart failure, n (%) | 386 (24.6) | 5,824 (17.0) | 0.187 |
| CAD, n (%) | 374 (23.8) | 5,977 (17.5) | 0.157 |
| COPD, n (%) | 428 (27.3) | 6,511 (19.0) | 0.196 |
| Hyperlipidemia, n (%) | 854 (54.4) | 15,179 (44.4) | 0.201 |
| Hypertension, n (%) | 1,047 (66.7) | 19,108 (55.9) | 0.224 |
| Atrial fibrillation, n (%) | 351 (22.4) | 5,342 (15.6) | 0.173 |
| Type 2 DM, n (%) | 460 (29.3) | 8,069 (23.6) | 0.130 |
| **Medication** |  |  |  |
| ASA, n (%) | 653 (41.6) | 12,256 (35.8) | 0.119 |
| P2Y<sub>12</sub> Inhibitor, n (%) | 76 (4.8) | 1,571 (4.6) | 0.012 |
| Statin | 749 (47.7) | 13,548 (39.6) | 0.164 |
| ACEi, n (%) | 311 (19.8) | 6,882 (20.1) | 0.008 |
| ARB, n (%) | 222 (14.1) | 3,770 (11.0) | 0.094 |
| Beta blocker, n (%) | 681 (43.4) | 12,115 (35.4) | 0.163 |
| Loop diuretic, n (%) | 474 (30.2) | 7,084 (20.7) | 0.219 |
| MRA, n (%) | 80 (5.1) | 1,399 (4.1) | 0.048 |
| **Virus testing method** |  |  |  |
| Method |  |  | 0.027 |
| A, n (%) | 808 (51.5) | 17,135 (50.1) |  |
| B, n (%) | 762 (48.5) | 17,073 (49.9) |  |

Abbreviations. SMD: standardized mean difference, CAD: coronary heart disease, COPD: chronic obstructive pulmonary disease, DM: diabetes mellitus, ASA: acetyl-salicylic acid, ACEi: angiotensin-converting enzyme inhibitor, ARB: angiotensin II receptor blocker, MRA: mineralocorticoid receptor antagonist

**Table S6.** Baseline characteristics for survival analysis of heart failure hospitalization stratified by RSV infection (matched)

|  | RSV positive | RSV negative | SMD |
| --- | --- | --- | --- |
| Number patients | 1531 | 24,496 |  |
| **Demographics** |  |  |  |
| Age, years ± SD | 64.6 ± 18.3 | 64.2 ± 17.7 | 0.023 |
| Gender, Male (%) | 643 (42.0) | 10,445 (42.6) | 0.013 |
| History of Smoking, Yes (%) | 731 (47.7) | 11,524 (47.0) | 0.014 |
| Race |  |  | 0.019 |
| White, n (%) | 1,143 (74.7) | 18,153 (74.1) |  |
| Black, n (%) | 156 (10.2) | 2,571 (10.5) |  |
| Hispanic, n (%) | 43 (2.8) | 716 (2.9) |  |
| Asian, n (%) | 49 (3.2) | 795 (3.2) |  |
| Others, n (%) | 107 (7.0) | 1,769 (7.2) |  |
| Unknown, n (%) | 33 (2.2) | 492 (2.0) |  |
| **Disease history** |  |  |  |
| Heart failure, n (%) | 355 (23.2) | 5,030 (20.5) | 0.064 |
| CAD, n (%) | 361 (23.6) | 5,315 (21.7) | 0.045 |
| COPD, n (%) | 413 (27.0) | 5,947 (24.3) | 0.062 |
| Hyperlipidemia, n (%) | 823 (53.8) | 12,933 (52.8) | 0.019 |
| Hypertension, n (%) | 1,014 (66.2) | 16,043 (65.5) | 0.016 |
| Atrial fibrillation, n (%) | 332 (21.7) | 4,746 (19.4) | 0.057 |
| Type 2 DM, n (%) | 444 (29.0) | 6,910 (28.2) | 0.018 |
| **Medication** |  |  |  |
| ASA, n (%) | 626 (40.9) | 9,979 (40.7) | 0.003 |
| P2Y<sub>12</sub> Inhibitor, n (%) | 74 (4.8) | 1,224 (5.0) | 0.008 |
| Statin | 718 (46.9) | 11,501 (47.0) | 0.001 |
| ACEi, n (%) | 300 (19.6) | 5,686 (23.2) | 0.088 |
| ARB, n (%) | 215 (14.0) | 3,240 (13.2) | 0.024 |
| Beta blocker, n (%) | 654 (42.7) | 10,108 (41.3) | 0.029 |
| Loop diuretic, n (%) | 445 (29.1) | 6,282 (25.6) | 0.077 |
| MRA, n (%) | 71 (4.6) | 1,168 (4.8) | 0.006 |
| **Virus testing method** |  |  |  |
| Method |  |  | <0.001 |
| A, n (%) | 784 (51.2) | 12,539 (51.2) |  |
| B, n (%) | 747 (48.8) | 11,957 (48.8) |  |

Abbreviations. SMD: standardized mean difference, CAD: coronary heart disease, COPD: chronic obstructive pulmonary disease, DM: diabetes mellitus, ASA: acetyl-salicylic acid, ACEi: angiotensin-converting enzyme inhibitor, ARB: angiotensin II receptor blocker, MRA: mineralocorticoid receptor antagonist

**Table S7.** Baseline characteristics for survival analysis of urgent revascularization stratified by RSV infection (matched)

|  | RSV positive | RSV negative | SMD |
| --- | --- | --- | --- |
| Number patients | 1572 | 25,152 |  |
| **Demographics** |  |  |  |
| Age, years ± SD | 64.8 ± 18.2 | 64.4 ± 17.7 | 0.022 |
| Gender, Male (%) | 660 (42.0) | 10,670 (42.4) | 0.009 |
| History of Smoking, Yes (%) | 758 (48.2) | 12,039 (47.9) | 0.007 |
| Race |  |  | 0.011 |
| White, n (%) | 1,171 (74.5) | 18,654 (74.2) |  |
| Black, n (%) | 163 (10.4) | 2,666 (10.6) |  |
| Hispanic, n (%) | 43 (2.7) | 673 (2.7) |  |
| Asian, n (%) | 49 (3.1) | 793 (3.2) |  |
| Others, n (%) | 112 (7.1) | 1,830 (7.3) |  |
| Unknown, n (%) | 34 (2.2) | 536 (2.1) |  |
| **Disease history** |  |  |  |
| Heart failure, n (%) | 386 (24.6) | 5,450 (21.7) | 0.069 |
| CAD, n (%) | 373 (23.7) | 5,524 (22.0) | 0.042 |
| COPD, n (%) | 430 (27.4) | 6,210 (24.7) | 0.061 |
| Hyperlipidemia, n (%) | 853 (54.3) | 13,399 (53.3) | 0.020 |
| Hypertension, n (%) | 1,050 (66.8) | 16,623 (66.1) | 0.015 |
| Atrial fibrillation, n (%) | 350 (22.3) | 5,060 (20.1) | 0.053 |
| Type 2 DM, n (%) | 461 (29.3) | 7,167 (28.5) | 0.018 |
| **Medication** |  |  |  |
| ASA, n (%) | 652 (41.5) | 10,418 (41.4) | 0.001 |
| P2Y<sub>12</sub> Inhibitor, n (%) | 76 (4.8) | 1,230 (4.9) | 0.003 |
| Statin | 748 (47.6) | 11,928 (47.4) | 0.003 |
| ACEi, n (%) | 313 (19.9) | 5,880 (23.4) | 0.084 |
| ARB, n (%) | 222 (14.1) | 3,354 (13.3) | 0.023 |
| Beta blocker, n (%) | 681 (43.3) | 10,558 (42.0) | 0.027 |
| Loop diuretic, n (%) | 475 (30.2) | 6,759 (26.9) | 0.074 |
| MRA, n (%) | 80 (5.1) | 1,233 (4.9) | 0.009 |
| **Virus testing method** |  |  |  |
| Method |  |  | 0.002 |
| A, n (%) | 808 (51.4) | 12,901 (51.4) |  |
| B, n (%) | 764 (48.6) | 12,251 (48.6) |  |

Abbreviations. SMD: standardized mean difference, CAD: coronary heart disease, COPD: chronic obstructive pulmonary disease, DM: diabetes mellitus, ASA: acetyl-salicylic acid, ACEi: angiotensin-converting enzyme inhibitor, ARB: angiotensin II receptor blocker, MRA: mineralocorticoid receptor antagonist

**Table S8.**
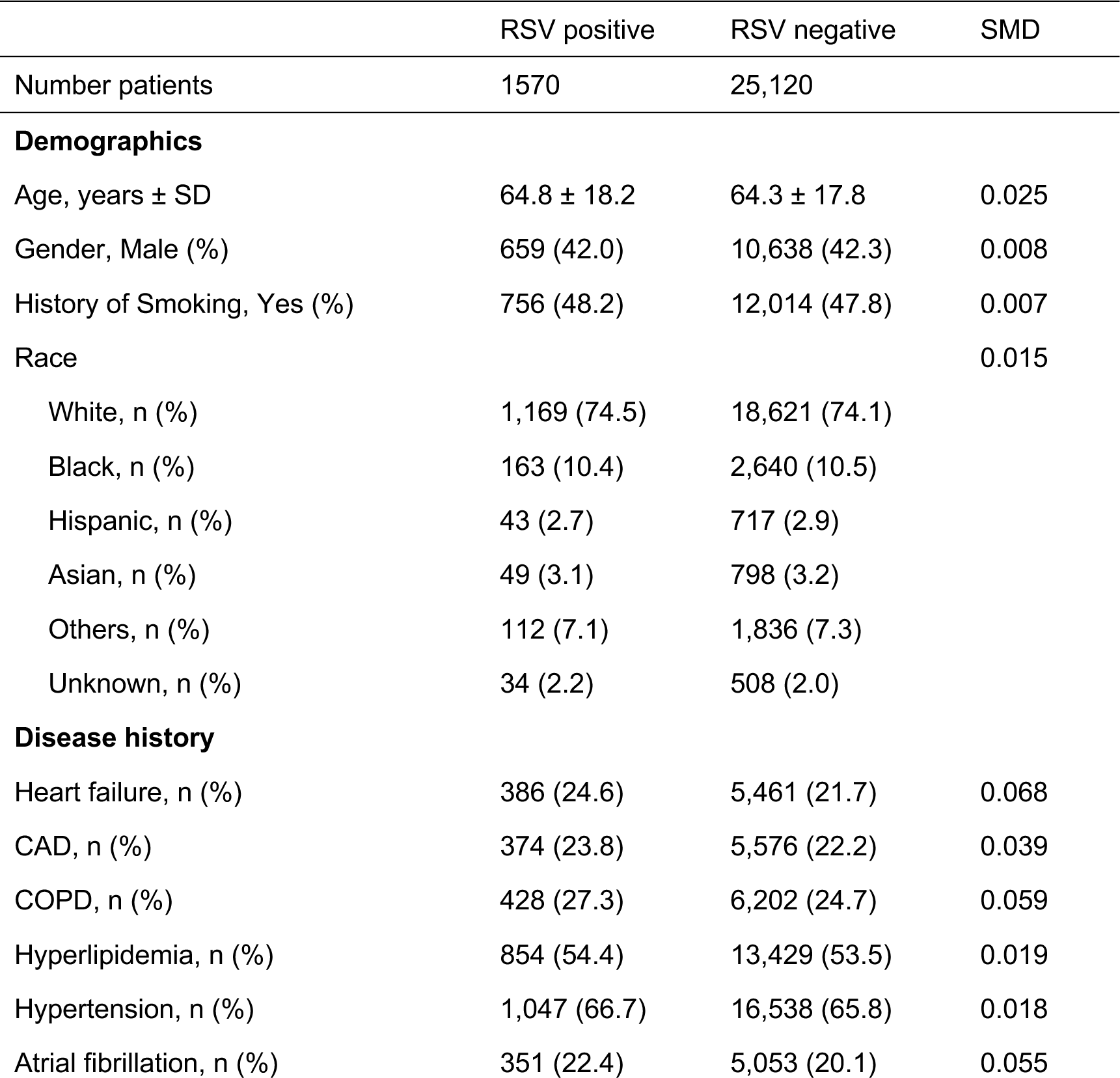
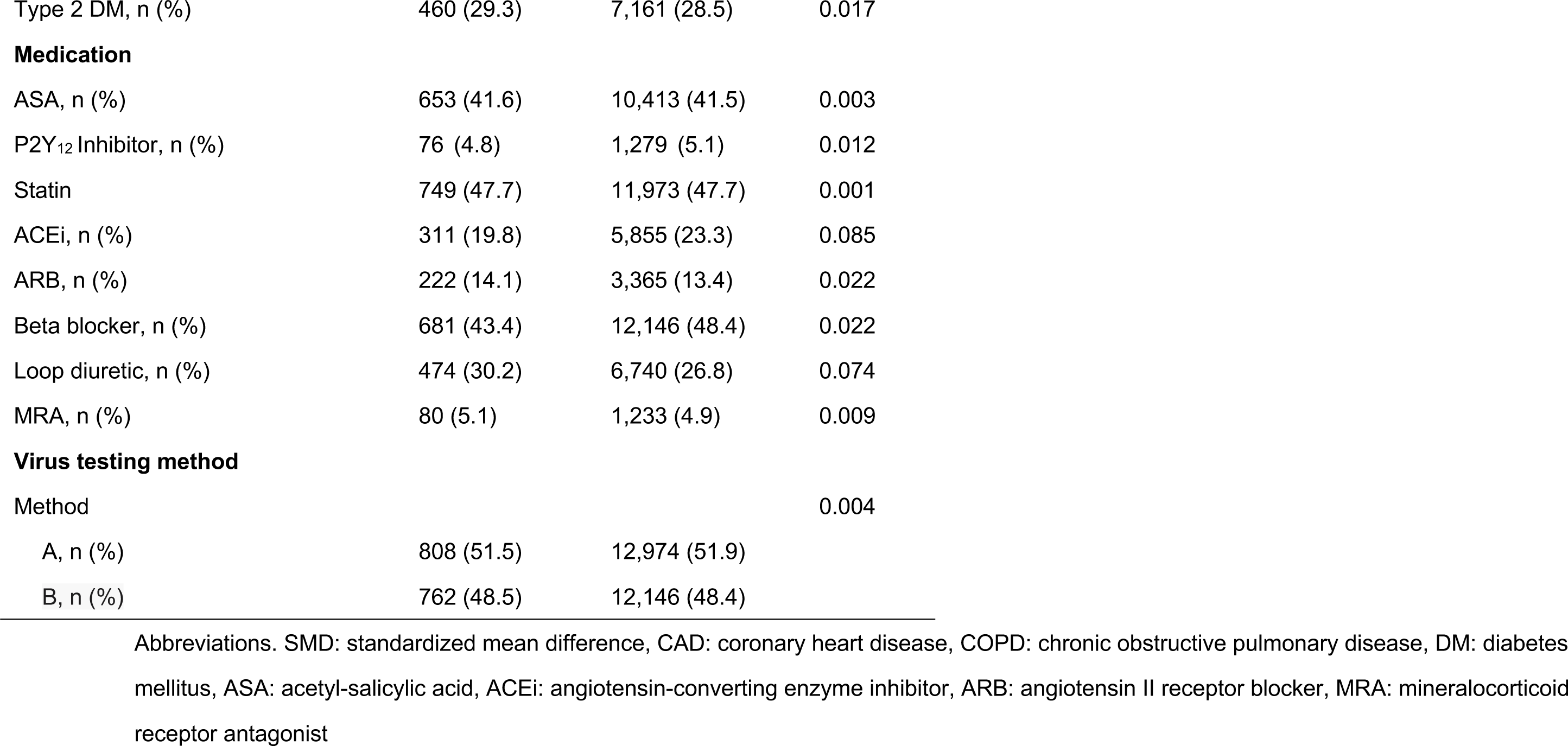
Baseline characteristics for survival analysis of stroke stratified by RSV infection (matched)

**Table S9.** Baseline characteristics for survival analysis of heart failure stratified by RSV infection (with 90 days exclusion window, unmatched)

|  | RSV positive | RSV negative | SMD |
| --- | --- | --- | --- |
| Number patients | 1,263 | 28,588 |  |
| **Demographics** |  |  |  |
| Age, years ± SD | 64.1 ± 18.3 | 58.6 ± 19.2 | 0.294 |
| Gender, Male (%) | 537 (42.5) | 12,405 (43.4) | 0.018 |
| History of Smoking, Yes (%) | 599 (47.4) | 12,770 (44.7) | 0.055 |
| Race |  |  | 0.117 |
| White, n (%) | 953 (75.5) | 20,254 (70.8) |  |
| Black, n (%) | 123 (9.7) | 3,227 (11.3) |  |
| Hispanic, n (%) | 32 (2.5) | 991 (3.5) |  |
| Asian, n (%) | 43 (3.4) | 1,069 (3.7) |  |
| Others, n (%) | 87 (6.9) | 2,178 (7.6) |  |
| Unknown, n (%) | 25 (2.0) | 869 (3.0) |  |
| **Disease history** |  |  |  |
| Heart failure, n (%) | 272 (21.5) | 4,329 (15.1) | 0.166 |
| CAD, n (%) | 271 (21.5) | 4,805 (16.8) | 0.118 |
| COPD, n (%) | 347 (27.5) | 5,380 (18.8) | 0.206 |
| Hyperlipidemia, n (%) | 671 (53.1) | 12,720 (44.5) | 0.173 |
| Hypertension, n (%) | 837 (66.3) | 15,909 (55.6) | 0.219 |
| Atrial fibrillation, n (%) | 257 (14.5) | 4,155 (14.5) | 0.154 |
| Type 2 DM, n (%) | 351 (23.3) | 6,651 (23.3) | 0.104 |
| **Medication** |  |  |  |
| ASA, n (%) | 500 (39.6) | 10,079 (35.3) | 0.090 |
| P2Y<sub>12</sub> Inhibitor, n (%) | 58 (4.6) | 1,230 (4.3) | 0.014 |
| Statin | 589 (46.6) | 11,043 (38.6) | 0.162 |
| ACEi, n (%) | 248 (19.6) | 5,715 (20.0) | 0.009 |
| ARB, n (%) | 176 (13.9) | 3,106 (10.9) | 0.093 |
| Beta blocker, n (%) | 525 (41.6) | 9,701 (33.9) | 0.158 |
| Loop diuretic, n (%) | 336 (26.6) | 5,204 (18.2) | 0.202 |
| MRA, n (%) | 54 (4.3) | 1,033 (3.6) | 0.034 |
| **Virus testing method** |  |  |  |
| Method |  |  | 0.025 |
| A, n (%) | 652 (51.6) | 14,403 (50.4) |  |
| B, n (%) | 611 (48.4) | 14,185 (49.6) |  |

Abbreviations. SMD: standardized mean difference, CAD: coronary heart disease, COPD: chronic obstructive pulmonary disease, DM: diabetes mellitus, ASA: acetyl-salicylic acid, ACEi: angiotensin-converting enzyme inhibitor, ARB: angiotensin II receptor blocker, MRA: mineralocorticoid receptor antagonist

**Table S10.** Baseline characteristics for survival analysis of urgent revascularization stratified by RSV infection (with 90 days exclusion window, unmatched)

|  | RSV positive | RSV negative | SMD |
| --- | --- | --- | --- |
| Number patients | 1,340 | 29,781 |  |
| **Demographics** |  |  |  |
| Age, years ± SD | 64.6 ± 18.2 | 59.1 ± 19.2 | 0.295 |
| Gender, Male (%) | 568 (42.4) | 13,001 (43.7) | 0.026 |
| History of Smoking, Yes (%) | 645 (48.1) | 13,438 (45.1) | 0.060 |
| Race |  |  | 0.103 |
| White, n (%) | 1,008 (75.2) | 21,188 (71.1) |  |
| Black, n (%) | 134 (10.0) | 3,356 (11.3) |  |
| Hispanic, n (%) | 33 (2.5) | 1,017 (3.4) |  |
| Asian, n (%) | 43 (3.2) | 1,097 (3.7) |  |
| Others, n (%) | 93 (6.9) | 2,228 (7.5) |  |
| Unknown, n (%) | 29 (2.2) | 895 (3.0) |  |
| **Disease history** |  |  |  |
| Heart failure, n (%) | 342 (25.5) | 5,230 (17.6) | 0.195 |
| CAD, n (%) | 311 (23.2) | 5,283 (17.7) | 0.136 |
| COPD, n (%) | 381 (28.4) | 5,819 (19.5) | 0.209 |
| Hyperlipidemia, n (%) | 732 (54.6) | 13,531 (45.4) | 0.185 |
| Hypertension, n (%) | 909 (67.8) | 16,947 (56.9) | 0.227 |
| Atrial fibrillation, n (%) | 298 (22.2) | 4,739 (15.9) | 0.162 |
| Type 2 DM, n (%) | 391 (23.3) | 7,178 (24.1) | 0.115 |
| **Medication** |  |  |  |
| ASA, n (%) | 548 (40.9) | 10,748 (36.1) | 0.099 |
| P2Y<sub>12</sub> Inhibitor, n (%) | 61 (4.6) | 1,347 (4.5) | 0.001 |
| Statin | 647 (48.3) | 11,815 (39.7) | 0.174 |
| ACEi, n (%) | 271 (20.2) | 6,075 (20.4) | 0.004 |
| ARB, n (%) | 193 (14.4) | 3,320 (11.1) | 0.098 |
| Beta blocker, n (%) | 583 (43.5) | 10,529 (35.4) | 0.167 |
| Loop diuretic, n (%) | 403 (30.1) | 6,055 (20.3) | 0.226 |
| MRA, n (%) | 69 (5.1) | 1,199 (4.0) | 0.054 |
| **Virus testing method** |  |  |  |
| Method |  |  | 0.026 |
| A, n (%) | 693 (51.7) | 15,020 (50.4) |  |
| B, n (%) | 647 (48.3) | 14,761 (49.6) |  |

Abbreviations. SMD: standardized mean difference, CAD: coronary heart disease, COPD: chronic obstructive pulmonary disease, DM: diabetes mellitus, ASA: acetyl-salicylic acid, ACEi: angiotensin-converting enzyme inhibitor, ARB: angiotensin II receptor blocker, MRA: mineralocorticoid receptor antagonist

**Table S11.** Baseline characteristics for survival analysis of ischemic stroke stratified by RSV infection (with 90 days exclusion window, unmatched)

|  | RSV positive | RSV negative | SMD |
| --- | --- | --- | --- |
| Number patients | 1,339 | 29,726 |  |
| **Demographics** |  |  |  |
| Age, years ± SD | 64.6 ± 18.2 | 59.1 ± 19.2 | 0.297 |
| Gender, Male (%) | 569 (42.5) | 12,981 (43.7) | 0.024 |
| History of Smoking, Yes (%) | 644 (48.1) | 13,426 (45.2) | 0.059 |
| Race |  |  | 0.101 |
| White, n (%) | 1,006 (75.1) | 21,169 (71.2) |  |
| Black, n (%) | 135 (10.1) | 3,333 (11.2) |  |
| Hispanic, n (%) | 33 (2.5) | 1,017 (3.4) |  |
| Asian, n (%) | 43 (3.2) | 1,090 (3.7) |  |
| Others, n (%) | 93 (6.9) | 2,224 (7.5) |  |
| Unknown, n (%) | 29 (2.2) | 893 (3.0) |  |
| **Disease history** |  |  |  |
| Heart failure, n (%) | 343 (25.6) | 5,217 (17.6) | 0.197 |
| CAD, n (%) | 314 (23.5) | 5,307 (17.9) | 0.139 |
| COPD, n (%) | 379 (28.3) | 5,812 (19.6) | 0.206 |
| Hyperlipidemia, n (%) | 735 (54.9) | 13,509 (45.4) | 0.190 |
| Hypertension, n (%) | 907 (67.7) | 16,903 (56.9) | 0.226 |
| Atrial fibrillation, n (%) | 299 (22.3) | 4,713 (15.9) | 0.165 |
| Type 2 DM, n (%) | 390 (29.1) | 7,146 (24.0) | 0.115 |
| **Medication** |  |  |  |
| ASA, n (%) | 550 (41.1) | 10,729 (36.1) | 0.102 |
| P2Y<sub>12</sub> Inhibitor, n (%) | 63 (4.7) | 1,346 (4.5) | 0.008 |
| Statin | 650 (48.5) | 11,785 (39.6) | 0.180 |
| ACEi, n (%) | 269 (20.1) | 6,054 (20.4) | 0.007 |
| ARB, n (%) | 193 (14.4) | 3,321 (11.1) | 0.098 |
| Beta blocker, n (%) | 584 (43.6) | 10,499 (35.3) | 0.170 |
| Loop diuretic, n (%) | 402 (30.0) | 6,035 (20.3) | 0.225 |
| MRA, n (%) | 69 (5.2) | 1,197 (4.0) | 0.054 |
| **Virus testing method** |  |  |  |
| Method |  |  | 0.030 |
| A, n (%) | 695 (51.9) | 14,990 (50.4) |  |
| B, n (%) | 644 (48.1) | 14,736 (49.6) |  |

Abbreviations. SMD: standardized mean difference, CAD: coronary heart disease, COPD: chronic obstructive pulmonary disease, DM: diabetes mellitus, ASA: acetyl-salicylic acid, ACEi: angiotensin-converting enzyme inhibitor, ARB: angiotensin II receptor blocker, MRA: mineralocorticoid receptor antagonist

**Table S12.**
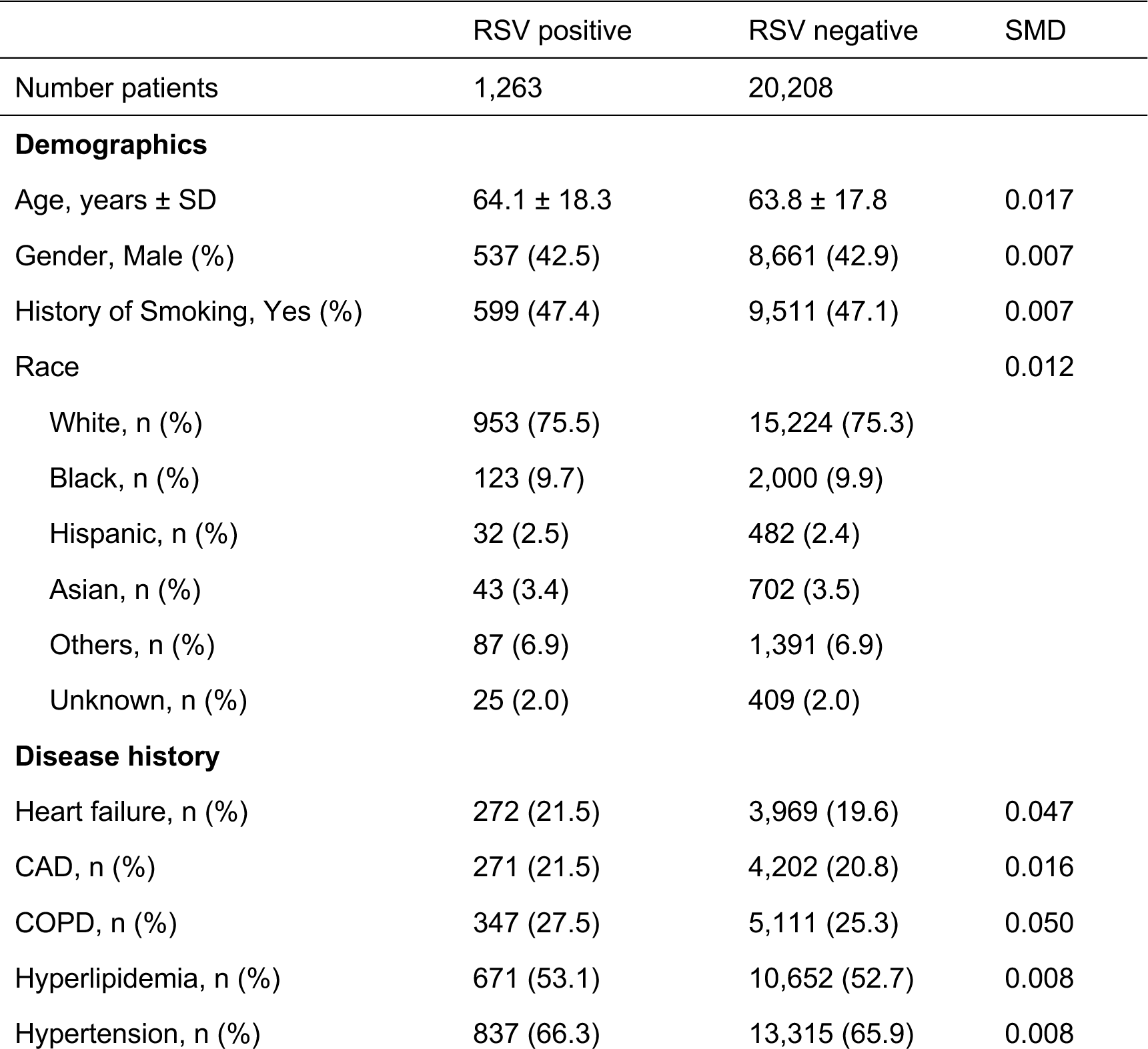
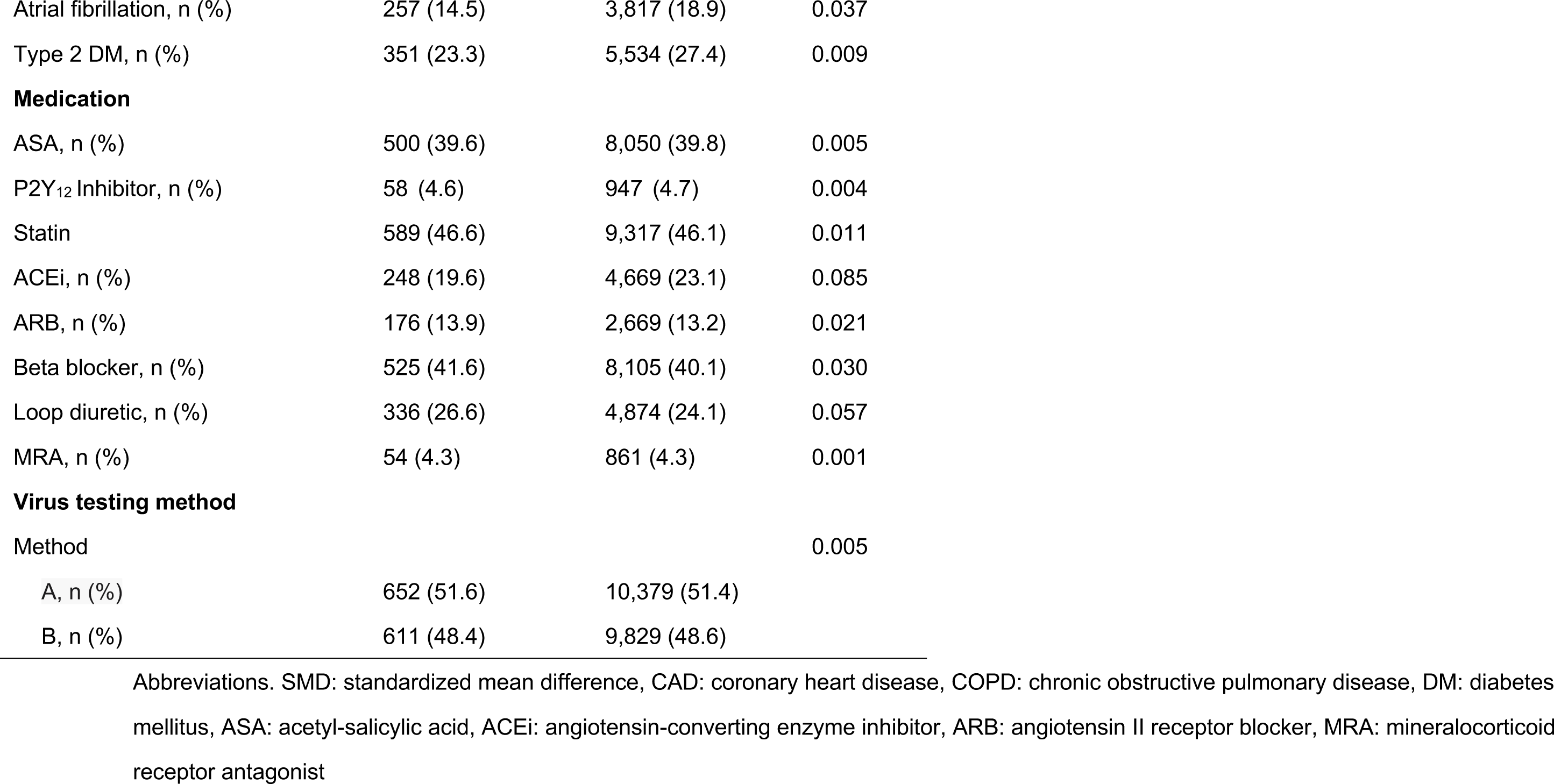
Baseline characteristics for survival analysis of heart failure stratified by RSV infection (with 90 days exclusion window, matched)

|  | RSV positive | RSV negative | SMD |
| --- | --- | --- | --- |
| Number patients | 1,263 | 20,208 |  |
| **Demographics** |  |  |  |
| Age, years ± SD | 64.1 ± 18.3 | 63.8 ± 17.8 | 0.017 |
| Gender, Male (%) | 537 (42.5) | 8,661 (42.9) | 0.007 |
| History of Smoking, Yes (%) | 599 (47.4) | 9,511 (47.1) | 0.007 |
| Race |  |  | 0.012 |
| White, n (%) | 953 (75.5) | 15,224 (75.3) |  |
| Black, n (%) | 123 (9.7) | 2,000 (9.9) |  |
| Hispanic, n (%) | 32 (2.5) | 482 (2.4) |  |
| Asian, n (%) | 43 (3.4) | 702 (3.5) |  |
| Others, n (%) | 87 (6.9) | 1,391 (6.9) |  |
| Unknown, n (%) | 25 (2.0) | 409 (2.0) |  |
| **Disease history** |  |  |  |
| Heart failure, n (%) | 272 (21.5) | 3,969 (19.6) | 0.047 |
| CAD, n (%) | 271 (21.5) | 4,202 (20.8) | 0.016 |
| COPD, n (%) | 347 (27.5) | 5,111 (25.3) | 0.050 |
| Hyperlipidemia, n (%) | 671 (53.1) | 10,652 (52.7) | 0.008 |
| Hypertension, n (%) | 837 (66.3) | 13,315 (65.9) | 0.008 |
| Atrial fibrillation, n (%) | 257 (14.5) | 3,817 (18.9) | 0.037 |
| Type 2 DM, n (%) | 351 (23.3) | 5,534 (27.4) | 0.009 |
| **Medication** |  |  |  |
| ASA, n (%) | 500 (39.6) | 8,050 (39.8) | 0.005 |
| P2Y<sub>12</sub> Inhibitor, n (%) | 58 (4.6) | 947 (4.7) | 0.004 |
| Statin | 589 (46.6) | 9,317 (46.1) | 0.011 |
| ACEi, n (%) | 248 (19.6) | 4,669 (23.1) | 0.085 |
| ARB, n (%) | 176 (13.9) | 2,669 (13.2) | 0.021 |
| Beta blocker, n (%) | 525 (41.6) | 8,105 (40.1) | 0.030 |
| Loop diuretic, n (%) | 336 (26.6) | 4,874 (24.1) | 0.057 |
| MRA, n (%) | 54 (4.3) | 861 (4.3) | 0.001 |
| **Virus testing method** |  |  |  |
| Method |  |  | 0.005 |
| A, n (%) | 652 (51.6) | 10,379 (51.4) |  |
| B, n (%) | 611 (48.4) | 9,829 (48.6) |  |

Abbreviations. SMD: standardized mean difference, CAD: coronary heart disease, COPD: chronic obstructive pulmonary disease, DM: diabetes mellitus, ASA: acetyl-salicylic acid, ACEi: angiotensin-converting enzyme inhibitor, ARB: angiotensin II receptor blocker, MRA: mineralocorticoid receptor antagonist

**Table S13.** Baseline characteristics for survival analysis of urgent revascularization stratified by RSV infection (with 90 days exclusion window, matched)

|  | RSV positive | RSV negative | SMD |
| --- | --- | --- | --- |
| Number patients | 1,340 | 21,440 |  |
| **Demographics** |  |  |  |
| Age, years ± SD | 64.6 ± 18.2 | 64.2 ± 17.7 | 0.022 |
| Gender, Male (%) | 568 (42.4) | 9,202 (42.9) | 0.011 |
| History of Smoking, Yes (%) | 645 (48.1) | 10,238 (47.8) | 0.008 |
| Race |  |  | 0.011 |
| White, n (%) | 1,008 (75.2) | 16,094 (75.1) |  |
| Black, n (%) | 134 (10.0) | 2,202 (10.3) |  |
| Hispanic, n (%) | 33 (2.5) | 517 (2.4) |  |
| Asian, n (%) | 43 (3.2) | 699 (3.3) |  |
| Others, n (%) | 93 (6.9) | 1,474 (6.9) |  |
| Unknown, n (%) | 29 (2.2) | 454 (2.1) |  |
| **Disease history** |  |  |  |
| Heart failure, n (%) | 342 (25.5) | 4,926 (23.0) | 0.059 |
| CAD, n (%) | 311 (23.2) | 4,736 (22.1) | 0.027 |
| COPD, n (%) | 381 (28.4) | 5,564 (26.0) | 0.056 |
| Hyperlipidemia, n (%) | 732 (54.6) | 11,604 (54.1) | 0.010 |
| Hypertension, n (%) | 909 (67.8) | 14,411 (67.2) | 0.013 |
| Atrial fibrillation, n (%) | 298 (22.2) | 4,378 (20.4) | 0.044 |
| Type 2 DM, n (%) | 391 (23.3) | 6,141 (28.6) | 0.012 |
| **Medication** |  |  |  |
| ASA, n (%) | 548 (40.9) | 8,804 (41.1) | 0.003 |
| P2Y<sub>12</sub> Inhibitor, n (%) | 61 (4.6) | 1,013 (4.7) | 0.008 |
| Statin | 647 (48.3) | 10,191 (47.5) | 0.015 |
| ACEi, n (%) | 271 (20.2) | 5,110 (23.8) | 0.087 |
| ARB, n (%) | 193 (14.4) | 2,905 (13.5) | 0.025 |
| Beta blocker, n (%) | 583 (43.5) | 8,977 (41.9) | 0.033 |
| Loop diuretic, n (%) | 403 (30.1) | 6,055 (20.3) | 0.226 |
| MRA, n (%) | 69 (5.1) | 1,047 (4.9) | 0.012 |
| **Virus testing method** |  |  |  |
| Method |  |  | 0.001 |
| A, n (%) | 693 (51.7) | 11,100 (51.8) |  |
| B, n (%) | 647 (48.3) | 10,340 (48.2) |  |

Abbreviations. SMD: standardized mean difference, CAD: coronary heart disease, COPD: chronic obstructive pulmonary disease, DM: diabetes mellitus, ASA: acetyl-salicylic acid, ACEi: angiotensin-converting enzyme inhibitor, ARB: angiotensin II receptor blocker, MRA: mineralocorticoid receptor antagonist

**Table S14.** Baseline characteristics for survival analysis of ischemic stroke stratified by RSV infection (with 90 days exclusion window, matched)

|  | RSV positive | RSV negative | SMD |
| --- | --- | --- | --- |
| Number patients | 1,339 | 21,424 |  |
| **Demographics** |  |  |  |
| Age, years ± SD | 64.6 ± 18.2 | 64.2 ± 17.7 | 0.024 |
| Gender, Male (%) | 569 (42.5) | 9,218 (43.0) | 0.011 |
| History of Smoking, Yes (%) | 644 (48.1) | 10,218 (47.7) | 0.008 |
| Race |  |  | 0.008 |
| White, n (%) | 1,006 (75.1) | 16,117 (75.2) |  |
| Black, n (%) | 135 (10.1) | 2,152 (10.0) |  |
| Hispanic, n (%) | 33 (2.5) | 518 (2.4) |  |
| Asian, n (%) | 43 (3.2) | 700 (3.3) |  |
| Others, n (%) | 93 (6.9) | 1,492 (7.0) |  |
| Unknown, n (%) | 29 (2.2) | 445 (2.1) |  |
| **Disease history** |  |  |  |
| Heart failure, n (%) | 343 (25.6) | 4,906 (22.9) | 0.063 |
| CAD, n (%) | 314 (23.5) | 4,813 (22.5) | 0.023 |
| COPD, n (%) | 379 (28.3) | 5,519 (25.8) | 0.057 |
| Hyperlipidemia, n (%) | 735 (54.9) | 11,668 (54.5) | 0.009 |
| Hypertension, n (%) | 907 (67.7) | 14,452 (67.5) | 0.006 |
| Atrial fibrillation, n (%) | 299 (22.3) | 4,391 (20.5) | 0.045 |
| Type 2 DM, n (%) | 390 (29.1) | 6,108 (28.5) | 0.014 |
| **Medication** |  |  |  |
| ASA, n (%) | 550 (41.1) | 8,852 (41.3) | 0.005 |
| P2Y<sub>12</sub> Inhibitor, n (%) | 63 (4.7) | 1,045 (4.9) | 0.008 |
| Statin | 650 (48.5) | 10,218 (47.7) | 0.017 |
| ACEi, n (%) | 269 (20.1) | 5,089 (23.8) | 0.089 |
| ARB, n (%) | 193 (14.4) | 2,898 (13.5) | 0.026 |
| Beta blocker, n (%) | 584 (43.6) | 9,025 (42.1) | 0.030 |
| Loop diuretic, n (%) | 402 (30.0) | 5,749 (26.8) | 0.071 |
| MRA, n (%) | 69 (5.2) | 1,045 (4.9) | 0.013 |
| **Virus testing method** |  |  |  |
| Method |  |  | 0.001 |
| A, n (%) | 695 (51.9) | 11,135 (52.0) |  |
| B, n (%) | 644 (48.1) | 10,289 (48.0) |  |

Abbreviations. SMD: standardized mean difference, CAD: coronary heart disease, COPD: chronic obstructive pulmonary disease, DM: diabetes mellitus, ASA: acetyl-salicylic acid, ACEi: angiotensin-converting enzyme inhibitor, ARB: angiotensin II receptor blocker, MRA: mineralocorticoid receptor antagonist

**Table S15.** Baseline characteristics for survival analysis of heart failure hospitalization stratified by influenza infection (unmatched)

|  | influenza positive | influenza negative | SMD |
| --- | --- | --- | --- |
| Number patients | 5138 | 53,384 |  |
| **Demographics** |  |  |  |
| Age, years ± SD | 55.2 ± 19.8 | 58.6 ± 19.5 | 0.172 |
| Gender, Male (%) | 1,867 (36.3) | 22,599 (42.3) | 0.123 |
| History of Smoking, Yes (%) | 1,720 (33.5) | 22,900 (42.9) | 0.195 |
| Race |  |  | 0.084 |
| White, n (%) | 3,702 (72.1) | 39,644 (74.3) |  |
| Black, n (%) | 424 (8.3) | 4,877 (9.1) |  |
| Hispanic, n (%) | 169 (3.3) | 1,422 (2.7) |  |
| Asian, n (%) | 220 (4.3) | 1,906 (3.6) |  |
| Others, n (%) | 420 (8.2) | 3,833 (7.2) |  |
| Unknown, n (%) | 203 (4.0) | 1,702 (3.2) |  |
| **Disease history** |  |  |  |
| Heart failure, n (%) | 425 (8.3) | 7,387 (13.8) | 0.178 |
| CAD, n (%) | 541 (10.5) | 8,086 (15.1) | 0.138 |
| COPD, n (%) | 639 (12.4) | 9,657 (18.1) | 0.158 |
| Hyperlipidemia, n (%) | 2,035 (39.6) | 22,652 (42.4) | 0.057 |
| Hypertension, n (%) | 2,179 (42.4) | 27,570 (51.6) | 0.186 |
| Atrial fibrillation, n (%) | 460 (9.0) | 7,406 (13.9) | 0.155 |
| Type 2 DM, n (%) | 864 (16.8) | 11,210 (21.0) | 0.107 |
| **Medication** |  |  |  |
| ASA, n (%) | 1,326 (25.8) | 17,379 (32.6) | 0.149 |
| P2Y<sub>12</sub> Inhibitor, n (%) | 145 (2.8) | 2,104 (3.9) | 0.062 |
| Statin | 1,577 (30.7) | 19,265 (36.1) | 0.115 |
| ACEi, n (%) | 779 (15.2) | 9,894 (18.5) | 0.090 |
| ARB, n (%) | 451 (8.8) | 5,462 (10.2) | 0.050 |
| Beta blocker, n (%) | 1137 (22.1) | 16,798 (31.5) | 0.212 |
| Loop diuretic, n (%) | 529 (10.3) | 9,263 (17.4) | 0.206 |
| MRA, n (%) | 117 (2.3) | 1,840 (3.4) | 0.070 |
| **Virus testing method** |  |  |  |
| Method |  |  | 0.724 |
| A, n (%) | 3,133 (61.0) | 17,285 (32.4) |  |
| B, n (%) | 725 (14.1) | 5,016 (9.4) |  |
| C, n (%) | 720 (14.0) | 16,196 (30.3) |  |
| D, n (%) | 560 (10.9) | 14,887 (27.9) |  |

Abbreviations. SMD: standardized mean difference, CAD: coronary heart disease, COPD: chronic obstructive pulmonary disease, DM: diabetes mellitus, ASA: acetyl-salicylic acid, ACEi: angiotensin-converting enzyme inhibitor, ARB: angiotensin II receptor blocker, MRA: mineralocorticoid receptor antagonist

**Table S16.** Baseline characteristics for survival analysis of urgent revascularization stratified by influenza infection (unmatched)

|  | influenza positive | influenza negative | SMD |
| --- | --- | --- | --- |
| Number patients | 5,176 | 54,418 |  |
| **Demographics** |  |  |  |
| Age, years ± SD | 55.4 ± 19.8 | 58.9 ± 19.6 | 0.177 |
| Gender, Male (%) | 1,886 (36.4) | 23,097 (42.4) | 0.123 |
| History of Smoking, Yes (%) | 1,743 (33.7) | 23,496 (43.2) | 0.196 |
| Race |  |  | 0.087 |
| White, n (%) | 3,733 (72.1) | 40,475 (74.4) |  |
| Black, n (%) | 425 (8.2) | 4,973 (9.1) |  |
| Hispanic, n (%) | 170 (3.3) | 1,439 (2.6) |  |
| Asian, n (%) | 224 (4.3) | 1,931 (3.5) |  |
| Others, n (%) | 420 (8.1) | 3,876 (7.1) |  |
| Unknown, n (%) | 204 (3.9) | 1,724 (3.2) |  |
| **Disease history** |  |  |  |
| Heart failure, n (%) | 457 (8.8) | 8,107 (14.9) | 0.189 |
| CAD, n (%) | 560 (10.8) | 8,481 (15.6) | 0.141 |
| COPD, n (%) | 657 (12.4) | 10,029 (18.4) | 0.159 |
| Hyperlipidemia, n (%) | 2,060 (39.8) | 23,315 (42.8) | 0.062 |
| Hypertension, n (%) | 2,211 (42.7) | 28,435 (52.3) | 0.192 |
| Atrial fibrillation, n (%) | 487 (9.4) | 7,902 (14.5) | 0.158 |
| Type 2 DM, n (%) | 881 (17.0) | 11,613 (21.3) | 0.110 |
| **Medication** |  |  |  |
| ASA, n (%) | 1,353 (26.1) | 17,945 (33.0) | 0.150 |
| P2Y<sub>12</sub> Inhibitor, n (%) | 147 (2.8) | 2,201 (4.0) | 0.066 |
| Statin | 1,604 (31.0) | 19,891 (36.6) | 0.118 |
| ACEi, n (%) | 787 (15.2) | 10,177 (18.7) | 0.093 |
| ARB, n (%) | 457 (8.8) | 5,647 (10.4) | 0.053 |
| Beta blocker, n (%) | 1,164 (22.5) | 17,478 (32.1) | 0.217 |
| Loop diuretic, n (%) | 561 (10.8) | 9,950 (18.3) | 0.212 |
| MRA, n (%) | 123 (2.4) | 1,947 (3.6) | 0.071 |
| **Virus testing method** |  |  |  |
| Method |  |  | 0.728 |
| A, n (%) | 3,152 (60.9) | 17,486 (32.1) |  |
| B, n (%) | 726 (14.0) | 5,039 (9.3) |  |
| C, n (%) | 734 (14.2) | 16,632 (30.6) |  |
| D, n (%) | 564 (10.9) | 15,261 (28.0) |  |

Abbreviations. SMD: standardized mean difference, CAD: coronary heart disease, COPD: chronic obstructive pulmonary disease, DM: diabetes mellitus, ASA: acetyl-salicylic acid, ACEi: angiotensin-converting enzyme inhibitor, ARB: angiotensin II receptor blocker, MRA: mineralocorticoid receptor antagonist

**Table S17.** Baseline characteristics for survival analysis of stroke stratified by influenza infection (unmatched)

|  | influenza positive | influenza negative | SMD |
| --- | --- | --- | --- |
| Number patients | 5,172 | 54,351 |  |
| **Demographics** |  |  |  |
| Age, years ± SD | 55.3 ± 19.8 | 58.8 ± 19.6 | 0.178 |
| Gender, Male (%) | 1,885 (36.4) | 23,077 (42.5) | 0.123 |
| History of Smoking, Yes (%) | 1,742 (33.7) | 23,479 (43.2) | 0.197 |
| Race |  |  | 0.088 |
| White, n (%) | 3,730 (72.1) | 40,434 (74.4) |  |
| Black, n (%) | 424 (8.2) | 4,959 (9.1) |  |
| Hispanic, n (%) | 170 (3.3) | 1,440 (2.6) |  |
| Asian, n (%) | 224 (4.3) | 1,924 (3.5) |  |
| Others, n (%) | 420 (8.1) | 3,875 (7.1) |  |
| Unknown, n (%) | 204 (3.9) | 1,719 (3.2) |  |
| **Disease history** |  |  |  |
| Heart failure, n (%) | 456 (8.8) | 8,095 (14.9) | 0.189 |
| CAD, n (%) | 563 (10.9) | 8,488 (15.6) | 0.140 |
| COPD, n (%) | 658 (12.4) | 10,009 (18.4) | 0.158 |
| Hyperlipidemia, n (%) | 2,059 (39.8) | 23,281 (42.8) | 0.061 |
| Hypertension, n (%) | 2,207 (42.7) | 28,385 (52.2) | 0.192 |
| Atrial fibrillation, n (%) | 485 (9.4) | 7,876 (14.5) | 0.158 |
| Type 2 DM, n (%) | 881 (17.0) | 11,589 (21.3) | 0.109 |
| **Medication** |  |  |  |
| ASA, n (%) | 1,353 (26.2) | 17,913 (33.0) | 0.149 |
| P2Y<sub>12</sub> Inhibitor, n (%) | 147 (2.8) | 2,191 (4.0) | 0.065 |
| Statin | 1,601 (31.0) | 19,855 (36.5) | 0.118 |
| ACEi, n (%) | 787 (15.2) | 10,151 (18.7) | 0.092 |
| ARB, n (%) | 456 (8.8) | 5,639 (10.4) | 0.053 |
| Beta blocker, n (%) | 1,167 (22.6) | 17,443 (32.1) | 0.215 |
| Loop diuretic, n (%) | 559 (10.8) | 9,934 (18.3) | 0.213 |
| MRA, n (%) | 122 (2.4) | 1,951 (3.6) | 0.072 |
| **Virus testing method** |  |  |  |
| Method |  |  | 0.727 |
| A, n (%) | 3,149 (60.9) | 17,475 (32.2) |  |
| B, n (%) | 725 (14.0) | 5,038 (9.3) |  |
| C, n (%) | 733 (14.2) | 16,602 (30.5) |  |
| D, n (%) | 565 (10.9) | 15,236 (28.0) |  |

Abbreviations. SMD: standardized mean difference, CAD: coronary heart disease, COPD: chronic obstructive pulmonary disease, DM: diabetes mellitus, ASA: acetyl-salicylic acid, ACEi: angiotensin-converting enzyme inhibitor, ARB: angiotensin II receptor blocker, MRA: mineralocorticoid receptor antagonist

**Table S18.** Baseline characteristics for survival analysis of heart failure hospitalization stratified by influenza infection (matched)

|  | influenza positive | influenza negative | SMD |
| --- | --- | --- | --- |
| Number patients | 5,138 | 25,690 |  |
| **Demographics** |  |  |  |
| Age, years ± SD | 55.2 ± 19.8 | 55.1 ± 19.6 | 0.007 |
| Gender, Male (%) | 1,867 (36.3) | 9,313 (36.3) | 0.002 |
| History of Smoking, Yes (%) | 1,720 (33.5) | 8,674 (33.8) | 0.006 |
| Race |  |  | 0.021 |
| White, n (%) | 3,702 (72.1) | 18,559 (72.2) |  |
| Black, n (%) | 424 (8.3) | 2,167 (8.4) |  |
| Hispanic, n (%) | 169 (3.3) | 844 (3.3) |  |
| Asian, n (%) | 220 (4.3) | 1,049 (4.1) |  |
| Others, n (%) | 420 (8.2) | 2,137 (8.3) |  |
| Unknown, n (%) | 203 (4.0) | 934 (3.6) |  |
| **Disease history** |  |  |  |
| Heart failure, n (%) | 425 (8.3) | 2,159 (8.4) | 0.005 |
| CAD, n (%) | 541 (10.5) | 2,719 (10.6) | 0.002 |
| COPD, n (%) | 639 (12.4) | 3,268 (12.7) | 0.009 |
| Hyperlipidemia, n (%) | 2,035 (39.6) | 10,036 (39.1) | 0.011 |
| Hypertension, n (%) | 2,179 (42.4) | 10,902 (42.4) | 0.001 |
| Atrial fibrillation, n (%) | 460 (9.0) | 2,319 (9.0) | 0.003 |
| Type 2 DM, n (%) | 864 (16.8) | 4,376 (17.0) | 0.006 |
| **Medication** |  |  |  |
| ASA, n (%) | 1,326 (25.8) | 6,608 (25.7) | 0.002 |
| P2Y<sub>12</sub> Inhibitor, n (%) | 145 (2.8) | 708 (2.8) | 0.004 |
| Statin | 1,577 (30.7) | 7,562 (29.4) | 0.027 |
| ACEi, n (%) | 779 (15.2) | 3,931 (15.3) | 0.004 |
| ARB, n (%) | 451 (8.8) | 2,177 (8.5) | 0.011 |
| Beta blocker, n (%) | 1137 (22.1) | 6,151 (23.9) | 0.043 |
| Loop diuretic, n (%) | 529 (10.3) | 2,729 (10.3) | 0.011 |
| MRA, n (%) | 117 (2.3) | 653 (2.5) | 0.017 |
| **Virus testing method** |  |  |  |
| Method |  |  | 0.028 |
| A, n (%) | 3,133 (61.0) | 15,478 (60.2) |  |
| B, n (%) | 725 (14.1) | 3,801 (14.8) |  |
| C, n (%) | 720 (14.0) | 3,729 (14.5) |  |
| D, n (%) | 560 (10.9) | 2,682 (10.4) |  |

Abbreviations. SMD: standardized mean difference, CAD: coronary heart disease, COPD: chronic obstructive pulmonary disease, DM: diabetes mellitus, ASA: acetyl-salicylic acid, ACEi: angiotensin-converting enzyme inhibitor, ARB: angiotensin II receptor blocker, MRA: mineralocorticoid receptor antagonist

**Table S19.** Baseline characteristics for survival analysis of urgent revascularization stratified by influenza infection (matched)

|  | influenza positive | influenza negative | SMD |
| --- | --- | --- | --- |
| Number patients | 5,176 | 25,880 |  |
| **Demographics** |  |  |  |
| Age, years ± SD | 55.4 ± 19.8 | 55.1 ± 19.7 | 0.013 |
| Gender, Male (%) | 1,886 (36.4) | 9,522 (36.8) | 0.007 |
| History of Smoking, Yes (%) | 1,743 (33.7) | 8,776 (33.9) | 0.005 |
| Race |  |  | 0.017 |
| White, n (%) | 3,733 (72.1) | 18,703 (72.3) |  |
| Black, n (%) | 425 (8.2) | 2,111 (8.2) |  |
| Hispanic, n (%) | 170 (3.3) | 834 (3.2) |  |
| Asian, n (%) | 224 (4.3) | 1,076 (4.2) |  |
| Others, n (%) | 420 (8.1) | 2,184 (8.4) |  |
| Unknown, n (%) | 204 (3.9) | 972 (3.8) |  |
| **Disease history** |  |  |  |
| Heart failure, n (%) | 457 (8.8) | 2,245 (8.7) | 0.005 |
| CAD, n (%) | 560 (10.8) | 2,782 (10.7) | 0.002 |
| COPD, n (%) | 657 (12.4) | 3,320 (12.8) | 0.004 |
| Hyperlipidemia, n (%) | 2,060 (39.8) | 10,073 (38.9) | 0.018 |
| Hypertension, n (%) | 2,211 (42.7) | 11,017 (42.6) | 0.003 |
| Atrial fibrillation, n (%) | 487 (9.4) | 2,499 (9.7) | 0.008 |
| Type 2 DM, n (%) | 881 (17.0) | 4,386 (16.9) | 0.002 |
| **Medication** |  |  |  |
| ASA, n (%) | 1,353 (26.1) | 6,792 (26.2) | 0.002 |
| P2Y<sub>12</sub> Inhibitor, n (%) | 147 (2.8) | 721 (2.8) | 0.003 |
| Statin | 1,604 (31.0) | 7,740 (29.9) | 0.024 |
| ACEi, n (%) | 787 (15.2) | 3,920 (15.1) | 0.002 |
| ARB, n (%) | 457 (8.8) | 2,232 (8.6) | 0.007 |
| Beta blocker, n (%) | 1,164 (22.5) | 6,208 (24.0) | 0.036 |
| Loop diuretic, n (%) | 561 (10.8) | 2,780 (10.7) | 0.003 |
| MRA, n (%) | 123 (2.4) | 643 (2.5) | 0.007 |
| **Virus testing method** |  |  |  |
| Method |  |  | 0.027 |
| A, n (%) | 3,152 (60.9) | 15,543 (60.1) |  |
| B, n (%) | 726 (14.0) | 3,835 (14.8) |  |
| C, n (%) | 734 (14.2) | 3,757 (14.5) |  |
| D, n (%) | 564 (10.9) | 2,745 (10.6) |  |

Abbreviations. SMD: standardized mean difference, CAD: coronary heart disease, COPD: chronic obstructive pulmonary disease, DM: diabetes mellitus, ASA: acetyl-salicylic acid, ACEi: angiotensin-converting enzyme inhibitor, ARB: angiotensin II receptor blocker, MRA: mineralocorticoid receptor antagonist

**Table S20.**
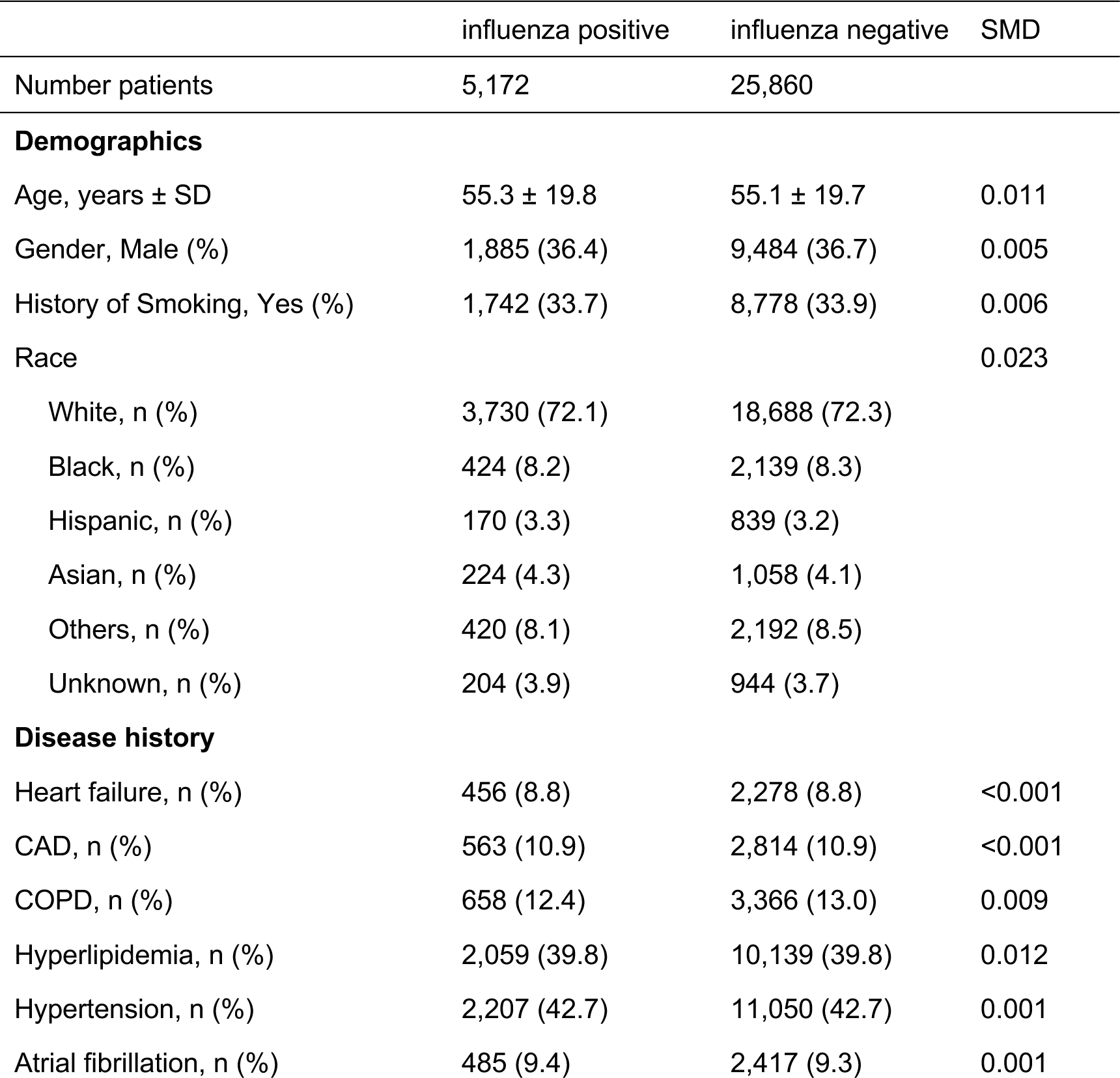
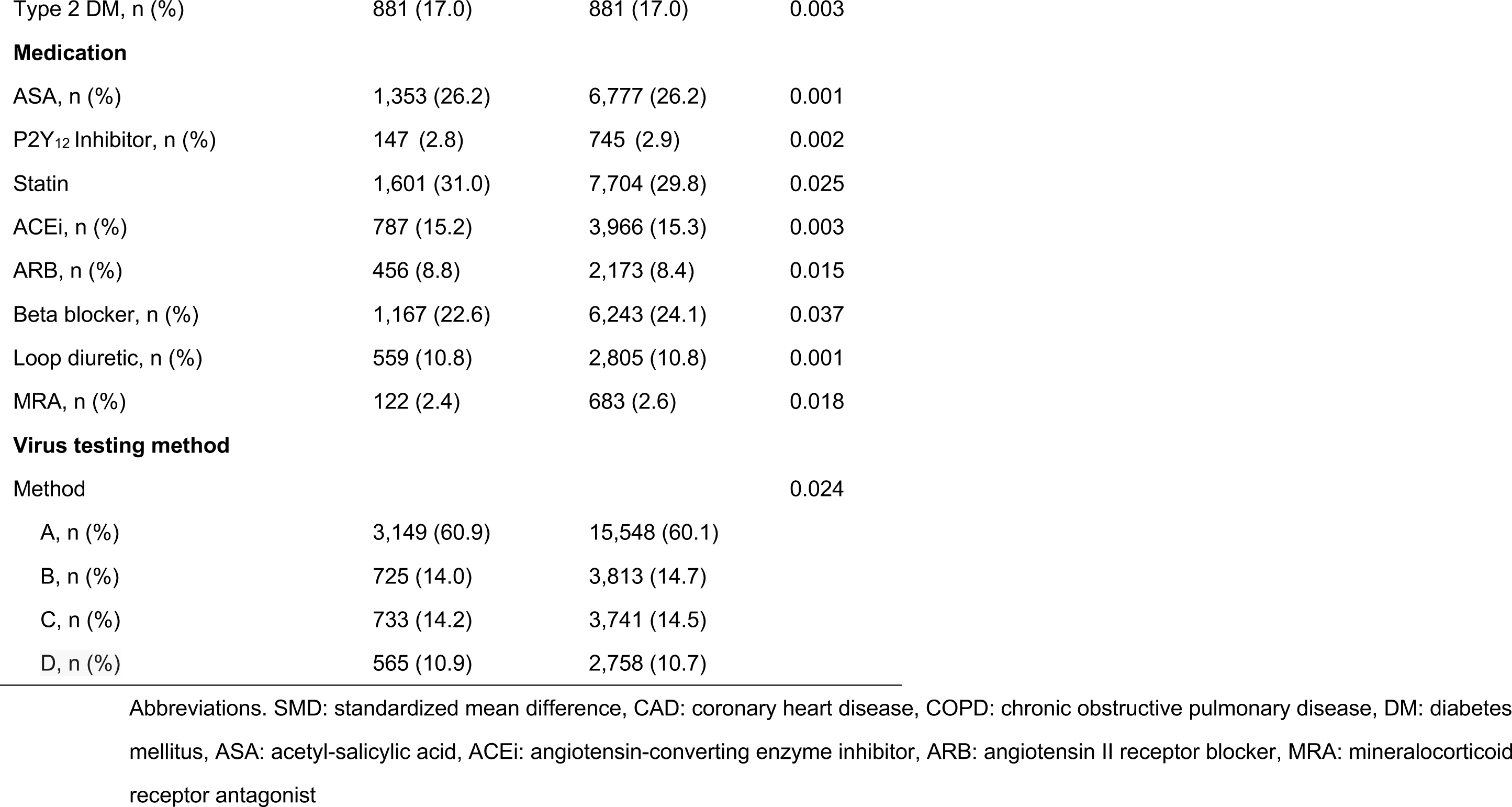
Baseline characteristics for survival analysis of stroke stratified by influenza infection (matched)

**Table S21.** Baseline characteristics for survival analysis of heart failure stratified by influenza infection (with 90 days exclusion window, unmatched)

|  | influenza positive | influenza negative | SMD |
| --- | --- | --- | --- |
| Number patients | 4,877 | 46,856 |  |
| **Demographics** |  |  |  |
| Age, years ± SD | 55.1 ± 19.6 | 58.0 ± 19.4 | 0.145 |
| Gender, Male (%) | 1,760 (36.1) | 19,533 (41.7) | 0.115 |
| History of Smoking, Yes (%) | 1,626 (33.3) | 19,878 (42.4) | 0.188 |
| Race |  |  | 0.078 |
| White, n (%) | 3,517 (72.1) | 34,621 (73.9) |  |
| Black, n (%) | 406 (8.3) | 4,362 (9.3) |  |
| Hispanic, n (%) | 157 (3.2) | 1,312 (2.8) |  |
| Asian, n (%) | 209 (4.3) | 1,675 (3.6) |  |
| Others, n (%) | 396 (8.1) | 3,421 (7.3) |  |
| Unknown, n (%) | 192 (3.9) | 1,465 (3.1) |  |
| **Disease history** |  |  |  |
| Heart failure, n (%) | 391 (8.0) | 6,118 (13.1) | 0.165 |
| CAD, n (%) | 527 (10.8) | 7,003 (14.9) | 0.124 |
| COPD, n (%) | 628 (12.9) | 8,535 (18.2) | 0.148 |
| Hyperlipidemia, n (%) | 1,984 (40.7) | 20,076 (42.8) | 0.044 |
| Hypertension, n (%) | 2,094 (42.9) | 24,185 (51.6) | 0.175 |
| Atrial fibrillation, n (%) | 432 (8.9) | 6,248 (13.3) | 0.143 |
| Type 2 DM, n (%) | 822 (16.9) | 9,727 (20.8) | 0.100 |
| **Medication** |  |  |  |
| ASA, n (%) | 1,269 (26.0) | 15,062 (32.1) | 0.135 |
| P2Y<sub>12</sub> Inhibitor, n (%) | 137 (2.8) | 1,756 (3.7) | 0.053 |
| Statin | 1,506 (30.9) | 16,554 (35.3) | 0.095 |
| ACEi, n (%) | 744 (15.3) | 8,642 (18.4) | 0.085 |
| ARB, n (%) | 429 (8.8) | 4,723 (10.1) | 0.044 |
| Beta blocker, n (%) | 1,067 (21.9) | 14,247 (30.4) | 0.195 |
| Loop diuretic, n (%) | 478 (9.8) | 7,450 (15.9) | 0.183 |
| MRA, n (%) | 104 (2.1) | 1,490 (3.2) | 0.065 |
| **Virus testing method** |  |  |  |
| Method |  |  | 0.694 |
| A, n (%) | 3,027 (62.1) | 16,119 (34.4) |  |
| B, n (%) | 696 (14.3) | 4,748 (10.1) |  |
| C, n (%) | 654 (13.4) | 13,819 (29.5) |  |
| D, n (%) | 500 (10.3) | 12,170 (26.0) |  |

Abbreviations. SMD: standardized mean difference, CAD: coronary heart disease, COPD: chronic obstructive pulmonary disease, DM: diabetes mellitus, ASA: acetyl-salicylic acid, ACEi: angiotensin-converting enzyme inhibitor, ARB: angiotensin II receptor blocker, MRA: mineralocorticoid receptor antagonist

**Table S22.**
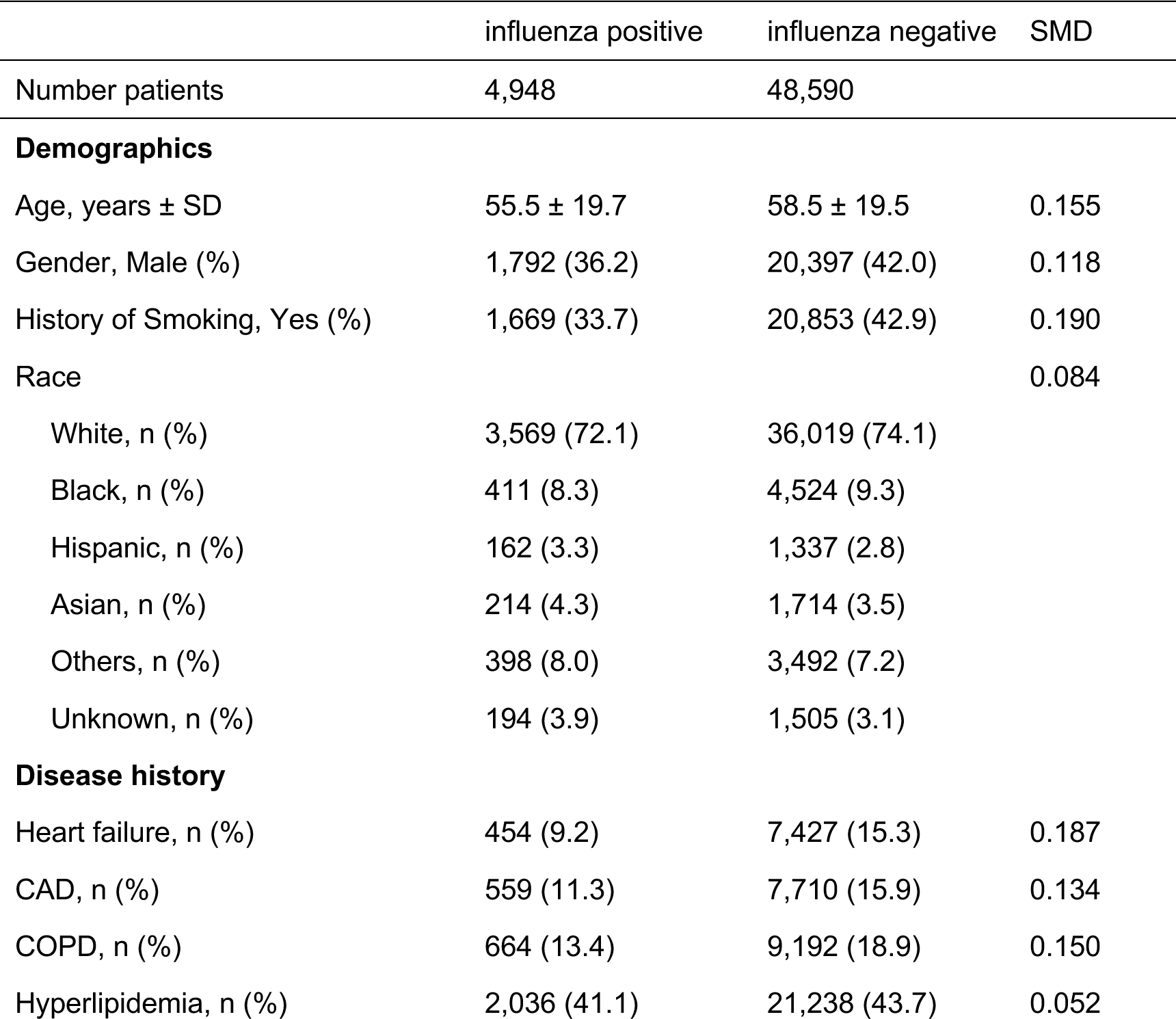
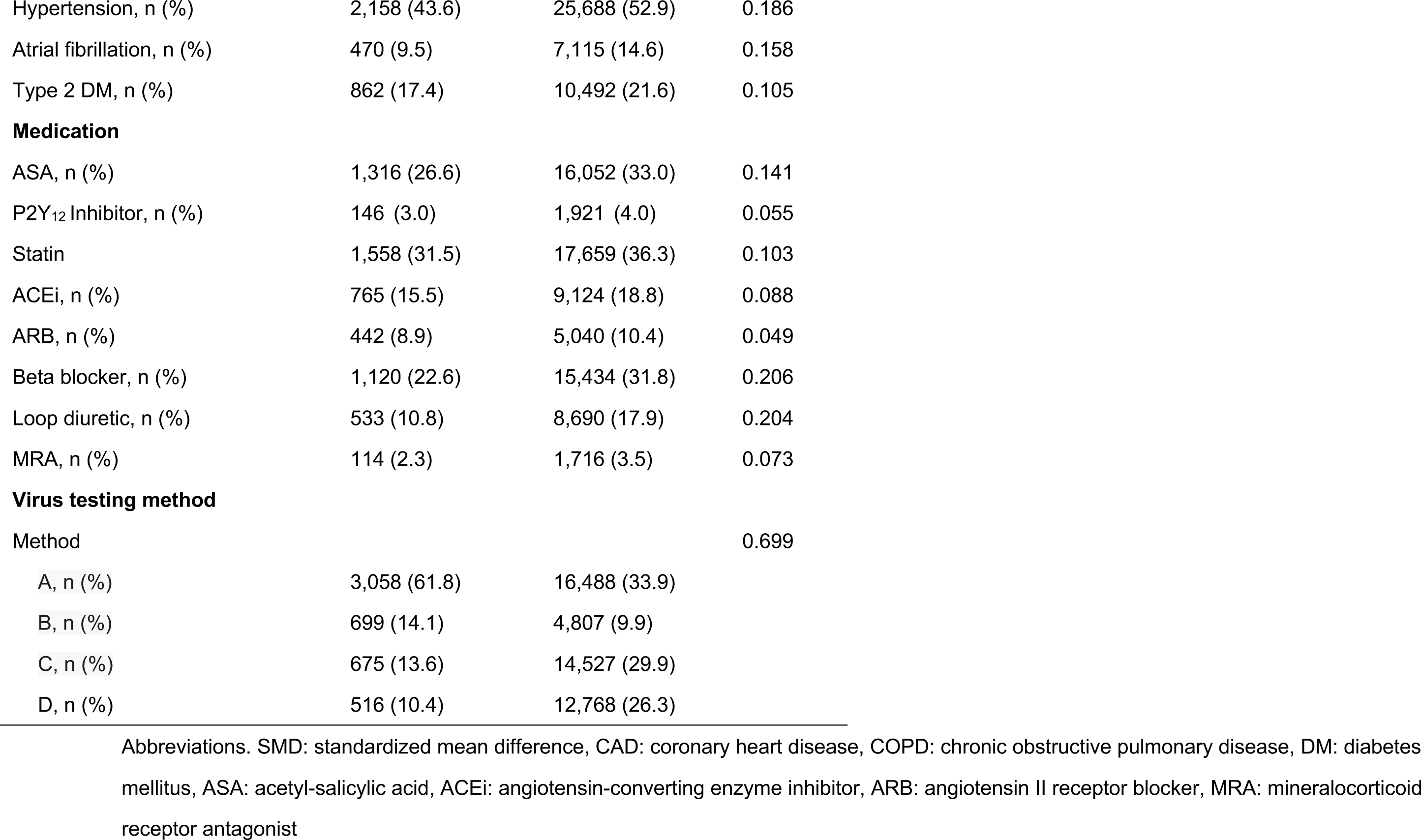
Baseline characteristics for survival analysis of urgent revascularization stratified by influenza infection (with 90 days exclusion window, unmatched)

**Table S23.** Baseline characteristics for survival analysis of ischemic stroke stratified by influenza infection (with 90 days exclusion window, unmatched)

|  | influenza positive | influenza negative | SMD |
| --- | --- | --- | --- |
| Number patients | 4,943 | 48,492 |  |
| **Demographics** |  |  |  |
| Age, years ± SD | 55.4 ± 19.6 | 58.5 ± 19.5 | 0.155 |
| Gender, Male (%) | 1,791 (36.2) | 20,359 (42.0) | 0.118 |
| History of Smoking, Yes (%) | 1,666 (33.7) | 20,814 (42.9) | 0.190 |
| Race |  |  | 0.084 |
| White, n (%) | 3,566 (72.1) | 35,956 (74.1) |  |
| Black, n (%) | 410 (8.3) | 4,500 (9.3) |  |
| Hispanic, n (%) | 162 (3.3) | 1,338 (2.8) |  |
| Asian, n (%) | 214 (4.3) | 1,705 (3.5) |  |
| Others, n (%) | 397 (8.0) | 3,492 (7.2) |  |
| Unknown, n (%) | 194 (3.9) | 1,501 (3.1) |  |
| **Disease history** |  |  |  |
| Heart failure, n (%) | 453 (9.2) | 7,398 (15.3) | 0.187 |
| CAD, n (%) | 563 (11.4) | 7,723 (15.9) | 0.132 |
| COPD, n (%) | 663 (13.4) | 9,161 (18.9) | 0.149 |
| Hyperlipidemia, n (%) | 2,035 (41.2) | 21,192 (43.7) | 0.051 |
| Hypertension, n (%) | 2,153 (43.6) | 25,612 (52.8) | 0.186 |
| Atrial fibrillation, n (%) | 471 (9.5) | 7,072 (14.6) | 0.156 |
| Type 2 DM, n (%) | 861 (17.4) | 10,443 (21.5) | 0.104 |
| **Medication** |  |  |  |
| ASA, n (%) | 1,314 (26.6) | 16,009 (33.0) | 0.141 |
| P2Y<sub>12</sub> Inhibitor, n (%) | 146 (3.0) | 1,912 (3.9) | 0.054 |
| Statin | 1,555 (31.5) | 17,608 (36.3) | 0.103 |
| ACEi, n (%) | 761 (15.4) | 9,091 (18.7) | 0.089 |
| ARB, n (%) | 443 (9.0) | 5,028 (10.4) | 0.048 |
| Beta blocker, n (%) | 1,120 (22.7) | 15,381 (31.7) | 0.205 |
| Loop diuretic, n (%) | 531 (10.7) | 8,649 (17.8) | 0.204 |
| MRA, n (%) | 113 (2.3) | 1,711 (3.5) | 0.074 |
| **Virus testing method** |  |  |  |
| Method |  |  | 0.698 |
| A, n (%) | 3,054 (61.8) | 16,465 (34.0) |  |
| B, n (%) | 699 (14.1) | 4,800 (9.9) |  |
| C, n (%) | 673 (13.6) | 14,493 (29.9) |  |
| D, n (%) | 517 (10.5) | 12,734 (26.3) |  |

Abbreviations. SMD: standardized mean difference, CAD: coronary heart disease, COPD: chronic obstructive pulmonary disease, DM: diabetes mellitus, ASA: acetyl-salicylic acid, ACEi: angiotensin-converting enzyme inhibitor, ARB: angiotensin II receptor blocker, MRA: mineralocorticoid receptor antagonist

**Table S24.** Baseline characteristics for survival analysis of heart failure stratified by influenza infection (with 90 days exclusion window, matched)

|  | influenza positive | influenza negative | SMD |
| --- | --- | --- | --- |
| Number patients | 4,877 | 24,385 |  |
| **Demographics** |  |  |  |
| Age, years ± SD | 55.1 ± 19.6 | 55.2 ± 19.5 | 0.001 |
| Gender, Male (%) | 1,760 (36.1) | 8,952 (36.7) | 0.013 |
| History of Smoking, Yes (%) | 1,626 (33.3) | 8,426 (34.6) | 0.026 |
| Race |  |  | 0.022 |
| White, n (%) | 3,517 (72.1) | 17,653 (72.4) |  |
| Black, n (%) | 406 (8.3) | 2,093 (8.6) |  |
| Hispanic, n (%) | 157 (3.2) | 764 (3.1) |  |
| Asian, n (%) | 209 (4.3) | 983 (4.0) |  |
| Others, n (%) | 396 (8.1) | 2,001 (8.2) |  |
| Unknown, n (%) | 192 (3.9) | 891 (3.7) |  |
| **Disease history** |  |  |  |
| Heart failure, n (%) | 391 (8.0) | 2,029 (8.3) | 0.011 |
| CAD, n (%) | 527 (10.8) | 2,691 (11.0) | 0.007 |
| COPD, n (%) | 628 (12.9) | 3,176 (13.0) | 0.004 |
| Hyperlipidemia, n (%) | 1,984 (40.7) | 9,774 (40.1) | 0.012 |
| Hypertension, n (%) | 2,094 (42.9) | 10,610 (43.5) | 0.012 |
| Atrial fibrillation, n (%) | 432 (8.9) | 2,215 (9.1) | 0.008 |
| Type 2 DM, n (%) | 822 (16.9) | 4,166 (17.1) | 0.006 |
| **Medication** |  |  |  |
| ASA, n (%) | 1,269 (26.0) | 6,409 (26.3) | 0.006 |
| P2Y<sub>12</sub> Inhibitor, n (%) | 137 (2.8) | 672 (2.8) | 0.003 |
| Statin | 1,506 (30.9) | 7,249 (29.7) | 0.025 |
| ACEi, n (%) | 744 (15.3) | 3,748 (15.4) | 0.003 |
| ARB, n (%) | 429 (8.8) | 2,132 (8.7) | 0.002 |
| Beta blocker, n (%) | 1,067 (21.9) | 5,853 (24.0) | 0.051 |
| Loop diuretic, n (%) | 478 (9.8) | 2,434 (10.0) | 0.006 |
| MRA, n (%) | 104 (2.1) | 600 (2.5) | 0.022 |
| **Virus testing method** |  |  |  |
| Method |  |  | 0.023 |
| A, n (%) | 3,027 (62.1) | 14,941 (61.3) |  |
| B, n (%) | 696 (14.3) | 3,662 (15.0) |  |
| C, n (%) | 654 (13.4) | 3,321 (13.6) |  |
| D, n (%) | 500 (10.3) | 2,461 (10.1) |  |

Abbreviations. SMD: standardized mean difference, CAD: coronary heart disease, COPD: chronic obstructive pulmonary disease, DM: diabetes mellitus, ASA: acetyl-salicylic acid, ACEi: angiotensin-converting enzyme inhibitor, ARB: angiotensin II receptor blocker, MRA: mineralocorticoid receptor antagonist

**Table S25.**
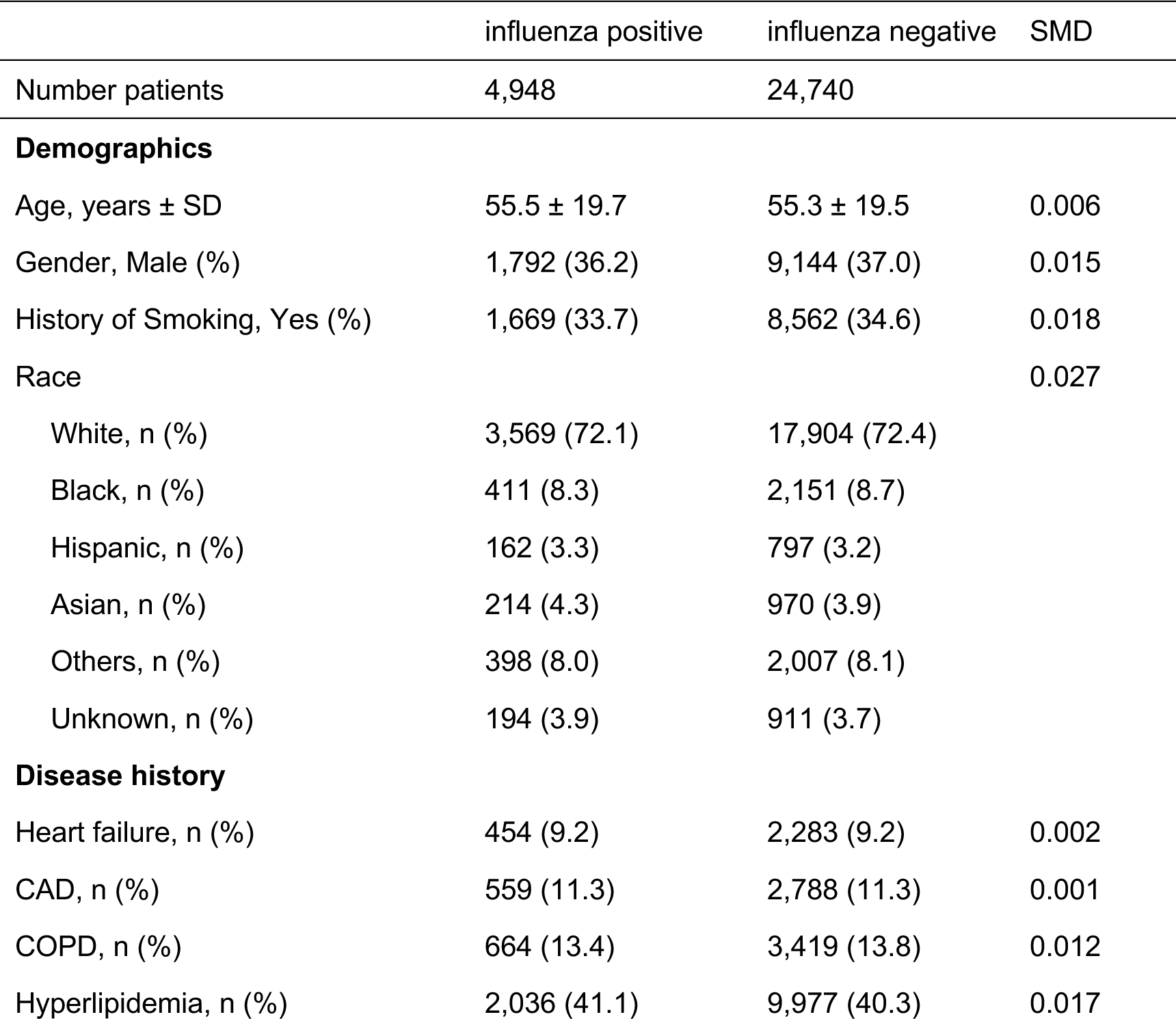
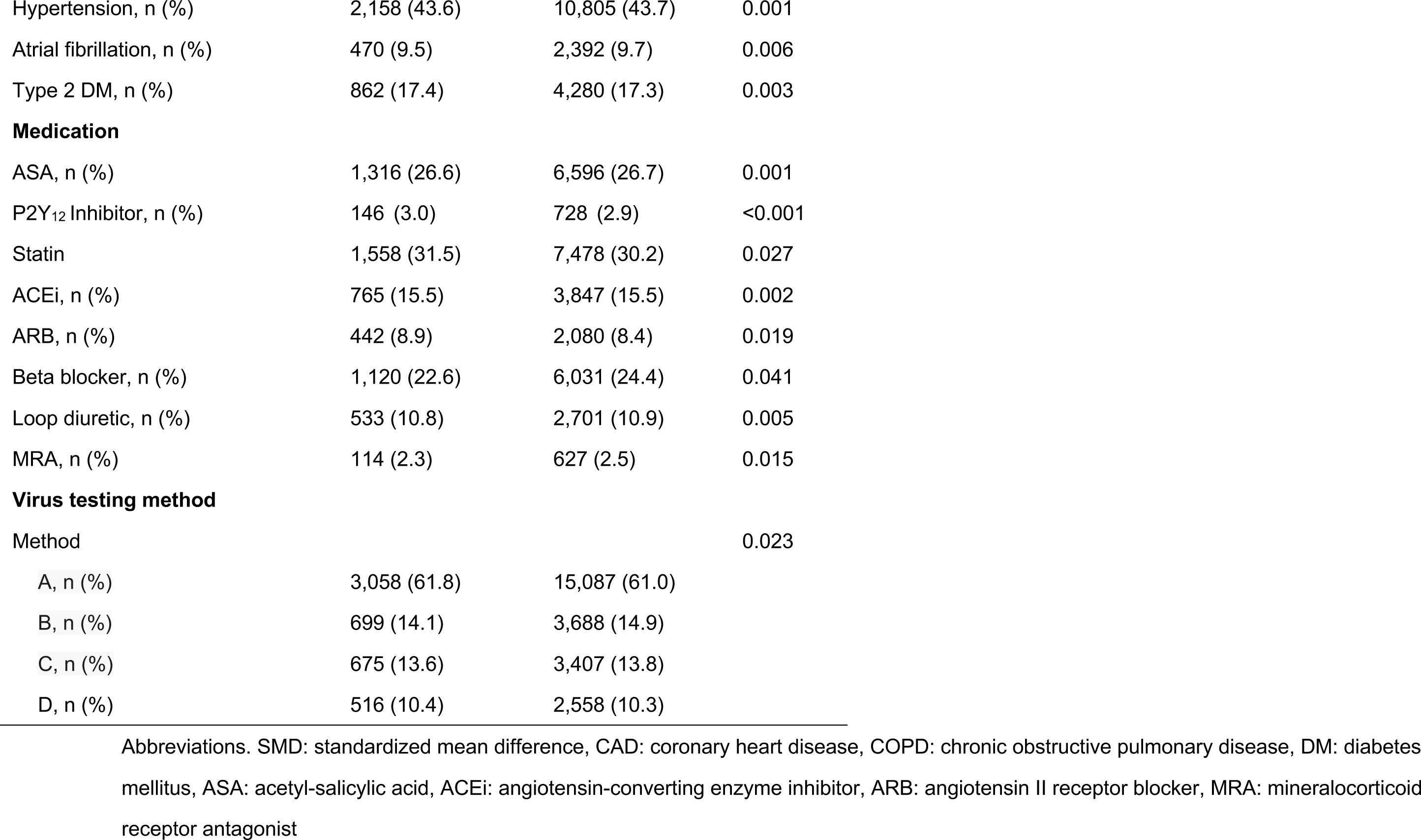
Baseline characteristics for survival analysis of urgent revascularization stratified by influenza infection (with 90 days exclusion window, matched)

|  | influenza positive | influenza negative | SMD |
| --- | --- | --- | --- |
| Number patients | 4,948 | 24,740 |  |
| **Demographics** |  |  |  |
| Age, years ± SD | 55.5 ± 19.7 | 55.3 ± 19.5 | 0.006 |
| Gender, Male (%) | 1,792 (36.2) | 9,144 (37.0) | 0.015 |
| History of Smoking, Yes (%) | 1,669 (33.7) | 8,562 (34.6) | 0.018 |
| Race |  |  | 0.027 |
| White, n (%) | 3,569 (72.1) | 17,904 (72.4) |  |
| Black, n (%) | 411 (8.3) | 2,151 (8.7) |  |
| Hispanic, n (%) | 162 (3.3) | 797 (3.2) |  |
| Asian, n (%) | 214 (4.3) | 970 (3.9) |  |
| Others, n (%) | 398 (8.0) | 2,007 (8.1) |  |
| Unknown, n (%) | 194 (3.9) | 911 (3.7) |  |
| **Disease history** |  |  |  |
| Heart failure, n (%) | 454 (9.2) | 2,283 (9.2) | 0.002 |
| CAD, n (%) | 559 (11.3) | 2,788 (11.3) | 0.001 |
| COPD, n (%) | 664 (13.4) | 3,419 (13.8) | 0.012 |
| Hyperlipidemia, n (%) | 2,036 (41.1) | 9,977 (40.3) | 0.017 |
| Hypertension, n (%) | 2,158 (43.6) | 10,805 (43.7) | 0.001 |
| Atrial fibrillation, n (%) | 470 (9.5) | 2,392 (9.7) | 0.006 |
| Type 2 DM, n (%) | 862 (17.4) | 4,280 (17.3) | 0.003 |
| **Medication** |  |  |  |
| ASA, n (%) | 1,316 (26.6) | 6,596 (26.7) | 0.001 |
| P2Y<sub>12</sub> Inhibitor, n (%) | 146 (3.0) | 728 (2.9) | <0.001 |
| Statin | 1,558 (31.5) | 7,478 (30.2) | 0.027 |
| ACEi, n (%) | 765 (15.5) | 3,847 (15.5) | 0.002 |
| ARB, n (%) | 442 (8.9) | 2,080 (8.4) | 0.019 |
| Beta blocker, n (%) | 1,120 (22.6) | 6,031 (24.4) | 0.041 |
| Loop diuretic, n (%) | 533 (10.8) | 2,701 (10.9) | 0.005 |
| MRA, n (%) | 114 (2.3) | 627 (2.5) | 0.015 |
| **Virus testing method** |  |  |  |
| Method |  |  | 0.023 |
| A, n (%) | 3,058 (61.8) | 15,087 (61.0) |  |
| B, n (%) | 699 (14.1) | 3,688 (14.9) |  |
| C, n (%) | 675 (13.6) | 3,407 (13.8) |  |
| D, n (%) | 516 (10.4) | 2,558 (10.3) |  |

Abbreviations. SMD: standardized mean difference, CAD: coronary heart disease, COPD: chronic obstructive pulmonary disease, DM: diabetes mellitus, ASA: acetyl-salicylic acid, ACEi: angiotensin-converting enzyme inhibitor, ARB: angiotensin II receptor blocker, MRA: mineralocorticoid receptor antagonist

**Table S26.**
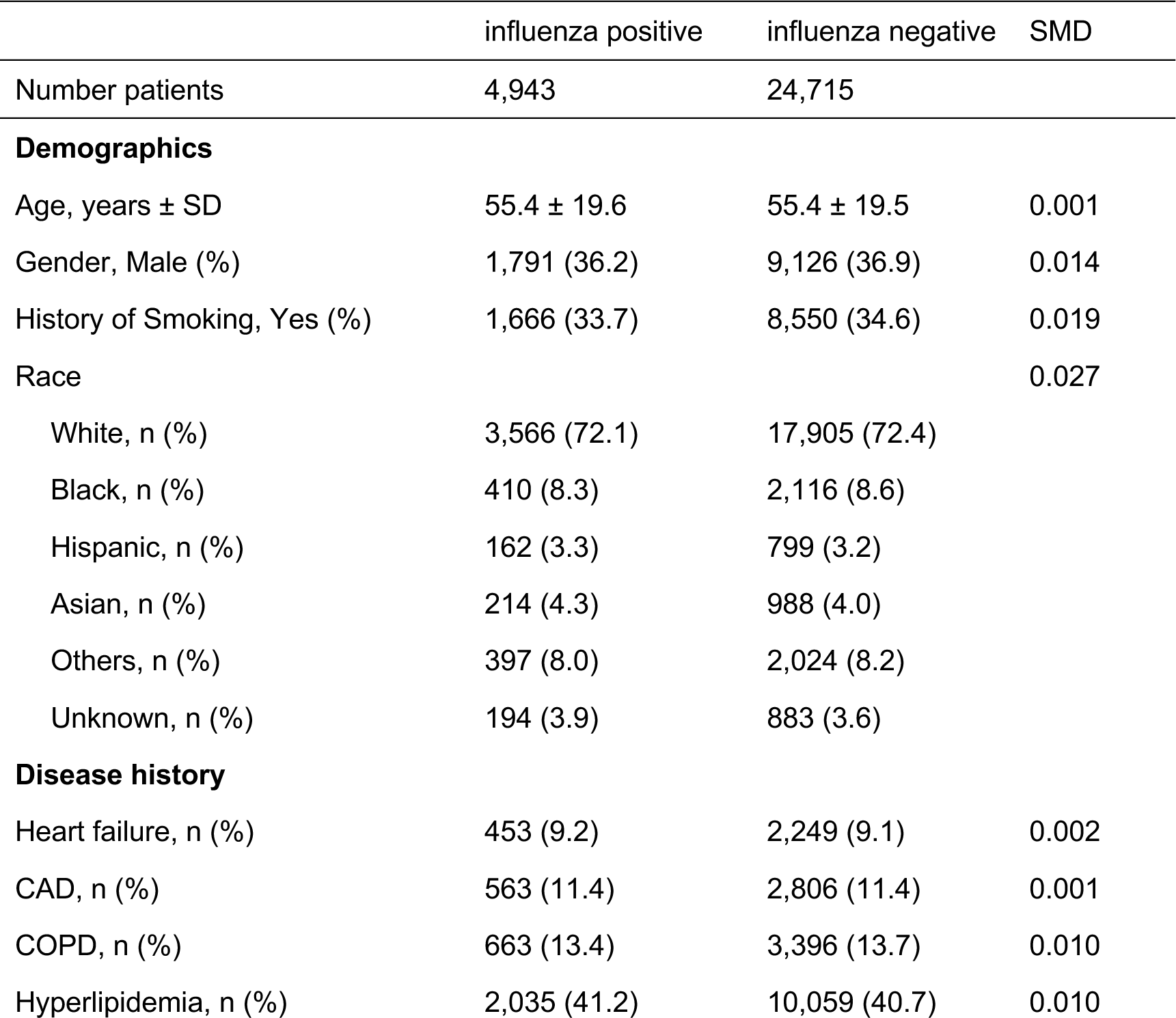
Baseline characteristics for survival analysis of ischemic stroke stratified by influenza infection (with 90 days exclusion window, matched)

**Table S26.**
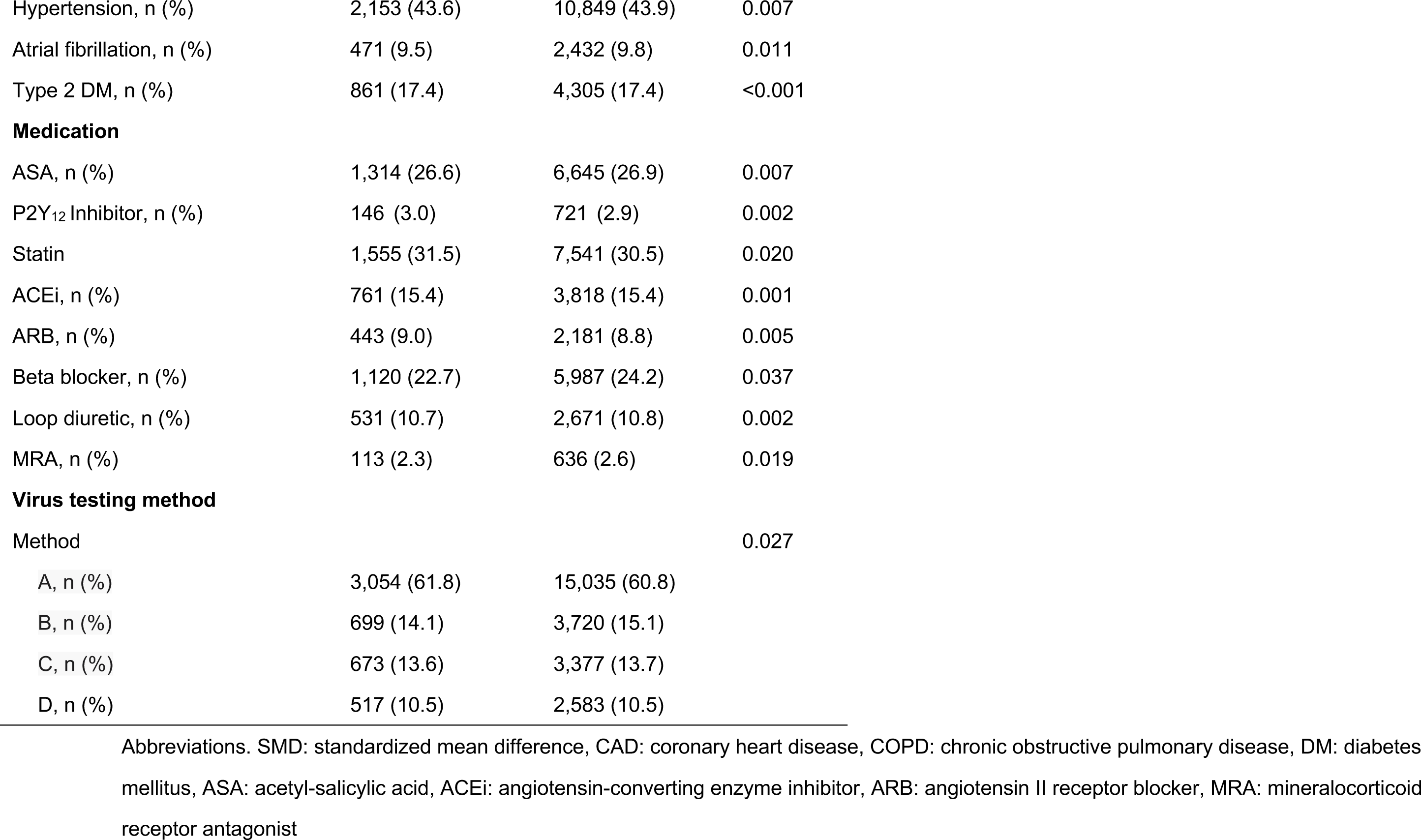
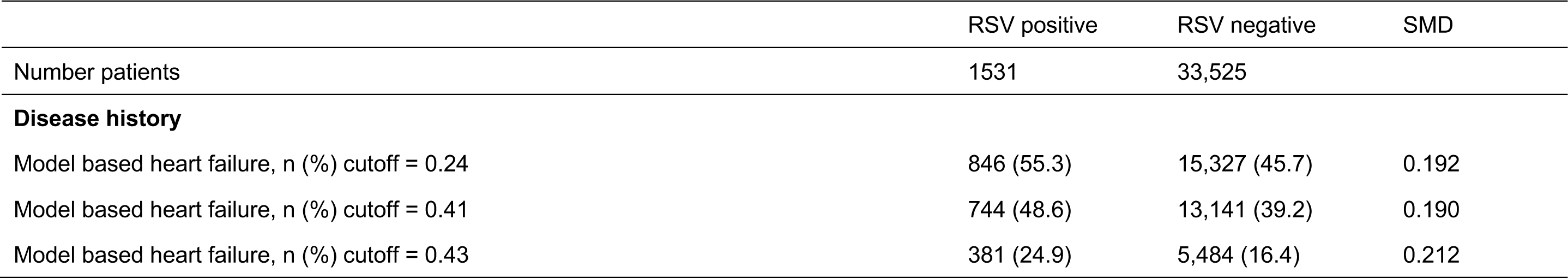
History of heart failure based on the model with various cutoffs for heart failure hospitalization analysis on RSV infection (unmatched)

**Table S27.**
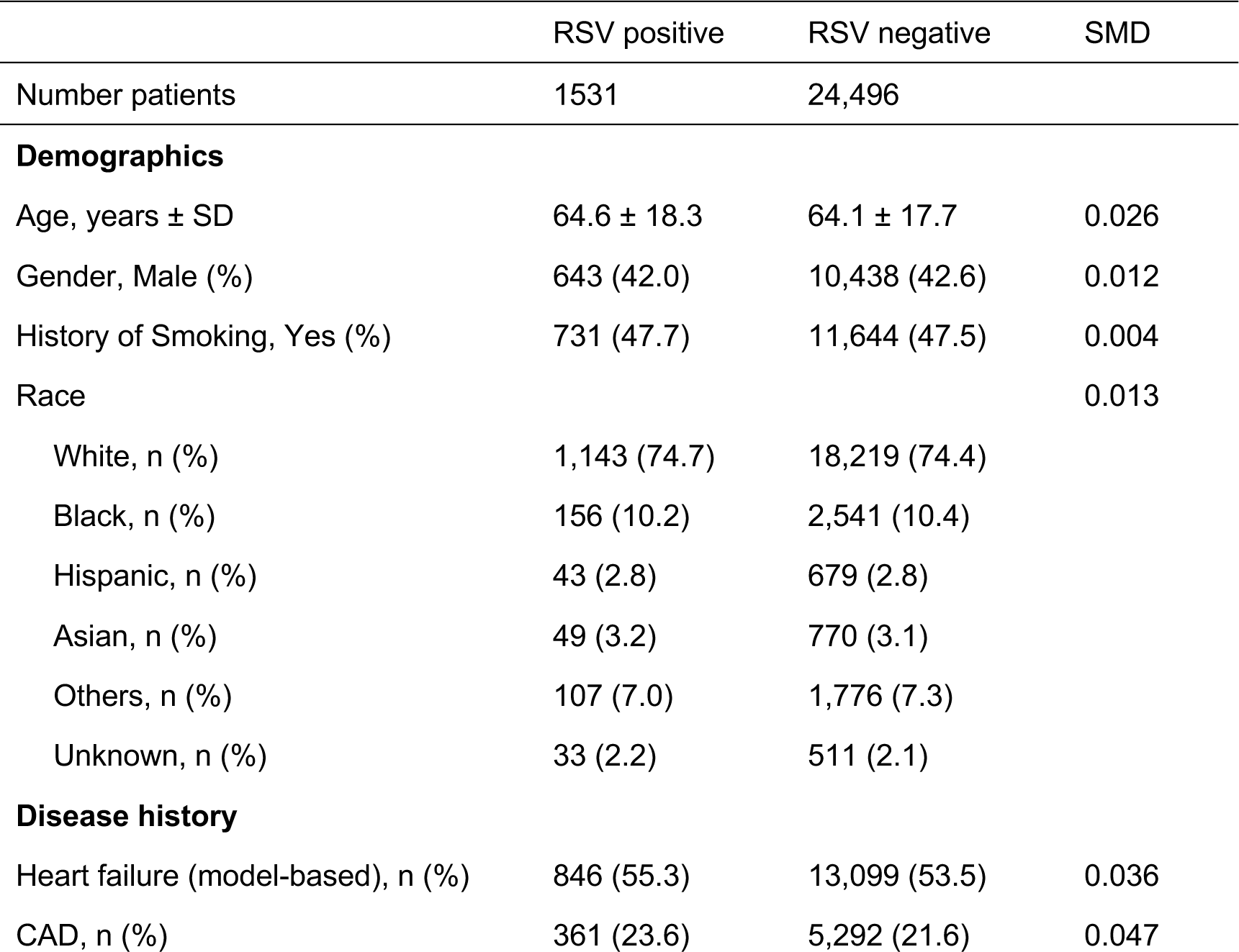
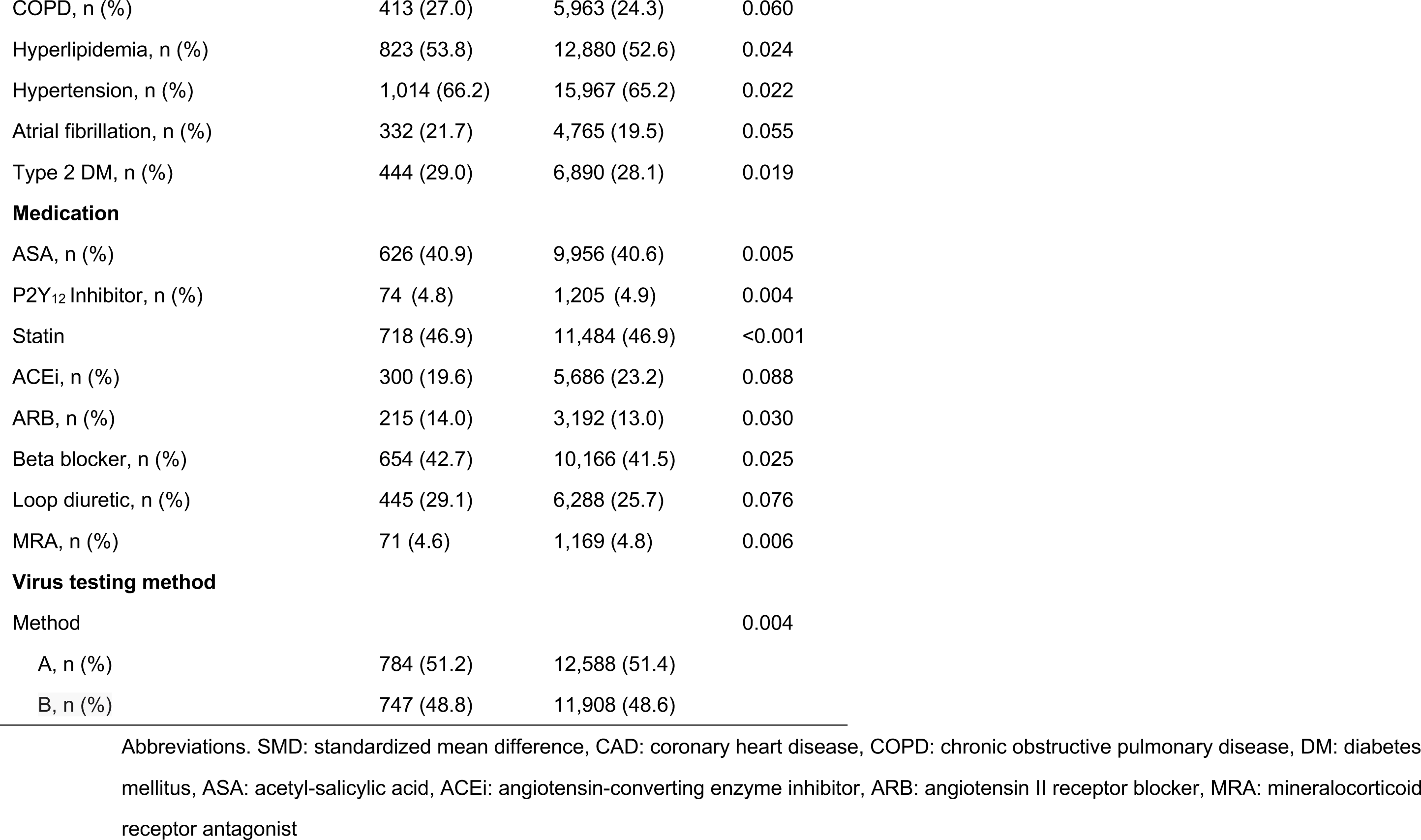
Baseline characteristics for survival analysis of heart failure hospitalization stratified by RSV infection (matched, model-based heart failure history with cutoff 0.24)

**Table S28.** Baseline characteristics for survival analysis of heart failure hospitalization stratified by RSV infection (matched, model-based heart failure history with cutoff 0.41)

|  | RSV positive | RSV negative | SMD |
| --- | --- | --- | --- |
| Number patients | 1531 | 24,496 |  |
| **Demographics** |  |  |  |
| Age, years ± SD | 64.6 ± 18.3 | 64.1 ± 17.7 | 0.026 |
| Gender, Male (%) | 643 (42.0) | 10,450 (42.7) | 0.013 |
| History of Smoking, Yes (%) | 731 (47.7) | 11,650 (47.6) | 0.004 |
| Race |  |  | 0.009 |
| White, n (%) | 1,143 (74.7) | 18,254 (74.5) |  |
| Black, n (%) | 156 (10.2) | 2,540 (10.4) |  |
| Hispanic, n (%) | 43 (2.8) | 695 (2.8) |  |
| Asian, n (%) | 49 (3.2) | 779 (3.2) |  |
| Others, n (%) | 107 (7.0) | 1,724 (7.0) |  |
| Unknown, n (%) | 33 (2.2) | 504 (2.1) |  |
| **Disease history** |  |  |  |
| Heart failure (model-based), n (%) | 744 (48.6) | 11,355 (46.4) | 0.045 |
| CAD, n (%) | 361 (23.6) | 5,320 (21.7) | 0.044 |
| COPD, n (%) | 413 (27.0) | 5,981 (24.4) | 0.059 |
| Hyperlipidemia, n (%) | 823 (53.8) | 12,929 (52.8) | 0.020 |
| Hypertension, n (%) | 1,014 (66.2) | 16,005 (65.3) | 0.019 |
| Atrial fibrillation, n (%) | 332 (21.7) | 4,724 (19.3) | 0.060 |
| Type 2 DM, n (%) | 444 (29.0) | 6,871 (28.0) | 0.021 |
| **Medication** |  |  |  |
| ASA, n (%) | 626 (40.9) | 9,989 (40.8) | 0.002 |
| P2Y<sub>12</sub> Inhibitor, n (%) | 74 (4.8) | 1,211 (4.9) | 0.005 |
| Statin | 718 (46.9) | 11,538 (47.1) | 0.004 |
| ACEi, n (%) | 300 (19.6) | 5,663 (23.1) | 0.086 |
| ARB, n (%) | 215 (14.0) | 3,212 (13.1) | 0.027 |
| Beta blocker, n (%) | 654 (42.7) | 10,138 (41.4) | 0.027 |
| Loop diuretic, n (%) | 445 (29.1) | 6,263 (25.6) | 0.079 |
| MRA, n (%) | 71 (4.6) | 1,165 (4.8) | 0.006 |
| **Virus testing method** |  |  |  |
| Method |  |  | 0.005 |
| A, n (%) | 784 (51.2) | 12,601 (51.2) |  |
| B, n (%) | 747 (48.8) | 11,895 (48.8) |  |

Abbreviations. SMD: standardized mean difference, CAD: coronary heart disease, COPD: chronic obstructive pulmonary disease, DM: diabetes mellitus, ASA: acetyl-salicylic acid, ACEi: angiotensin-converting enzyme inhibitor, ARB: angiotensin II receptor blocker, MRA: mineralocorticoid receptor antagonist

**Table S29.** Baseline characteristics for survival analysis of heart failure hospitalization stratified by RSV infection (matched, model-based heart failure history with cutoff 0.43)

|  | RSV positive | RSV negative | SMD |
| --- | --- | --- | --- |
| Number patients | 1531 | 24,496 |  |
| **Demographics** |  |  |  |
| Age, years ± SD | 64.6 ± 18.3 | 64.1 ± 17.7 | 0.028 |
| Gender, Male (%) | 643 (42.0) | 10,368 (42.3) | 0.007 |
| History of Smoking, Yes (%) | 731 (47.7) | 11,560 (47.2) | 0.011 |
| Race |  |  | 0.012 |
| White, n (%) | 1,143 (74.7) | 18,188 (74.2) |  |
| Black, n (%) | 156 (10.2) | 2,508 (10.2) |  |
| Hispanic, n (%) | 43 (2.8) | 717 (2.9) |  |
| Asian, n (%) | 49 (3.2) | 812 (3.3) |  |
| Others, n (%) | 107 (7.0) | 1,750 (7.1) |  |
| Unknown, n (%) | 33 (2.2) | 521 (2.1) |  |
| **Disease history** |  |  |  |
| Heart failure (model-based), n (%) | 381 (24.9) | 5,232 (21.4) | 0.084 |
| CAD, n (%) | 361 (23.6) | 5,258 (21.5) | 0.051 |
| COPD, n (%) | 413 (27.0) | 5,928 (24.2) | 0.064 |
| Hyperlipidemia, n (%) | 823 (53.8) | 12,870 (52.5) | 0.024 |
| Hypertension, n (%) | 1,014 (66.2) | 16,028 (65.4) | 0.017 |
| Atrial fibrillation, n (%) | 332 (21.7) | 4,730 (19.3) | 0.059 |
| Type 2 DM, n (%) | 444 (29.0) | 6,587 (28.0) | 0.022 |
| **Medication** |  |  |  |
| ASA, n (%) | 626 (40.9) | 9,936 (40.6) | 0.007 |
| P2Y<sub>12</sub> Inhibitor, n (%) | 74 (4.8) | 1,213 (5.0) | 0.005 |
| Statin | 718 (46.9) | 11,488 (46.9) | <0.001 |
| ACEi, n (%) | 300 (19.6) | 5,676 (23.2) | 0.087 |
| ARB, n (%) | 215 (14.0) | 3,213 (13.1) | 0.027 |
| Beta blocker, n (%) | 654 (42.7) | 10,118 (41.3) | 0.029 |
| Loop diuretic, n (%) | 445 (29.1) | 6,246 (25.5) | 0.080 |
| MRA, n (%) | 71 (4.6) | 1,154 (4.7) | 0.003 |
| **Virus testing method** |  |  |  |
| Method |  |  | 0.006 |
| A, n (%) | 784 (51.2) | 12,623 (51.5) |  |
| B, n (%) | 747 (48.8) | 11,873 (48.5) |  |

Abbreviations. SMD: standardized mean difference, CAD: coronary heart disease, COPD: chronic obstructive pulmonary disease, DM: diabetes mellitus, ASA: acetyl-salicylic acid, ACEi: angiotensin-converting enzyme inhibitor, ARB: angiotensin II receptor blocker, MRA: mineralocorticoid receptor antagonist

**Table S30.** Baseline characteristics for survival analysis of heart failure hospitalization stratified by influenza infection (matched, model- based heart failure history with cutoff 0.24)

|  | influenza positive | influenza negative | SMD |
| --- | --- | --- | --- |
| Number patients | 5138 | 25,690 |  |
| **Demographics** |  |  |  |
| Age, years ± SD | 55.2 ± 19.8 | 55.2 ± 19.6 | 0.002 |
| Gender, Male (%) | 1,867 (36.3) | 9,482 (36.9) | 0.012 |
| History of Smoking, Yes (%) | 1,720 (33.5) | 8,839 (34.4) | 0.020 |
| Race |  |  | 0.022 |
| White, n (%) | 3,702 (72.1) | 18,553 (72.2) |  |
| Black, n (%) | 424 (8.3) | 2,166 (8.4) |  |
| Hispanic, n (%) | 169 (3.3) | 821 (3.2) |  |
| Asian, n (%) | 220 (4.3) | 1,042 (4.1) |  |
| Others, n (%) | 420 (8.2) | 2,167 (8.4) |  |
| Unknown, n (%) | 203 (4.0) | 941 (3.7) |  |
| **Disease history** |  |  |  |
| Heart failure (model-based), n (%) | 1,161 (22.6) | 5,774 (22.5) | 0.003 |
| CAD, n (%) | 541 (10.5) | 2,750 (10.7) | 0.006 |
| COPD, n (%) | 639 (12.4) | 3,328 (13.0) | 0.016 |
| Hyperlipidemia, n (%) | 2,035 (39.6) | 10,091 (39.3) | 0.007 |
| Hypertension, n (%) | 2,179 (42.4) | 11,042 (43.0) | 0.012 |
| Atrial fibrillation, n (%) | 460 (9.0) | 2,332 (9.1) | 0.004 |
| Type 2 DM, n (%) | 864 (16.8) | 4,335 (16.9) | 0.002 |
| **Medication** |  |  |  |
| ASA, n (%) | 1,326 (25.8) | 6,764 (26.3) | 0.012 |
| P2Y<sub>12</sub> Inhibitor, n (%) | 145 (2.8) | 772 (3.0) | 0.011 |
| Statin | 1,577 (30.7) | 7,691 (29.9) | 0.016 |
| ACEi, n (%) | 779 (15.2) | 3,962 (15.4) | 0.007 |
| ARB, n (%) | 451 (8.8) | 2,234 (8.7) | 0.003 |
| Beta blocker, n (%) | 1137 (22.1) | 6,073 (23.6) | 0.036 |
| Loop diuretic, n (%) | 529 (10.3) | 2,710 (10.5) | 0.008 |
| MRA, n (%) | 117 (2.3) | 632 (2.5) | 0.012 |
| **Virus testing method** |  |  |  |
| Method |  |  | 0.039 |
| A, n (%) | 3,133 (61.0) | 15,312 (59.6) |  |
| B, n (%) | 725 (14.1) | 3,955 (15.4) |  |
| C, n (%) | 720 (14.0) | 3,659 (14.2) |  |
| D, n (%) | 560 (10.9) | 2,764 (10.8) |  |

Abbreviations. SMD: standardized mean difference, CAD: coronary heart disease, COPD: chronic obstructive pulmonary disease, DM: diabetes mellitus, ASA: acetyl-salicylic acid, ACEi: angiotensin-converting enzyme inhibitor, ARB: angiotensin II receptor blocker, MRA: mineralocorticoid receptor antagonist

**Table S31.**
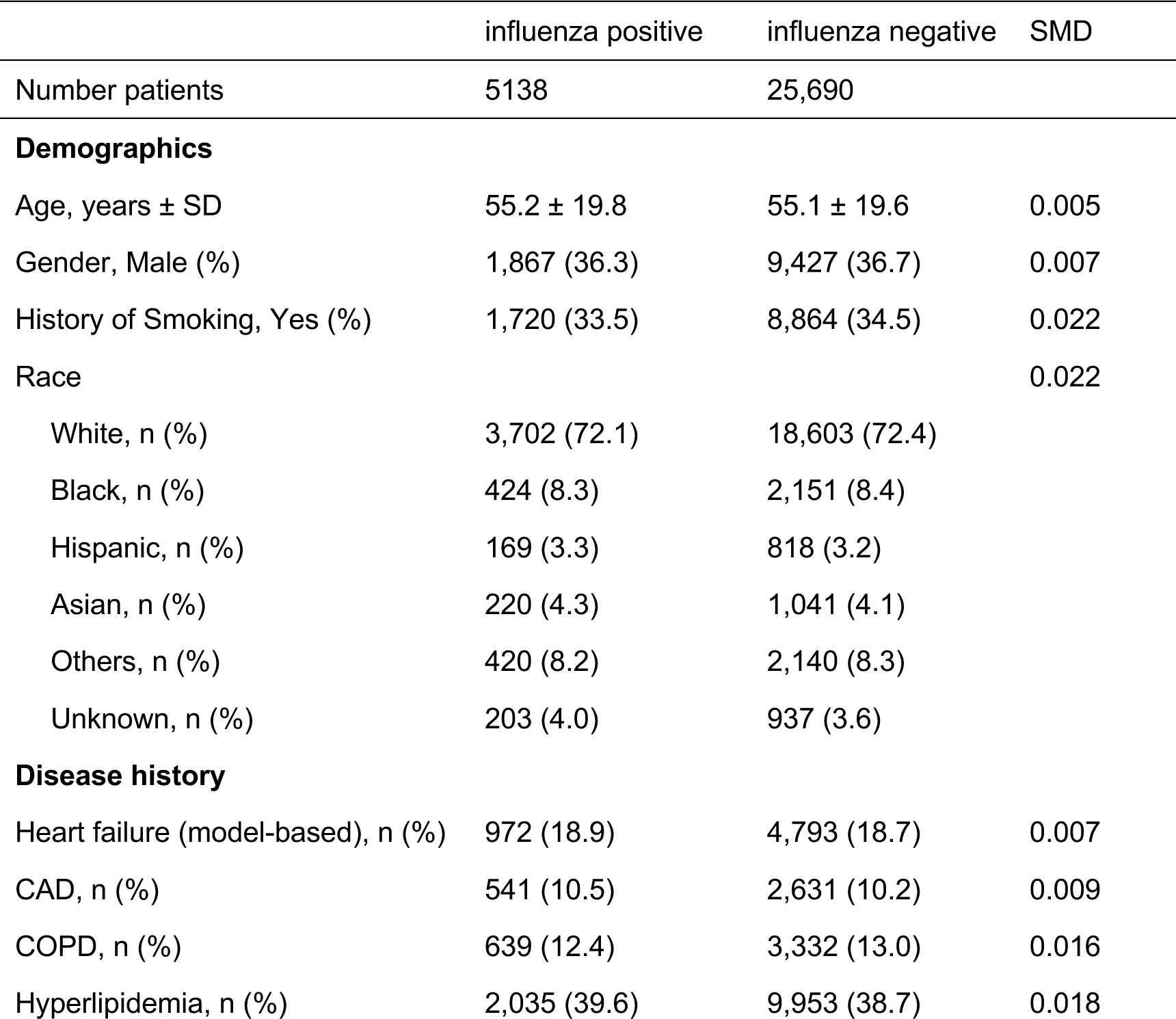
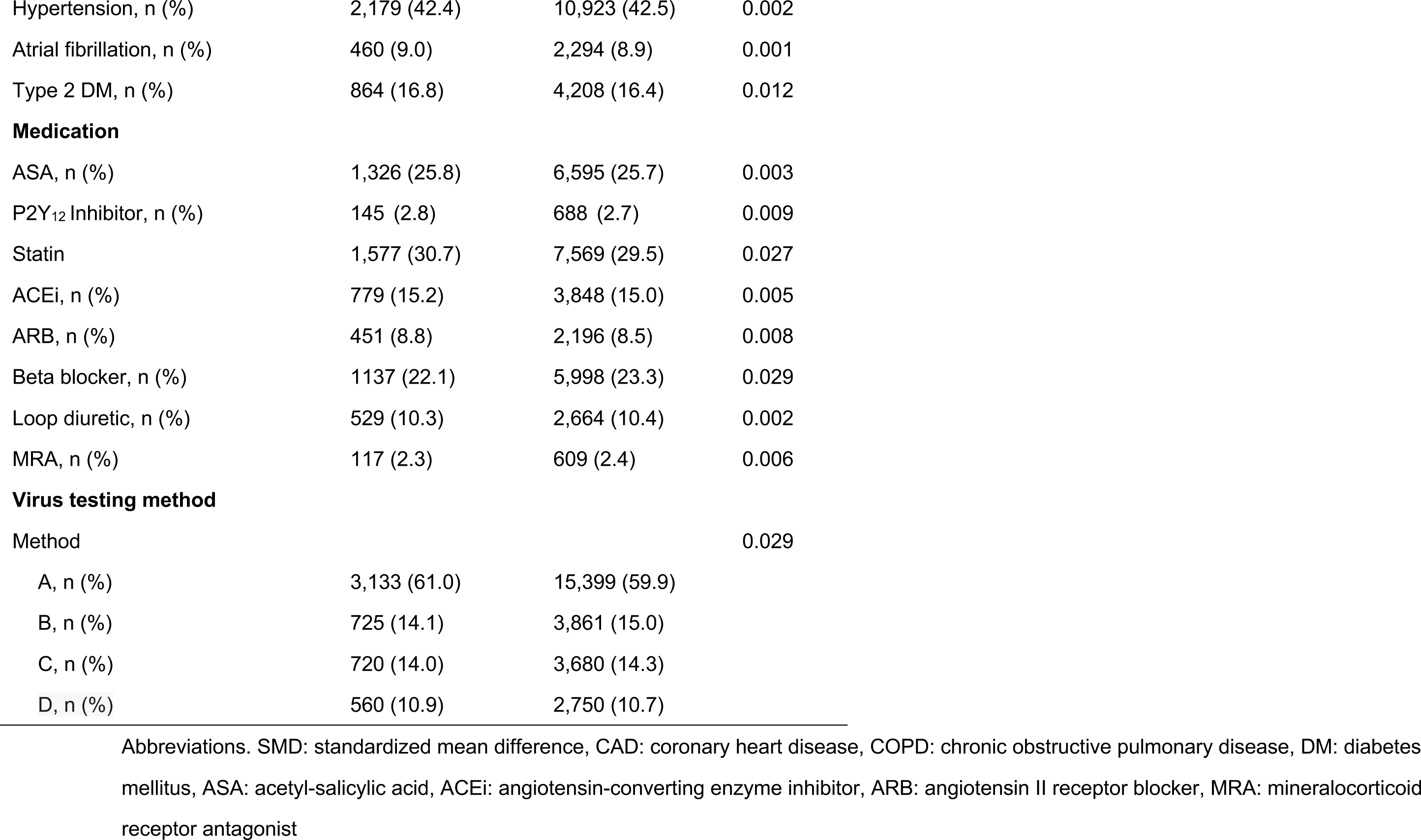
Baseline characteristics for survival analysis of heart failure hospitalization stratified by influenza infection (matched, model- based heart failure history with cutoff 0.41)

**Table S32.** Baseline characteristics for survival analysis of heart failure hospitalization stratified by influenza infection (matched, model- based heart failure history with cutoff 0.43)

|  | influenza positive | influenza negative | SMD |
| --- | --- | --- | --- |
| Number patients | 5138 | 25,690 |  |
| **Demographics** |  |  |  |
| Age, years ± SD | 55.2 ± 19.8 | 55.1 ± 19.6 | 0.003 |
| Gender, Male (%) | 1,867 (36.3) | 9,342 (36.4) | 0.001 |
| History of Smoking, Yes (%) | 1,720 (33.5) | 8,662 (33.7) | 0.005 |
| Race |  |  | 0.022 |
| White, n (%) | 3,702 (72.1) | 18,538 (72.2) |  |
| Black, n (%) | 424 (8.3) | 2,178 (8.5) |  |
| Hispanic, n (%) | 169 (3.3) | 849 (3.3) |  |
| Asian, n (%) | 220 (4.3) | 1,040 (4.0) |  |
| Others, n (%) | 420 (8.2) | 2,147 (8.4) |  |
| Unknown, n (%) | 203 (4.0) | 938 (3.7) |  |
| **Disease history** |  |  |  |
| Heart failure (model-based), n (%) | 401 (7.8) | 2,049 (8.0) | 0.006 |
| CAD, n (%) | 541 (10.5) | 2,719 (10.6) | 0.002 |
| COPD, n (%) | 639 (12.4) | 3,253 (12.7) | 0.007 |
| Hyperlipidemia, n (%) | 2,035 (39.6) | 10,015 (39.0) | 0.013 |
| Hypertension, n (%) | 2,179 (42.4) | 10,888 (42.4) | 0.001 |
| Atrial fibrillation, n (%) | 460 (9.0) | 2,338 (9.1) | 0.005 |
| Type 2 DM, n (%) | 864 (16.8) | 4,302 (16.7) | 0.002 |
| **Medication** |  |  |  |
| ASA, n (%) | 1,326 (25.8) | 6,616 (25.8) | 0.001 |
| P2Y<sub>12</sub> Inhibitor, n (%) | 145 (2.8) | 707 (2.8) | 0.004 |
| Statin | 1,577 (30.7) | 7,572 (29.5) | 0.027 |
| ACEi, n (%) | 779 (15.2) | 3,882 (15.1) | 0.001 |
| ARB, n (%) | 451 (8.8) | 2,205 (8.6) | 0.007 |
| Beta blocker, n (%) | 1137 (22.1) | 6,151 (23.9) | 0.043 |
| Loop diuretic, n (%) | 529 (10.3) | 2,704 (10.5) | 0.008 |
| MRA, n (%) | 117 (2.3) | 657 (2.6) | 0.018 |
| **Virus testing method** |  |  |  |
| Method |  |  | 0.032 |
| A, n (%) | 3,133 (61.0) | 15,449 (60.1) |  |
| B, n (%) | 725 (14.1) | 3,834 (14.9) |  |
| C, n (%) | 720 (14.0) | 3,738 (14.6) |  |
| D, n (%) | 560 (10.9) | 2,669 (10.4) |  |

Abbreviations. SMD: standardized mean difference, CAD: coronary heart disease, COPD: chronic obstructive pulmonary disease, DM: diabetes mellitus, ASA: acetyl-salicylic acid, ACEi: angiotensin-converting enzyme inhibitor, ARB: angiotensin II receptor blocker, MRA: mineralocorticoid receptor antagonist

## References

1. O’Malley KJ, Cook KF, Price MD, Wildes KR, Hurdle JF, Ashton CM. Measuring Diagnoses: ICD Code Accuracy. Health Serv Res. 2005;40:1620–1639.

2. Wei W-Q, Teixeira PL, Mo H, Cronin RM, Warner JL, Denny JC. Combining billing codes, clinical notes, and medications from electronic health records provides superior phenotyping performance. J Am Med Inform Assn. 2016;23:e20–e27.

3. Lash TL, Fox MP, MacLehose RF, Maldonado G, McCandless LC, Greenland S. Good practices for quantitative bias analysis. Int J Epidemiol. 2014;43:1969–1985.

4. Patadia VK, Coloma P, Schuemie MJ, Herings R, Gini R, Mazzaglia G, Picelli G, Fornari C, Pedersen L, Lei J van der, Sturkenboom M, Trifirò G, consortium E-A. Using real-world healthcare data for pharmacovigilance signal detection – the experience of the EU-ADR project. Expert Rev Clin Phar. 2014;8:95–102.

5. Falsey AR, Hennessey PA, Formica MA, Cox C, Walsh EE. Respiratory Syncytial Virus Infection in Elderly and High-Risk Adults. New Engl J Med. 2005;352:1749–1759.

6. Kytömaa S, Hegde S, Claggett B, Udell JA, Rosamond W, Temte J, Nichol K, Wright JD, Solomon SD, Vardeny O. Association of Influenza-like Illness Activity With Hospitalizations for Heart Failure. Jama Cardiol. 2019;4:363–369.

7. Kwong JC, Schwartz KL, Campitelli MA, Chung H, Crowcroft NS, Karnauchow T, Katz K, Ko DT, McGeer AJ, McNally D, Richardson DC, Rosella LC, Simor A, Smieja M, Zahariadis G, Gubbay JB. Acute Myocardial Infarction after Laboratory-Confirmed Influenza Infection. New Engl J Medicine. 2018;378:345–353.

8. Barnes M, Heywood AE, Mahimbo A, Rahman B, Newall AT, Macintyre CR. Acute myocardial infarction and influenza: a meta-analysis of case–control studies. Heart. 2015;101:1738.

9. Inciardi RM, Lupi L, Zaccone G, Italia L, Raffo M, Tomasoni D, Cani DS, Cerini M, Farina D, Gavazzi E, Maroldi R, Adamo M, Ammirati E, Sinagra G, Lombardi CM, Metra M. Cardiac Involvement in a Patient With Coronavirus Disease 2019 (COVID-19). Jama Cardiol. 2020;5:819–824.

10. Doyen D, Moceri P, Ducreux D, Dellamonica J. Myocarditis in a patient with COVID-19: a cause of raised troponin and ECG changes. Lancet. 2020;395:1516.

11. Madjid M, Safavi-Naeini P, Solomon SD, Vardeny O. Potential Effects of Coronaviruses on the Cardiovascular System. Jama Cardiol. 2020;5:831–840.

12. Boehme AK, Luna J, Kulick ER, Kamel H, Elkind MSV. Influenza-like illness as a trigger for ischemic stroke. Ann Clin Transl Neur. 2018;5:456–463.

13. Hicks KA, Mahaffey KW, Mehran R, Nissen SE, Wiviott SD, Dunn B, Solomon SD, Marler JR, Teerlink JR, Farb A, Morrow DA, Targum SL, Sila CA, Hai MTT, Jaff MR, Joffe HV, Cutlip DE, Desai AS, Lewis EF, Gibson CM, Landray MJ, Lincoff AM, White CJ, Brooks SS, Rosenfield K, Domanski MJ, Lansky AJ, McMurray JJV, Tcheng JE, Steinhubl SR, Burton P, Mauri L, O’Connor CM, Pfeffer MA, Hung HMJ, Stockbridge NL, Chaitman BR, Temple RJ, (SCTI) SDC for CTI. 2017 Cardiovascular and Stroke Endpoint Definitions for Clinical Trials. Circulation. 2018;137:961–972.

14. Berkson J. Limitations of the Application of Fourfold Table Analysis to Hospital Data.,. Int J Epidemiol. 2014;43:511–515.

15. Rosenbaum PR, Rubin DB. Assessing Sensitivity to an Unobserved Binary Covariate in an Observational Study with Binary Outcome. J Royal Statistical Soc Ser B Methodol. 1983;45:212–218.

16. Dafny LS. How Do Hospitals Respond to Price Changes? Am Econ Rev. 2005;95:1525–1547.

17. Hripcsak G, Albers DJ. Next-generation phenotyping of electronic health records. J Am Med Inform Assn. 2012;20:117–121.

18. Usama M, Ahmad B, Wan J, Hossain MS, Alhamid MF, Hossain MA. Deep Feature Learning for Disease Risk Assessment Based on Convolutional Neural Network With Intra-Layer Recurrent Connection by Using Hospital Big Data. Ieee Access. 2018;6:67927–67939.

19. Magna AAR, Allende-Cid H, Taramasco C, Becerra C, Figueroa RL. Application of Machine Learning and Word Embeddings in the Classification of Cancer Diagnosis Using Patient Anamnesis. Ieee Access. 2020;8:106198–106213.

20. Ivey KS, Edwards KM, Talbot HK. Respiratory Syncytial Virus and Associations With Cardiovascular Disease in Adults. J Am Coll Cardiol. 2018;71:1574–1583.

21. Ukimura A, Satomi H, Ooi Y, Kanzaki Y. Myocarditis Associated with Influenza A H1N1pdm2009. Influ Res Treat. 2012;2012:351979.

22. Drozd M, Garland E, Walker AMN, Slater TA, Koshy A, Straw S, Gierula J, Paton M, Lowry J, Sapsford R, Witte KK, Kearney MT, Cubbon RM. Infection-Related Hospitalization in Heart Failure With Reduced Ejection Fraction. Circulation Hear Fail. 2020;13:e006746.

23. Hernán MA, Hernández-Díaz S, Robins JM. A Structural Approach to Selection Bias. Epidemiology. 2004;15:615–625.

24. Shi S, Qin M, Shen B, Cai Y, Liu T, Yang F, Gong W, Liu X, Liang J, Zhao Q, Huang H, Yang B, Huang C. Association of Cardiac Injury With Mortality in Hospitalized Patients With COVID-19 in Wuhan, China. Jama Cardiol. 2020;5:802–810.

25. Honnibal M, Montani I. SpaCy 2: Natural language understanding with bloom embeddings, convolutional neural networks and incremental parsing. To appear. 2017;7.

26. Bojanowski P, Grave E, Joulin A, Mikolov T. Enriching word vectors with subword information. Transactions of the Association for Computational Linguistics. 2017;5:135–146.

27. Ho DE, Imai K, King G, Stuart EA. Matching as Nonparametric Preprocessing for Reducing Model Dependence in Parametric Causal Inference. Polit Anal. 2007;15:199–236.

